# Determining context-specific economically feasible age ranges for female HPV catch-up vaccination in LMICs: a model-based health economic assessment

**DOI:** 10.64898/2026.03.26.26348394

**Authors:** Irene Man, Abrham Wondimu, Damien Georges, Rachel Wittenauer, Alina Macacu, Ahmad Fuady, Andrea Gini, Iacopo Baussano

## Abstract

To support the WHO global strategy to accelerate cervical cancer elimination in low- and middle-income countries (LMICs), we conducted a comprehensive health-economic impact assessment of HPV catch-up vaccination across different age ranges up to age 30, using IARC’s METHIS modelling platform. A one-time catch-up campaign (targeting HPV 16/18; 90% coverage) in 2026 across 132 LMICs among females aged 9-30 could prevent 6.93 million cervical cancer cases and 3.56 million deaths, on top of 2.70 and 1.57 million cases and deaths averted by past vaccination, requiring 951 million doses and 11,349 million US$. Median country-specific costs represented 0.17% of five-year health expenditure and 19.7% of immunization budgets. Among six countries with diverse epidemiological profiles and sufficient data for cost-effectiveness analyses, catch-up was cost-effective until age 30 in all but one country, where it was cost-effective until age 23 only. Female catch-up could be cost-effective and impactful for cervical cancer prevention in LMICs.

## Introduction

Despite progressive expansion of human papillomavirus (HPV) vaccination introduction globally over the past decade, approximately 15% of women worldwide still reside in countries where the HPV vaccine has not yet been introduced, most in low-and middle-income countries (LMICs).^1^ Even where implemented, HPV vaccination coverage is often suboptimal, leaving many girls and young women unprotected.

Catch-up vaccination, in some settings also known as multiple-age cohort (MAC) vaccination, offers a vital opportunity to protect those who missed routine immunization. Modelling^2,3^ and real-world evidence^4,5^ from diverse settings indicate that including older women in catch-up programmes markedly reduces HPV incidence and accelerates cervical cancer elimination. Yet, few LMICs have adopted such strategies; some have targeted girls only up to age 14 years, with only few rare exceptions extending to ages 18 years, e.g., in Bhutan and Rwanda.^5^ Recent developments, including the addition of newly licenced HPV vaccines, increases in production capacity by current manufacturers, and an endorsement of single-dose schedule by WHO,^6,7^ provide favourable conditions to expand HPV vaccine catch-up without compromising the coverage in girls of the primary target age group of 9–14 years.

Multiple modelling studies have clearly indicated that, after the primary target group of girls below aged 9-14, older girls and women up to age 25, or even age 30, depending on the epidemiological profile, would be the next most efficient vaccination target in terms of maximizing the number of cancers prevented per dose.^3,8,9^ However, when defining exact country-specific policies, additional finetuning is often required to decide on the exact age range and cohorts to vaccinate, accounting for the available resources. While HPV vaccination in the primary target age group has been demonstrated as a ‘best buy’ intervention^10,11^, few studies have examined until which age it remains cost-effective, considering single-dose schedule, and taking into account the resource constraints in LMICs.^12,13^ More comprehensive projections of health-economic impact of catch-up HPV vaccination could guide policy decisions, especially under the current uncertainties around global and local funding for public health programmes.^14^

Given these gaps and the growing momentum to expand HPV vaccination, this study aimed to identify context-specific age ranges for HPV catch-up vaccination in women across LMICs accounting for different epidemiological profiles, economic conditions, and historic vaccination coverage. Using IARC’s METHIS modelling platform^15^, we estimated the health and budget impact of implementing a one-time catch-up vaccination in women across 132 LMICs, considering different maximum age ranges for catch-up up to age 30. For a selection of six LMICs with the required cervical cancer treatment costs and from diverse epidemiological profiles, we also conducted cost-effectiveness analyses.

## Results

### Health impact

Past vaccination modelled (with coverage as reported on WHO HPV Dashboard^1^) in the 132 LMICs was estimated to prevent 2.70 million (95% Uncertainty Interval (UI): 2.68–2.73) cervical cancer cases and 1.57 million (95%UI: 1.55–1.58) deaths over the lifetime of the birth cohorts aged 9-30 years in 2026 (Figure 1). A one-time female catch-up HPV vaccination intervention in these birth cohorts (with a vaccine targeting HPV 16/18 and 90% coverage) was estimated to prevent additional 6.93 million (95%UI: 6.85–7.03) cases and 3.56 million (95%UI: 3.50–3.62) deaths, increasing the total percentage of cervical cancer cases averted from 14.1% to 50.2%. As expected, the impact declined with increasing age, as older cohorts were more likely to have been exposed to HPV through sexual activity. Among the WHO regions, the largest additional number of cervical cancers averted was in the South-East Asia Region (2.35 million cases, 95%UI: 2.35-2.36), closely followed by the African Region (2.14 million cases, 95%UI: 2.06-2.23), and the smallest in the European Region (0.151 million cases, 95%UI: 0.150-0.152). Expressed as percentage of the local burden, the additional impact was the largest at around 47% in the Western Pacific, Eastern Mediterranean, and South-East Asia Regions, and the smallest in the Americas Region. Estimates by country and different maximum catch-up age are reported in Table S1-2 and Figure S1-2.

**Figure 1.**
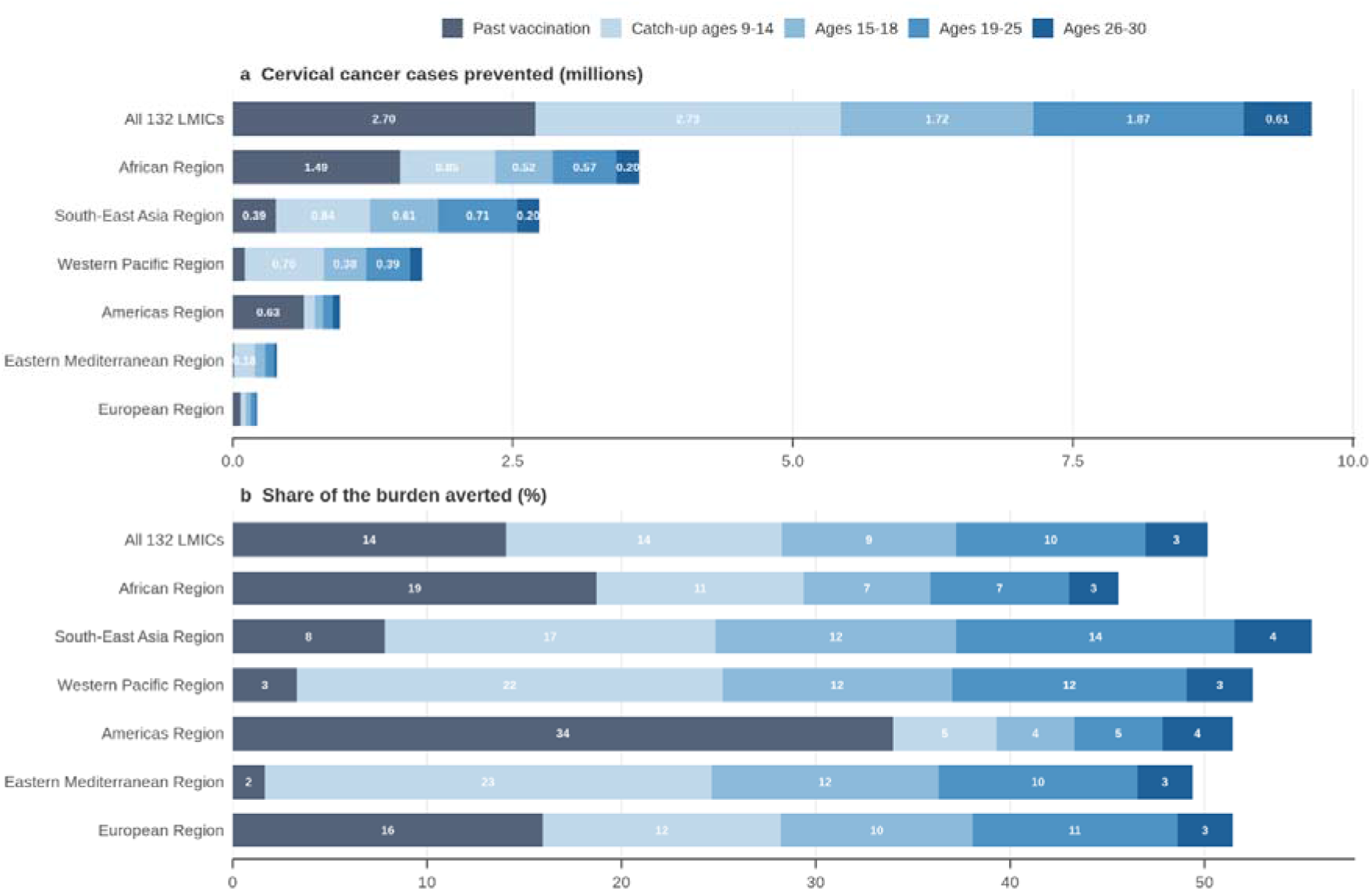
Cervical cancer cases averted by past vaccination (grey) and a one-time female catch-up HPV vaccination (with a vaccine targeting HPV16/18 and 90% coverage) up to 14, 18, 25, or 30 in the lifetime of birth cohort aged 9–30 years in 2026, by WHO region in panel a. In panel b, the estimates are in percentages of the total number of cervical cancer cases expected without vaccination. The age bands are incremental values: each is what it adds to the bands below it, so that a whole bar is past vaccination plus a catch-up campaign reaching age 30. Past vaccination is each country’s vaccination history together with its routine programme continuing at the last observed coverage. See Methods for the exact definition of “past vaccination”.

### Budget impact

The total budget needed in all 132 modelled LMICs for female catch-up vaccination up to age 18 and 30 were, respectively, 4,705 and 11,349 million US dollars (US$), requiring 405 and 951 million HPV vaccine doses. As percentage of the five-year Current Health Expenditure (CHE), the median country budgets needed for catch-up were 0.05% (Interquartile Range (IQR): 0.02%–0.16%) and 0.17% (IQR: 0.08%–0.40%), respectively (Figure 2). As percentage of the five-year immunization budget, it were 6.4% (IQR: 3.3%–13.4%) 19.7% (IQR: 12.0%–32.9%), respectively. There was variation in required campaign budget relative to CHE and immunization budget by WHO region and category of procurement route. For example, the proportions were comparatively high for Gavi-eligible countries, as these countries often have lower health budgets. Country-specific estimates as percentages and in US$ for different maximum catch-up ages are reported in Table S3 and Figure S2, respectively.

**Figure 2.**
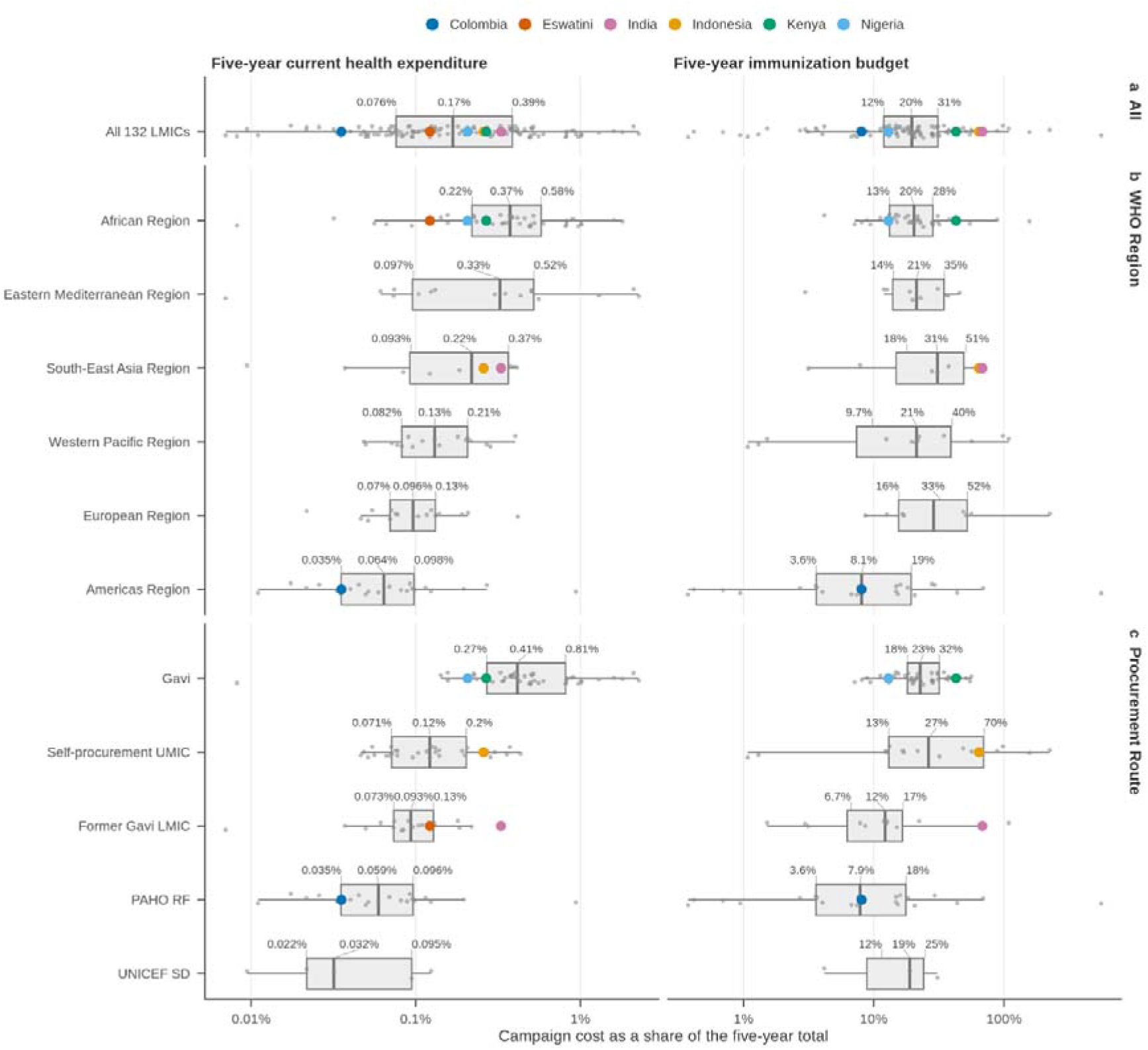
Estimated costs of female catch-up HPV vaccination up to age 30 years (assuming a vaccine targeting HPV 16/18 and 90% vaccine coverage) as a percentage of five-year current health expenditure (CHE) and immunization budgets across countries. Both vaccine cost and delivery cost are included. Financial cost (and not economic cost) is considered. Results of the six countries selected for cost-effectiveness analysis are indicated with coloured circles. Boxes are the interquartile range with the median marked and the three quartile values printed above; each grey point is one country. Both axes are logarithmic, because the shares span three orders of magnitude. Procurement route sets the price a country pays for each vaccine dose. Countries without a usable denominator were omitted rather than shown at zero: Two had no current health expenditure figure and 31 no usable immunization budget — 23 reported none, 6 reported one outside 0.03% to 20% of their own current health expenditure, one reported zero and one could not be rebased to 2024 dollars. Eswatini was among those 6, so only 5 of the six countries selected for cost-effectiveness analyses appear in the right-hand panel.

In general, delivery costs accounted for a smaller share of the total budget for catch-up vaccination programmes than vaccine procurement costs, with a median estimate of 30.6% (IQR: 16.4%–31.8%) for maximum catch-up age of 18 and increased to 35.3% (IQR: 20.0%–36.4%) for maximum age 30, due to higher delivery costs assumed at higher ages of vaccination (Figure S3). Gavi co-financing was estimated at 495 million US$ for catch-up vaccination up to age 18 years, the maximum age for which Gavi supports MAC campaigns under its most recent vaccine funding guidelines (Gavi 6.0) ^16^. This covered 49.0% of the vaccination costs of the 56 eligible countries and 10.5% of the total across all 132 countries. The financial contribution an individual country receives is set by its co-financing phase: Gavi covered a median 67.6% of vaccination costs in the 26 countries in the “initial self-financing” phase, 38.1% in the 23 in “preparatory transition”, and 18.6% in the 7 in “accelerated transition” (Figure S4).

### Health impact and cost-effectiveness in six selected countries

Six countries with high-quality primary data of cervical cancer treatment costs were selected for country-specific cost-effectiveness analysis, each from a distinct epidemiological profile as identified in Man et al.^16^: Colombia, Eswatini, India, Indonesia, Kenya, and Nigeria. For all six modelled countries, the total impact of past vaccination and the one-time catch-up combined (again assuming a vaccine targeting HPV 16/18 and 90% coverage) for six countries was all around 65-85% cervical cancer cases averted in the primary target age group (9-14) and decreased to 10-20% at age 30 (Figure 3). In some cases, e.g., in Colombia for cohort aged 20-28 in 2026, high impact was already achieved due to high coverage in the past. The impact per 100,000 women alive in 2026 (Figure S5) and the total absolute impact (Table 1, Figure S1) varied more widely across the countries; the former mainly driven by high baseline cervical cancer incidence (e.g., in Eswatini) while the latter is also driven by large population size (e.g., India).

**Figure 3.**
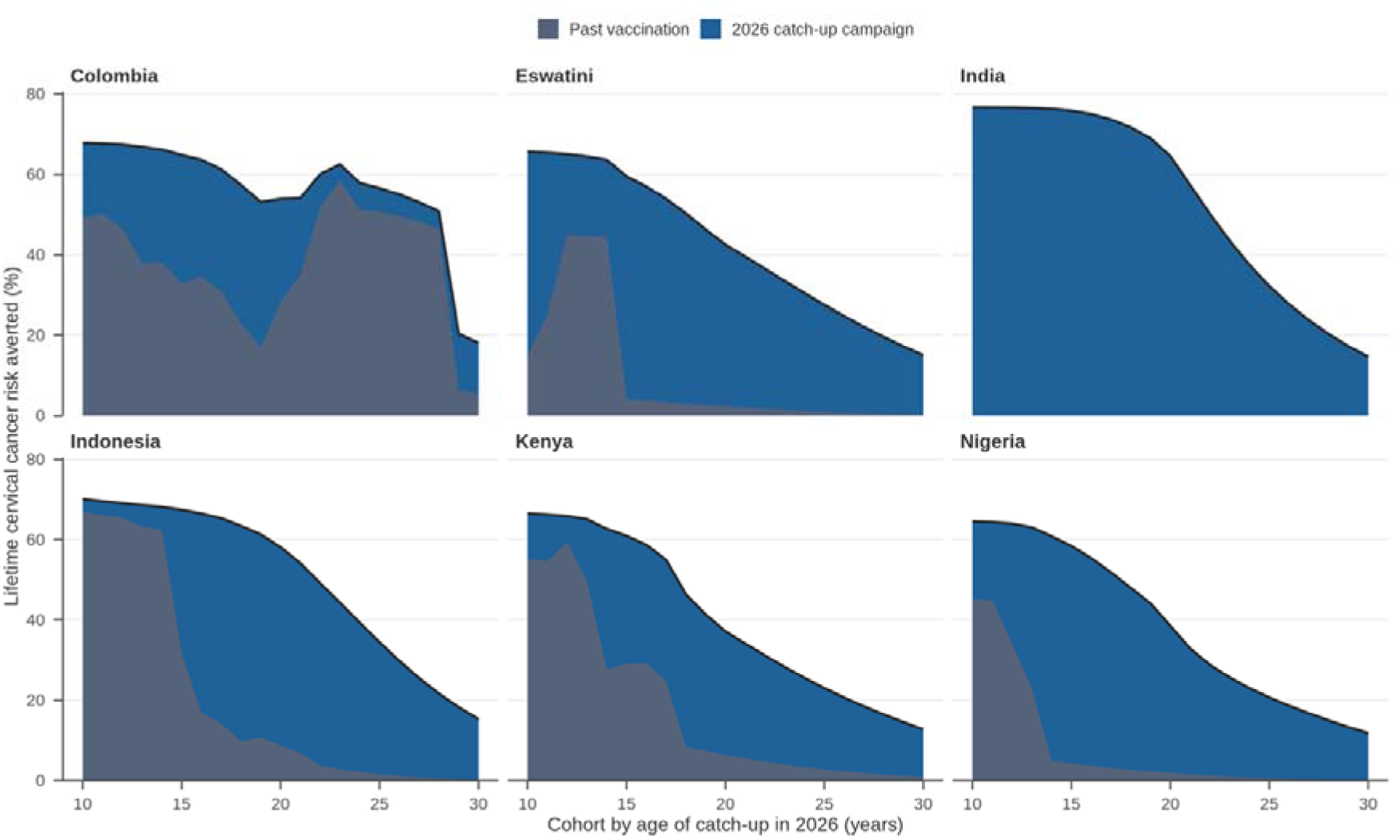
Share of the cervical cancer cases averted by past vaccination (grey) and a one-time female catch-up HPV vaccination (blue, with a vaccine targeting HPV16/18 and 90% coverage) by birth cohort aged 10–30 years in 2026 in the x-axis, in the six countries selected for cost-effectiveness analysis. Past vaccination is each country’s vaccination history together with its routine programme continuing at the last observed coverage. See Methods for the exact definition of “past vaccination”.

**Table 1:** Health and economic impact of HPV catch-up vaccination in the six countries from distinct epidemiological profiles selected for cost-effectiveness analyses based on sufficient cervical cancer treatment cost data. The impact is assessed over the lifetime of women in birth cohorts aged 9-30 years in 2026, assuming a vaccine targeting HPV16/18 and 90% coverage. ¶ Health impacts are calculated using a 0% discount rate, while costs are discounted at 3%. † Economic cost (not financial cost), which was the input for the cost-effectiveness analysis. ‡ ICER estimates are the same as those displayed in Figure 4. Hence, they are ICER estimates at the exact maximum catch-up age. — Cost of past vaccination was not estimated, as historical dose counts and the prices paid for them are not available, and past vaccination is a sunk cost that does not affect the economics of the catch-up campaign.

| Country | Past vaccination / Maximum catch-up age | Total health impact, median (95% UI) ¶ | | | Total cost, median (95% UI) ¶ | | ICER (US\$ / DALY averted) ‡ |
| --- | --- | --- | --- | --- | --- | --- | --- |
| | | Cases averted (thousands) | Deaths averted (thousands) | DALYs averted (thousands) | Treatment cost saved (US\$ million) | Vaccination cost (US\$ million) † | |
| Colombia | Past vaccination | 46.0 (44.7–48.4) | 24.2 (23.5–25.5) | 615.1 (598.2–647.7) | 61.7 (60.0–64.8) | — | Reference |
|  | 14 | 7.8 (7.3–7.9) | 4.1 (3.8–4.1) | 103.9 (96.8–104.8) | 8.7 (8.1–8.8) | 14.0 (14.0–14.0) | 66 (62–76) |
|  | 18 | 14.7 (13.8–15.0) | 7.6 (7.2–7.8) | 194.9 (183.8–200.1) | 17.2 (16.3–17.7) | 30.7 (30.7–30.7) | 106 (94–130) |
|  | 25 | 20.1 (18.8–20.7) | 10.5 (9.8–10.8) | 266.9 (250.3–276.1) | 24.7 (23.1–25.6) | 53.4 (53.4–53.4) | 372 (326–450) |
|  | 30 | 22.3 (20.8–23.1) | 11.7 (10.9–12.1) | 297.1 (277.4–307.5) | 28.4 (26.4–29.5) | 74.8 (74.8–74.8) | 706 (627–835) |
| Eswatini | Past vaccination | 2.0 (2.0–2.0) | 1.2 (1.2–1.2) | 29.6 (29.3–30.2) | 29.4 (29.0–30.0) | — | Reference |
|  | 14 | 2.2 (2.2–2.2) | 1.3 (1.3–1.3) | 33.0 (32.7–33.2) | 31.9 (31.6–32.2) | 0.5 (0.5–0.5) | -1,024 (-1,026–-1,023) |
|  | 18 | 4.2 (4.2–4.3) | 2.5 (2.5–2.6) | 63.0 (62.6–63.9) | 65.1 (64.7–66.2) | 1.1 (1.1–1.1) | -1,127 (-1,129–-1,126) |
|  | 25 | 6.4 (6.3–6.6) | 3.8 (3.8–4.0) | 95.5 (94.0–98.9) | 106.2 (104.3–110.4) | 2.5 (2.5–2.5) | -1,312 (-1,315–-1,310) |
|  | 30 | 7.2 (7.0–7.5) | 4.3 (4.2–4.5) | 107.4 (105.4–111.8) | 123.5 (121.1–129.2) | 3.4 (3.4–3.4) | -1,449 (-1,456–-1,447) |
| India | Past vaccination | 0.0 (0.0–0.0) | 0.0 (0.0–0.0) | 0.0 (0.0–0.0) | 0.0 (0.0–0.0) | — | Reference |
|  | 14 | 738.4 (737.5–739.5) | 344.1 (343.7–344.6) | 7,919.8 (7,910.4–7,931.0) | 787.3 (786.3–788.6) | 705.8 (705.8–705.8) | 4 (4–4) |
|  | 18 | 1,159.6 (1,158.6–1,160.8) | 540.3 (539.9–540.9) | 12,436.1 (12,425.8–12,449.2) | 1,301.5 (1,300.3–1,303.0) | 1,233.2 (1,233.2–1,233.2) | 4 (4–4) |
|  | 25 | 1,608.2 (1,605.9–1,611.1) | 749.4 (748.3–750.8) | 17,246.7 (17,222.4–17,278.5) | 1,919.7 (1,916.6–1,923.7) | 2,382.0 (2,382.0–2,382.0) | 282 (278–286) |
|  | 30 | 1,726.2 (1,723.0–1,730.5) | 804.2 (802.7–806.2) | 18,504.7 (18,469.9–18,550.8) | 2,113.4 (2,108.7–2,119.5) | 3,196.8 (3,196.8–3,196.8) | 714 (704–721) |
| Indonesia | Past vaccination | 224.0 (223.6–224.6) | 115.5 (115.3–115.8) | 2,540.2 (2,534.7–2,546.7) | 168.9 (168.5–169.4) | — | Reference |
|  | 14 | 13.7 (13.7–13.8) | 7.1 (7.1–7.1) | 155.8 (155.2–156.4) | 10.4 (10.4–10.5) | 20.5 (20.5–20.5) | 65 (64–65) |
|  | 18 | 117.9 (117.4–118.5) | 60.8 (60.5–61.1) | 1,336.4 (1,330.5–1,343.0) | 99.5 (99.0–100.0) | 160.7 (160.7–160.7) | 46 (45–46) |
|  | 25 | 248.8 (245.8–251.2) | 128.3 (126.8–129.6) | 2,820.4 (2,786.6–2,848.2) | 226.3 (223.4–228.6) | 451.8 (451.8–451.8) | 204 (200–211) |
|  | 30 | 288.3 (284.4–291.2) | 148.7 (146.7–150.2) | 3,266.0 (3,222.1–3,299.7) | 271.4 (267.5–274.3) | 662.5 (662.5–662.5) | 561 (550–576) |
| Kenya | Past vaccination | 61.4 (60.3–62.8) | 38.6 (37.9–39.4) | 809.1 (795.0–827.6) | 73.2 (71.8–74.9) | — | Reference |
|  | 14 | 16.3 (15.9–16.5) | 10.2 (10.0–10.3) | 214.5 (209.9–217.2) | 19.6 (19.2–19.8) | 12.8 (12.8–12.8) | -29 (-30–-27) |
|  | 18 | 36.0 (34.4–37.3) | 22.6 (21.6–23.5) | 474.0 (454.0–492.3) | 45.7 (43.6–47.6) | 34.8 (34.8–34.8) | -5 (-15–8) |
|  | 25 | 58.6 (54.6–62.9) | 36.8 (34.3–39.5) | 772.2 (719.2–828.7) | 79.9 (74.1–86.2) | 90.0 (90.0–90.0) | 123 (96–151) |
|  | 30 | 66.3 (61.6–71.3) | 41.6 (38.7–44.8) | 873.1 (811.9–940.0) | 93.7 (86.8–101.3) | 123.1 (123.1–123.1) | 260 (222–305) |
| Nigeria | Past vaccination | 94.4 (94.1–94.8) | 60.1 (59.8–60.3) | 1,002.7 (999.1–1,006.7) | 23.1 (23.0–23.2) | — | Reference |
|  | 14 | 89.6 (89.5–89.9) | 57.0 (56.9–57.2) | 951.1 (949.7–954.6) | 22.9 (22.8–22.9) | 95.3 (95.3–95.3) | 83 (82–84) |
|  | 18 | 172.9 (171.4–175.4) | 109.9 (109.0–111.5) | 1,835.0 (1,819.2–1,862.1) | 46.7 (46.2–47.4) | 214.0 (214.0–214.0) | 124 (118–128) |
|  | 25 | 240.1 (236.8–244.7) | 152.7 (150.6–155.6) | 2,549.1 (2,514.2–2,598.1) | 68.6 (67.6–70.0) | 420.9 (420.9–420.9) | 401 (387–424) |
|  | 30 | 259.7 (256.0–264.7) | 165.2 (162.8–168.3) | 2,756.3 (2,716.4–2,808.6) | 76.2 (75.0–77.8) | 540.9 (540.9–540.9) | 660 (643–687) |

Incremental cost-effectiveness ratio (ICER) estimates generally increased with each additional year of catch-up age, indicating diminishing marginal returns relative to the additional resources required (Figure 4). Female catch-up HPV vaccination up to age 30 years was cost-effective in all but one of the six modelled countries when using a cost-effectiveness threshold of 30% of GDP per capita. Catch-up in Eswatini was cost-saving up to age 30 years, i.e., savings through cervical cancer treatment costs averted were greater than vaccination costs of extending one more age, indicated as ICER < 0 in Figure 4. In Kenya and India, catch-up was cost-saving (incremental ICER < 0, Figure 4) up to ages 18 and 10 respectively. When considering ICER of the whole programme (instead of the incremental age), catch-up was cost-saving even up to ages 22 and 19 (Figure S10). In Colombia and Indonesia, catch-up vaccination was cost-effective across all evaluated age ranges up to age 30 years. In Nigeria (the country with the lowest GDP per capita among the six analysed), catch-up was cost-effective up to age 23 years.

**Figure 4.**
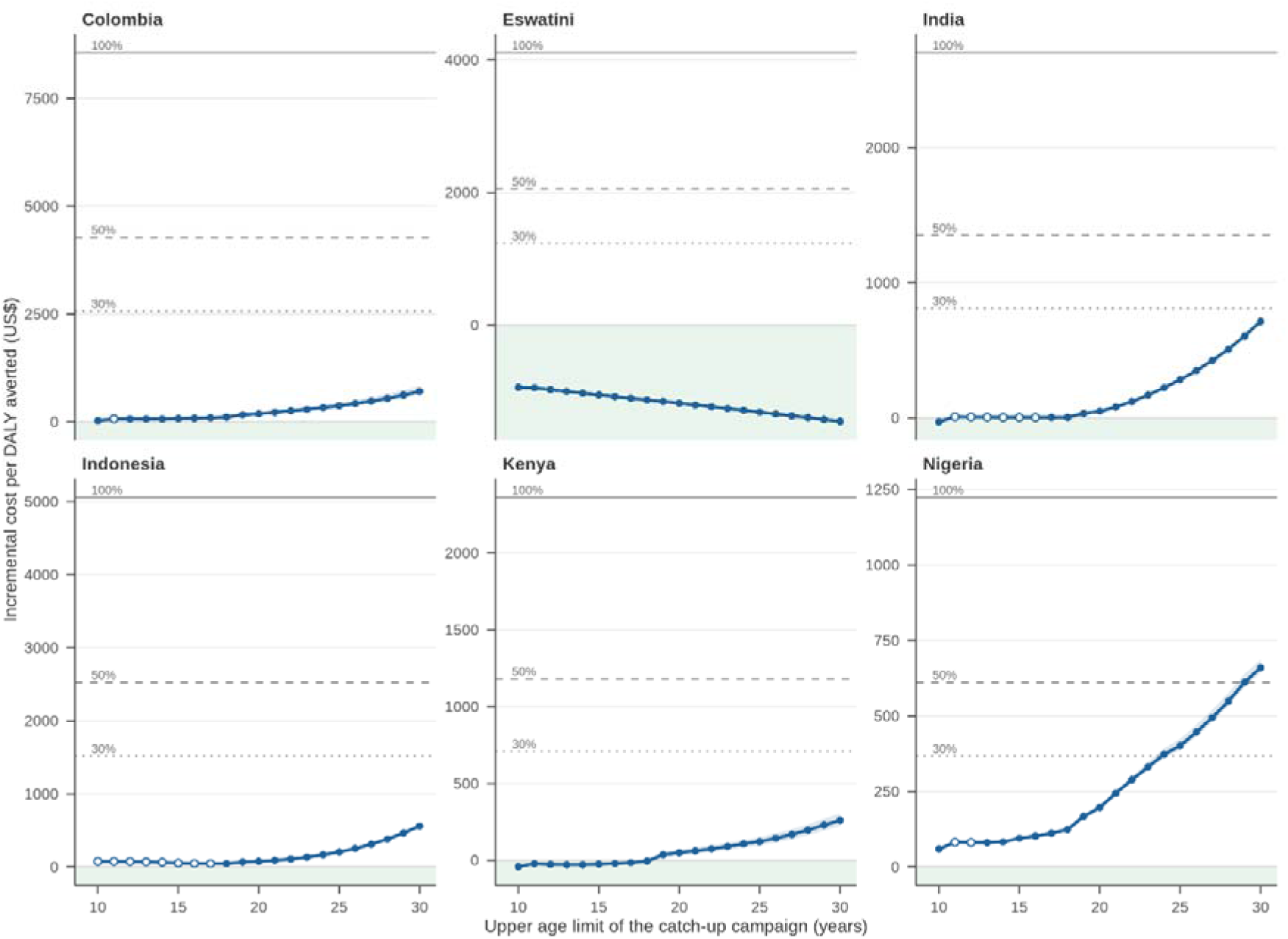
Incremental cost-effectiveness ratio (ICER) of female HPV catch-up vaccination assuming a vaccine targeting HPV16/18 and 90% coverage, and accounting for past vaccination. ICER estimates with 95%UI (in shaded area) are reported for each incremental catch-up age. Health impacts are calculated using a 0% discount rate, while treatment costs saved are discounted at 3%. The horizontal lines represent different cost-effectiveness thresholds at 30%, 50% and 100% of GDP per capita of each country. Solid and hollow dots represent being *on* and *not on* the efficiency frontier, respectively. The shaded region is where the campaign is cost-saving, that is, i.e., savings through cervical cancer treatment costs averted were greater than additional vaccination costs. Decreasing curves were due to discounting of high treatment costs. Filled circles indicate age limits on the efficiency frontier. Open circles indicate age limits that may still be affordable in itself, but where some older age limits had lower ICER.

In a series of sensitivity analyses, we assessed the impact of varying parameters. When applying 1× GDP per capita as cost-effectiveness threshold, catch-up vaccination remained cost-effective up to age 30 years for all countries (Figure 4). The results were the most sensitivity to the use of a 3% discount rate for health outcomes (not discounted in the main analysis), which decreased the maximum cost-effective catch-up age in Nigeria, India, Indonesia, and Kenya to 10, 24, 29, and 29, respectively (Table 2, S6). The results for Nigeria varied most due to its relatively low GDP per capita. The cost parameters (treatment costs, vaccine costs, delivery costs), requiring two doses for age 21 or above, and assuming waning of single-dose vaccine protection all had little effect on the maximum cost-effective age (Table 2, Figures S7). Interestingly, catch-up at lower coverages improved the cost-effectiveness slightly, due to proportionally greater herd effect (Figure S8). Lastly, in Kenya and Nigeria (the two of the six cost-effectiveness countries in an active Gavi co-financing phase), reducing the vaccination cost component in the ICER computation due to Gavi contribution (hence, meaning lower vaccination costs) slightly improves the cost-effectiveness of catch-up (Figure S9).

**Table 2:**
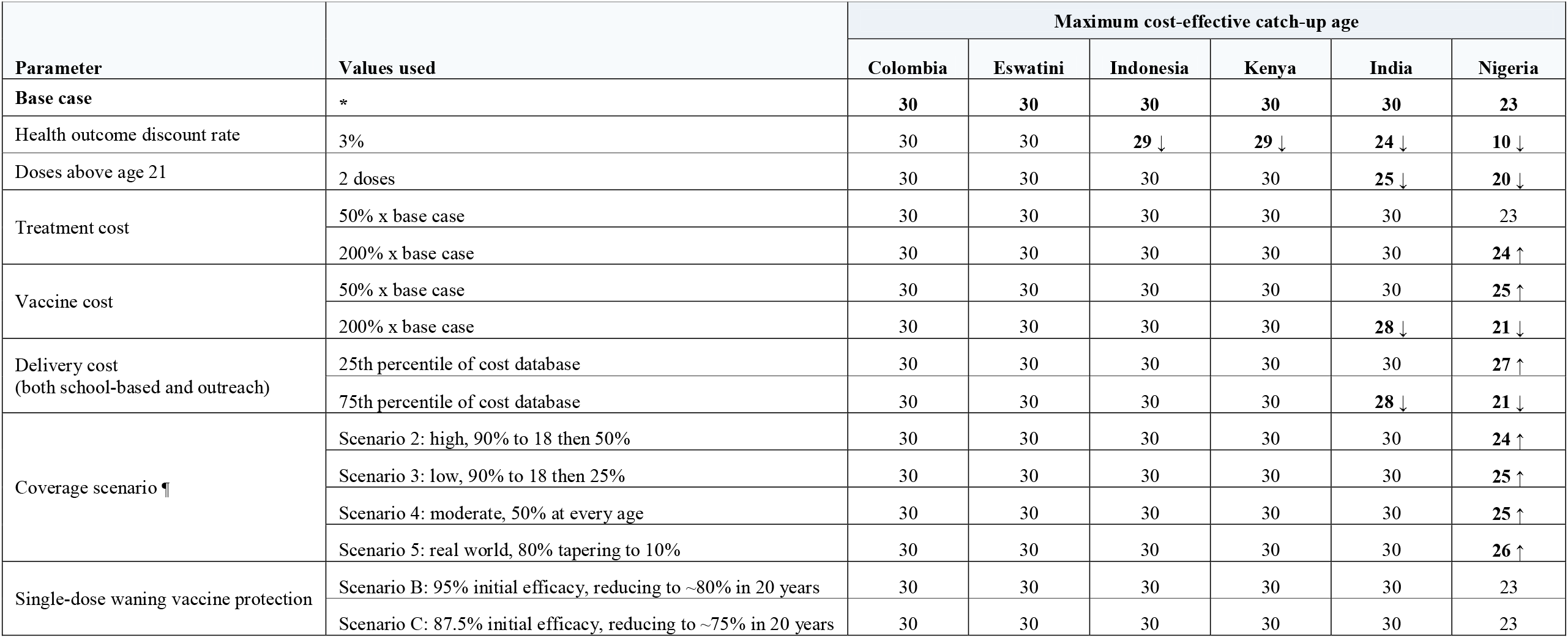
Sensitivity analyses on the impact of key input parameter variations on the maximum cost-effective age for HPV catch-up vaccination. * Values used in the base case: 0% discount rate for health outcomes, single-dose schedule for all ages, 90% coverage (scenario 1), 95% vaccine efficacy without waning (scenario A), cost estimates as described in the methods and Table S5. ¶ Figure S12.

| Parameter | Values used | Maximum cost-effective catch-up age |  |  |  |  |  |
| --- | --- | --- | --- | --- | --- | --- | --- |
|  |  | Colombia | Eswatini | Indonesia | Kenya | India | Nigeria |
| Base case | * | 30 | 30 | 30 | 30 | 30 | 23 |
| Health outcome discount rate | 3% | 30 | 30 | 29 ↓ | 29 ↓ | 24 ↓ | 10 ↓ |
| Doses above age 21 | 2 doses | 30 | 30 | 30 | 30 | 25 ↓ | 20 ↓ |
| Treatment cost | 50% x base case | 30 | 30 | 30 | 30 | 30 | 23 |
|  | 200% x base case | 30 | 30 | 30 | 30 | 30 | 24 ↑ |
| Vaccine cost | 50% x base case | 30 | 30 | 30 | 30 | 30 | 25 ↑ |
|  | 200% x base case | 30 | 30 | 30 | 30 | 28 ↓ | 21 ↓ |
| Delivery cost<br>(both school-based and outreach) | 25th percentile of cost database | 30 | 30 | 30 | 30 | 30 | 27 ↑ |
|  | 75th percentile of cost database | 30 | 30 | 30 | 30 | 28 ↓ | 21 ↓ |
| Coverage scenario ¶ | Scenario 2: high, 90% to 18 then 50% | 30 | 30 | 30 | 30 | 30 | 24 ↑ |
|  | Scenario 3: low, 90% to 18 then 25% | 30 | 30 | 30 | 30 | 30 | 25 ↑ |
|  | Scenario 4: moderate, 50% at every age | 30 | 30 | 30 | 30 | 30 | 25 ↑ |
|  | Scenario 5: real world, 80% tapering to 10% | 30 | 30 | 30 | 30 | 30 | 26 ↑ |
| Single-dose waning vaccine protection | Scenario B: 95% initial efficacy, reducing to ~80% in 20 years | 30 | 30 | 30 | 30 | 30 | 23 |
|  | Scenario C: 87.5% initial efficacy, reducing to ~75% in 20 years | 30 | 30 | 30 | 30 | 30 | 23 |

Overall ICER estimates, meaning ICER estimates comparing the impact of the entire catch-up programme to routine vaccination only (instead of by 1-year incremental age step) are reported in Figure S10. These estimates, while not used here to determine the maximum age of catch-up, are used for comparison to other public health interventions.

## Discussion

Catch-up vaccination offers a critical opportunity to reach individuals who previously missed HPV vaccination, thereby accelerating cervical cancer elimination. In this study, we showed that extending female HPV vaccination beyond the routine age range, potentially up to age 30 years, would be impactful and economically attractive across LMICs with diverse epidemiological patterns and economic conditions.

Our findings indicate that, in addition to the past vaccination, a one-time catch-up HPV vaccination for girls and young women aged 9-30 years in 2026, requiring 951 million doses and 11,349 million US$, can reduce the cervical cancer burden by more than 6.93 million cases and 3.56 million deaths in the 132 LMICs modelled. In our analysis we considered both absolute and per capita effects to guide equitable and effective global and local health resource allocation. Large countries like India would have substantial absolute reductions despite moderate incidence, while smaller nations like Eswatini would have significant relative benefits due to elevated incidence rates.

One of the key findings of our modelling study is that catch-up HPV vaccination for females up to age 30 years (the highest catch-up age considered in the analyses) is cost-effective for all six selected countries with different epidemiological settings and despite past vaccination coverages, in most modelled settings. Excellent value for money was observed in all six countries studied even under conservative thresholds (0.3–1× GDP per capita), rather than the 3× GDP per capita sometimes used.^17^ In some cases, catch-up vaccination may even be cost saving, particularly where cervical cancer treatment is costly. For example, in Eswatini, treatment expenses are the highest among the countries analysed, primarily because invasive cervical cancer care must be accessed outside the country, typically in private hospitals in South Africa.^18^ The value of catch-up vaccination would be even greater if broader societal benefits were considered in our analysis—such as prevention of other HPV-related diseases, herd effects in men, productivity gains, emotional well-being, the long-term welfare of children who might otherwise lose their mothers, reduced household financial toxicity, and enhanced equity. We expect these results to hold for all countries in the middle-income category, as a lower maximum cost-effective catch-up age was only found for Nigeria (a country at the lower range of GDP per capita in the middle-income category), and only under some modelled conditions (e.g., 3% discounting for health outcomes and two doses for age 21 or above). For low-income countries, a lower maximum cost-effective catch-up age could be expected when based on GDP/capita thresholds.

Affordability of HPV catch-up vaccination was contextualized in our study by comparing estimated programme costs to each country’s CHE and immunization budget, given the absence of a universal benchmark. This approach offers a practical reference point for policymakers to assess the relative scale of investment required. For example, as a share of five-year CHE, the median budget impact was 0.17% (IQR: 0.08%–0.40%) for catch-up to age 30 years. In comparison, spending on HIV/AIDS and other sexually transmitted diseases represented about 9% of annual mean CHE, based on LMIC data from 2013–2023.^19^ Although not directly comparable, these figures suggest that HPV catch-up vaccination, which requires only a one-time investment, may be achievable within existing health financing structures.

We acknowledge the importance of adopting flexible, context-sensitive approaches to HPV vaccination policy. Rather than defining a strict upper age limit for catch-up based on one criterion, we support decision-making by providing estimates for a comprehensive set of outcomes, covering health benefits, cost-effectiveness, as well as budget constraints. Cost-effectiveness and budget impact analyses offer two complementary lenses.^20,21^ In the absence of a universal threshold for affordability, these results together suggest a pragmatic strategy—selecting the widest catch-up age range that remains both cost-effective and feasible within available resources.

Our findings are in line with previous modelling studies which have shown that female catch-up vaccination up to possibly age 30 years (especially under single dose) is an efficient strategy to accelerate cervical cancer elimination across diverse settings.^3,8,9^ They are also consistent with other studies evaluating cost-effectiveness of catch-up vaccination within individual low- and middle-income countries.^12,13^

Our analysis highlights that vaccine price is a key driver of overall budget impact, underscoring the importance of targeted financial mechanisms to reduce vaccine purchase prices. In addition, our study showed that Gavi’s HPV vaccine support through reduced pricing and partial funding of delivery, totalling an estimated 495 million US$ representing 49.0% of the 56 eligible countries’ catch-up campaign costs up to 18 years old, is crucial to the feasibility of the proposed one-time catch-up programme. In the long term, expanding pooled procurement approaches, facilitating technology transfers, and strengthening local manufacturing capacity in LMICs are ways to ensure an affordable and sustainable vaccine supply and reduce reliance on external support. Another way to improve affordability and sustainability is to integrate catch up vaccination into existing health care delivery platforms, such as reproductive health clinics and community outreach programmes. This would be a practical and cost efficient approach.^22,23^

A major strength of our study is the use of a large volume of empirical data, notably from large databases of region-specific or even country-specific data: DHS sexual behaviour^24^, HPV prevalence (systematic review^16^), GLOBOCAN cervical cancer burden ^25^, cervical cancer attributable fraction (systematic review)^26^, and cervical cancer treatment costs (systematic review)^27^. In addition, we explicitly accounted for past vaccination, a step seldom done in this type of large-scale multi-country modelling study. Finally, our study is the first of its kind to estimate the budgetary impact across 132 LMICs, offering policymakers actionable, context-specific insights into affordability and resource allocation to support effective planning and prioritization of interventions.

Our study has some limitations. First, due to general scarcity of such data, our analysis relied on median delivery cost estimates from only 22 LMICs compiled from two comprehensive systematic reviews^28,29^ and WHO internal estimates. While this may not capture the full heterogeneity across settings, sensitivity analyses exploring a much wider range demonstrated that our conclusions were robust to plausible variations in delivery costs. Second, we did not explicitly model the transmission and the interaction of HPV with HIV. We expect this to cause only limited overestimation of health impact, as our model was calibrated to the local HPV prevalence and cervical cancer incidence. Moreover, the sensitivity analyses indicate that catch-up under a two-dose schedule, with which women living with HIV should be protected, could still be cost-effective.^30^ Finally, we did not consider the use of a nonavalent vaccine, as this option will likely remain too costly in the near future given the current vaccine price and budgetary constraints in LMICs.

In conclusion, HPV catch-up vaccination up age 30 years would be an impactful and cost-effective strategy for many LMICs, and its implementation will depend on budgetary constraints and country-specific priorities. While the investment may seem considerable, the cost of inaction may be even greater if young girls and women who may still benefit from vaccination age out of eligibility and remain unprotected.

## Methods

### Model description and parametrization

We adapted the previously validated HPV transmission model (RHEA)^3^ and cervical cancer progression model (ATLAS)^31^ from the IARC/WHO METHIS platform ^15^ to estimate the health-economic impact of female catch-up HPV vaccination. RHEA simulates the dynamic transmission of high-risk HPV types in populations through sexual contact. The model incorporates sexual contact parameters that depend on sex, age, sexual activity level, and type-specific HPV natural history parameters. The model estimates reductions in cumulative HPV incidence in female birth cohorts for which catch-up vaccination was considered. ATLAS then estimates cohort-specific lifetime number of cervical cancer cases and deaths expected for different vaccination scenarios, by combining RHEA estimates on HPV incidence, HPV type-specific attributable fractions of cervical cancer, and baseline cervical cancer incidence and mortality, while taking into account the competing risk of dying from other causes.

Model parametrization for the 132 past or current LMICs was done through the following workflow. Countries were first grouped into clusters based on Demographic and Health Surveys (DHS) data ^24^, capturing archetypical sexual behaviour patterns that drive HPV transmission. For each cluster, we calibrated RHEA to available data within each cluster regarding sexual behaviour (from DHS) and HPV prevalence data (from a comprehensive literature review) (see Figure S11 for model fit). Country-specific fine-tuning was done in ATLAS by utilizing country-specific cervical cancer incidence and mortality from GLOBOCAN^25^ and country- or region-specific HPV attributable fractions from a recent systematic review.^26^ This workflow enables modelling of 132 countries by leveraging maximum amount of available data within epidemiological clusters. Full details of the clustering, model parametrization, and data used are reported in a dedicated article.^16^

Estimation of health economic impact required epidemiological and cost inputs. Health impact and budget impact were estimated for all 132 countries. For countries where immunization budgets and Current Health Expenditure (CHE) data were available (Table S4), we also estimated corresponding proportions. For the cost effectiveness analysis, high-quality primary data on cervical cancer treatment costs were necessary. Among LMICs with such data identified through our systematic review^27^, we selected one representative country per epidemiological profile (as derived in Man et al.^16^) to serve as a proof of concept: Colombia, Eswatini, India, Indonesia, Kenya, and Nigeria (Table S6).

### Vaccination simulation

We simulated the following vaccination strategies:

- No vaccination in the past and future.
- “Past vaccination”, here meaning vaccination up to 2025 according to the reported age target and observed coverage reported on WHO HPV Dashboard^1^ and assuming, from 2026 onwards, continuation of the last observed coverage.
- A series of catch-up scenarios, with settings as described in point 2, but with an additional one-time female catch-up in 2026 up to older and older ages to a maximum age of 30. In the base-case analysis, 90% coverage was assumed, which here means a top-up until 90% of each cohort is vaccinated. For birth cohorts with past coverage exceeding 90%, no catch-up was simulated.

Throughout, vaccination regardless of dose schedule was assumed to provide lifelong protection with 95% efficacy against acquisition of new infection of HPV types 16 and 18, consistent with existing clinical trial evidence^32^, with slight (∼10%) cross-protection against HPV types 31, 33, and 45, which is a conservative estimate for a wide range of available vaccine products targeting HPV 16/18.^33^ The model incorporated reduced effectiveness with increasing age, as vaccination is assumed not to clear HPV infection acquired prior to HPV vaccination.

### Cost data

#### Vaccine and vaccine delivery costs

We used vaccine procurement prices from the Market Information for Access (MI4A), vaccine purchase database (Table S4)^34^. We assumed the cost of a single-dose schedule in the base-case analysis. Age-specific delivery was modelled: ages 9–14 years through school; ages 15–18 years through school for in-school girls plus community outreach for out-of-school girls; and ages ≥19 years through community outreach. Delivery costs for school-based and community outreach strategies were compiled from two systematic reviews^28,29^ and WHO internal estimates, in total including 48 estimates spanning 22 countries. We used the median delivery cost and adjusted for expected economies of scale (Table S4). For budget impact, we estimated financial costs as one third of economic costs.^28,35^

#### Cancer treatment costs

We used cervical cancer treatment cost data identified through a literature review (Table S5).^27^ All costs were expressed in 2024 US dollars (US$). Costs not originally reported in 2024 US$ were adjusted by first applying the Consumer Price Index (CPI) inflation adjustment^36^ and then converting local currencies using 2024 US$ exchange rates.^37^

### Model Output

#### Health impact

Health impact was estimated as the number of cervical cancer cases, deaths, and DALYS averted over the lifetime of the vaccinated cohorts aged 9–30 years in 2026 across 132 countries.

#### Budget impact

We estimated the vaccination budget in US$ and in proportion of countries’ five-year immunization budget and CHE (Table S5). We chose a five-year timeframe because the intervention may realistically take multiple years to implement. For Gavi eligible countries, the co-financing portion was included in the total country budget. In a sub-analysis, we estimated the proportion of the budget supported by Gavi.

#### Cost-effectiveness

We calculated the ICER for each vaccination strategy by progressively increasing the upper age limit of the catch-up vaccination cohort by one-year increments. For each increment, we compared the additional cost and disability-adjusted life years (DALYs) averted to the previous age. DALYs were calculated by combining estimates of years of life lost (YLL) with disability weights for cervical cancer.^38^ The maximum age at which catch-up vaccination remained cost-effective was identified using a willingess-to-pay threshold of less than 30% of the country-specific GDP per capita.^39^ We conducted the analysis from a healthcare sector perspective. Costs were discounted at a rate of 3% and DALYs were not discounted, in accordance with WHO recommendations.^40^

#### Sensitivity analysis

We conducted the following sensitivity analyses: four less ideal catch-up vaccination coverage scenarios (Figure S12); alternative discount rates for costs and/or health outcomes; a cost-effectiveness threshold of 100% of country-specific GDP per capita; higher or lower vaccine delivery costs and vaccine price; higher or lower treatment costs (from 50% to 200%); a two-dose schedule for ages 21 years or above; waning of single-dose long-term vaccine protection, with scenarios defined in our previous work (scenario B: waning to ∼80% efficacy 20 years postvaccination; scenario C: lower initial efficacies at approximately 87.5%, and waning to ∼75% efficacy 20 years postvaccination) ^3^; and inclusion or exclusion of the Gavi-supported share of the costs in the cost-effectiveness analysis.

Model outcomes were summarized across the 100 best-fitting parameter sets obtained by model calibration, reporting medians and 95% uncertainty intervals (2.5^th^–97.5^th^ percentiles). This study adheres to HPV-FRAME, a quality framework for modelled evaluations of HPV-related cancer control (Table S6).^41^

## Data Availability

All data produced in the present study are available upon reasonable request to the authors

## Contributors

IM and IB conceived and contributed to the study design and obtained funding for the study. IM supervised the overall project. AW, RW, AF extracted data to inform model parameters. AW, DG, and AM did the model analysis and visualisation of the study findings. IM and AW drafted the article. AG contributed to interpretation of results. All authors critically revised the manuscript for scientific content and approved the final version of the article.

## Acknowledgements

We are grateful to Paul Bloem and Karene Yeung (Department of Immunization, Vaccines and Biologicals, WHO) for providing HPV vaccination programmatic cost data used in this study. We also thank Dr Swaminathan Rajaraman and Dr Rama Ranganathan for sharing stage-specific cancer case distribution data from the Cancer Institute (WIA), a tertiary cancer care hospital in India. Additionally, we thank Dr Cebisile for clarifying Eswatini’s treatment cost data. We thank Nadia Akel (IARC/WHO) for her language editing and technical support.

## Disclaimer

Where authors are identified as personnel of the International Agency for Research on Cancer/World Health Organization, the authors alone are responsible for the views expressed in this article and they do not necessarily represent the decisions, policy or views of the International Agency for Research on Cancer / World Health Organization.

## References

1. World Health Organization. HPV Dashboard. https://www.who.int/teams/immunization-vaccines-and-biologicals/diseases/human-papillomavirus-vaccines-(HPV)/hpv-clearing-house/hpv-dashboard (accessed July 2026).

2. Dillner J, Elfström KM, Baussano I. Prospects for accelerated elimination of cervical cancer. Prev Med 2021; 153.

3. Man I, Georges D, Basu P, Baussano I. Leveraging single-dose human papillomavirus vaccination dose-efficiency to attain cervical cancer elimination in resource-constrained settings. J Natl Cancer Inst Monogr 2024; 2024(67): 400–9.

4. Arroyo Mühr LS, Gini A, Yilmaz E, et al. Concomitant human papillomavirus (HPV) vaccination and screening for elimination of HPV and cervical cancer. Nat Commun 2024; 15(1): 3679.

5. Baussano I, Sayinzoga F, Tshomo U, et al. Impact of Human Papillomavirus Vaccination, Rwanda and Bhutan. Emerg Infect Dis 2021; 27(1): 1–9.

6. World Health Organization. Human papillomavirus vaccines: WHO position paper (2022 update). Weekly epidemiological record 2022; 97(50): 645–72.

7. Schuind AE, Balaji KA, Du A, Yuan Y, Dull P. Human papillomavirus prophylactic vaccines: update on new vaccine development and implications for single-dose policy. J Natl Cancer Inst Monogr 2024; 2024(67): 410–6.

8. Drolet M, Laprise JF, Martin D, et al. Optimal human papillomavirus vaccination strategies to prevent cervical cancer in low-income and middle-income countries in the context of limited resources: a mathematical modelling analysis. Lancet Infect Dis 2021; 21(11): 1598–610.

9. Bénard É, Drolet M, Gingras G, et al. Ranking the most efficient human papillomavirus vaccination strategies in low-income and lower-middle income countries: a mathematical modelling analysis. Lancet Glob Health 2025; 13(12): e2153–e64.

10. Abbas KM, van Zandvoort K, Brisson M, Jit M. Effects of updated demography, disability weights, and cervical cancer burden on estimates of human papillomavirus vaccination impact at the global, regional, and national levels: a PRIME modelling study. Lancet Glob Health 2020; 8(4): e536–e44.

11. Gaythorpe KAM, Li X, Shankar M, et al. Quantifying relative health impact across Gavi, the Vaccine Alliance’s portfolio in 117 countries at the subregional level: a modelling study. Lancet 2026; 407(10542): 1941–52.

12. M De Carvalho T, Man I, Georges D, et al. Health and economic effects of introducing single-dose or two-dose human papillomavirus vaccination in India. BMJ Global Health 2023; 8(11): e012580.

13. Umutesi G, Hathaway CL, Heitner J, et al. The Potential Impact of a Single-Dose HPV Vaccination Schedule on Cervical Cancer Outcomes in Kenya: A Mathematical Modelling and Health Economic Analysis. Vaccines (Basel) 2024; 12(11).

14. Help save 2 million lives: close the vaccine funding gap. Nature 2025; 643(8073): 880.

15. IARC Public Health Decision Science Team METHIS modelling platform. https://iarc-miarc.gitlab.io/methis/methis.website/.

16. Man I, Macacu A, Eynard M, et al. A unified modeling platform for informing cervical cancer prevention policy decisions in 132 low- and middle-income countries (DOI: 10.64898/2026.03.18.26348700). medRxiv 2026.

17. Kazibwe J, Gheorghe A, Wilson D, Ruiz F, Chalkidou K, Chi YL. The Use of Cost-Effectiveness Thresholds for Evaluating Health Interventions in Low- and Middle-Income Countries From 2015 to 2020: A Review. Value Health 2022; 25(3): 385–9.

18. Ngcamphalala C, Ostensson E, Ginindza TG. The economic burden of cervical cancer in Eswatini: Societal perspective. Plos One 2021; 16(4): e0250113.

19. World Health Organization. Global Health Expenditure Database. https://apps.who.int/nha/database/Select/Indicators/en (accessed May 2025).

20. Bilinski A, Neumann P, Cohen J, Thorat T, McDaniel K, Salomon JA. When cost-effective interventions are unaffordable: Integrating cost-effectiveness and budget impact in priority setting for global health programs. PLoS Med 2017; 14(10): e1002397.

21. Sullivan SD, Mauskopf JA, Augustovski F, et al. Budget impact analysis-principles of good practice: report of the ISPOR 2012 Budget Impact Analysis Good Practice II Task Force. Value Health 2014; 17(1): 5–14.

22. Ndiaye C, Kyesi F, Masupha T, et al. Integrating HPV vaccine service delivery with adolescent health programmes - Experiences and perspectives from selected countries in Africa. Vaccine 2024; 42(18): S45–S8.

23. Morgan C, Giattas MR, Holroyd T, et al. Integration of other services with human papillomavirus vaccination; lessons from earlier in the life course highlight the need for new policy and implementation evidence. Vaccine 2022; 40: A94–A9.

24. The DHS Program. Demographic and Health Surveys. https://dhsprogram.com/ (accessed June 2025).

25. Ervik M, Laversanne M, Lam F, et al. Global Cancer Observatory: Cancer Today (2024 estimates, version 1.0). Lyon, France: International Agency for Research on Cancer. 2026. https://gco.iarc.who.int/today,10.3322/caac.21834 (accessed 2026 June).

26. Wei F, Georges D, Man I, Baussano I, Clifford GM. Causal attribution of human papillomavirus genotypes to invasive cervical cancer worldwide: a systematic analysis of the global literature. Lancet 2024; 404(10451): 435–44.

27. Fuady A, Setiawan D, Man I, de Kok I, Baussano I. Toward a Framework to Assess the Financial and Economic Burden of Cervical Cancer in Low- and Middle-Income Countries: A Systematic Review. JCO Glob Oncol 2024; 10: e2400066.

28. Slavkovsky R, Callen E, Pecenka C, Mvundura M. Costs of human papillomavirus vaccine delivery in low- and middle-income countries: A systematic review. Vaccine 2024; 42(6): 1200–10.

29. ThinkWell. Immunization Delivery Cost Catalogue. Geneva, 2024.

30. Nuwagaba-Biribonwoha H, Lamb MR, Dlamini X, et al. Immunogenicity of 2 versus 3 dose HPV vaccination by HIV infection status in Eswatini. Vaccine 2026; 85: 128692.

31. Bonjour M, Charvat H, Franco EL, et al. Global estimates of expected and preventable cervical cancers among girls born between 2005 and 2014: a birth cohort analysis. The Lancet Public Health 2021; 6(7): e510–e21.

32. Whitworth HS, Mounier-Jack S, Choi EM, et al. Efficacy and immunogenicity of a single dose of human papillomavirus vaccine compared to multidose vaccination regimens or no vaccination: An updated systematic review of evidence from clinical trials. Vaccine-X 2024; 19.

33. Basu P, Malvi SG, Joshi S, et al. Vaccine efficacy against persistent human papillomavirus (HPV) 16/18 infection at 10 years after one, two, and three doses of quadrivalent HPV vaccine in girls in India: a multicentre, prospective, cohort study. Lancet Oncol 2021; 22(11): 1518–29.

34. World Health Organization. Market Information for Access (MI4A) vaccine purchase database. https://www.who.int/teams/immunization-vaccines-and-biologicals/vaccine-access/mi4a/mi4a-vaccine-purchase-data (accessed May 2025).

35. Mvundura M, Slavkovsky R, Debellut F, et al. Cost and operational context for national human papillomavirus (HPV) vaccine delivery in six low- and middle-income countries. Vaccine 2023; 41(49): 7435–43.

36. World Bank Group. Consumer price index (2010 = 100). https://data.worldbank.org/indicator/FP.CPI.TOTL (accessed May 2025).

37. World Bank Group. Official exchange rate (LCU per US$, period average). https://data.worldbank.org/indicator/PA.NUS.FCRF (accessed May 2025).

38. GBD 2021 Diseases and Injuries Collaborators. Global incidence, prevalence, years lived with disability (YLDs), disability-adjusted life-years (DALYs), and healthy life expectancy (HALE) for 371 diseases and injuries in 204 countries and territories and 811 subnational locations, 1990-2021: a systematic analysis for the Global Burden of Disease Study 2021. Lancet 2024; 403(10440): 2133–61.

39. Jit M. Informing Global Cost-Effectiveness Thresholds Using Country Investment Decisions: Human Papillomavirus Vaccine Introductions in 2006-2018. Value Health 2021; 24(1): 61–6.

40. World Health Organization. WHO guide for standardization of economic evaluations of immunization programmes. 2nd ed: World Health Organization; 2019.

41. Canfell K, Kim JJ, Kulasingam S, et al. HPV-FRAME: A consensus statement and quality framework for modelled evaluations of HPV-related cancer control. Papillomavirus Res 2019; 8: 100184.

