## Supplementary appendix for "Determining context-specific economically feasible age ranges for female HPV catch-up vaccination in LMICs: a model-based health economic assessment"

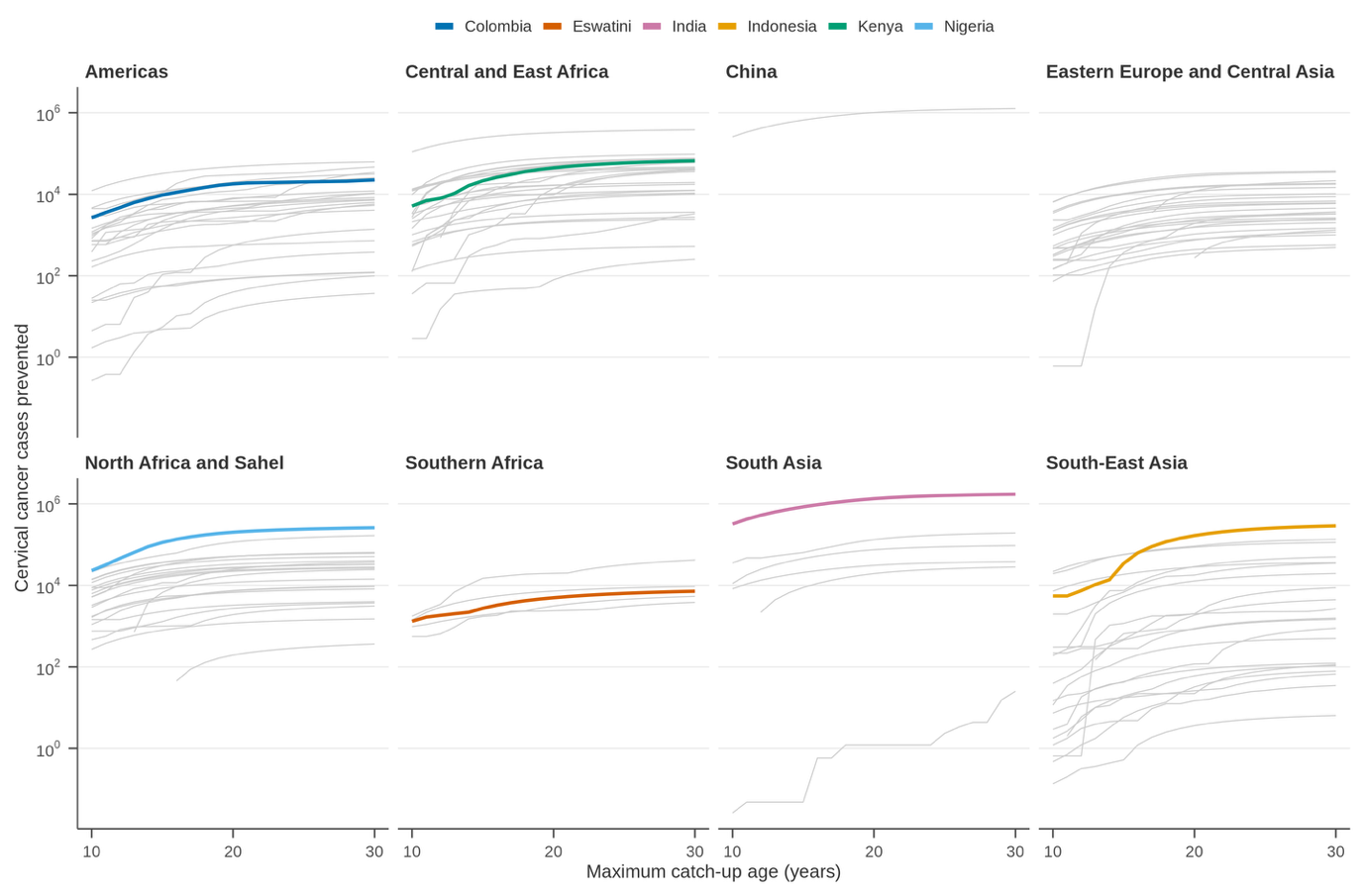

**Figure S1: Cumulative cervical cancer cases averted by one-time female catch-up HPV vaccination (with a vaccine targeting HPV16/18 and 90% coverage) up to different maximum catch-up ages in the lifetime of birth cohort aged 9–30 years in 2026 in all LMICs.** Note that it is the impact of the one-time catch-up, i.e., the additional number of cases prevention compared to the cases prevented by past vaccination. Each grey line represents a country. Each panel represents a cluster of LIMCs with similar cervical cancer epidemiological pattern as identified in Man et al.^1^ The six countries selected for the cost-effectiveness analysis are highlighted. Plateauing of curves is due to diminishing return of vaccination increasingly older women. Step-like patterns are due to past vaccination.

**African Region**

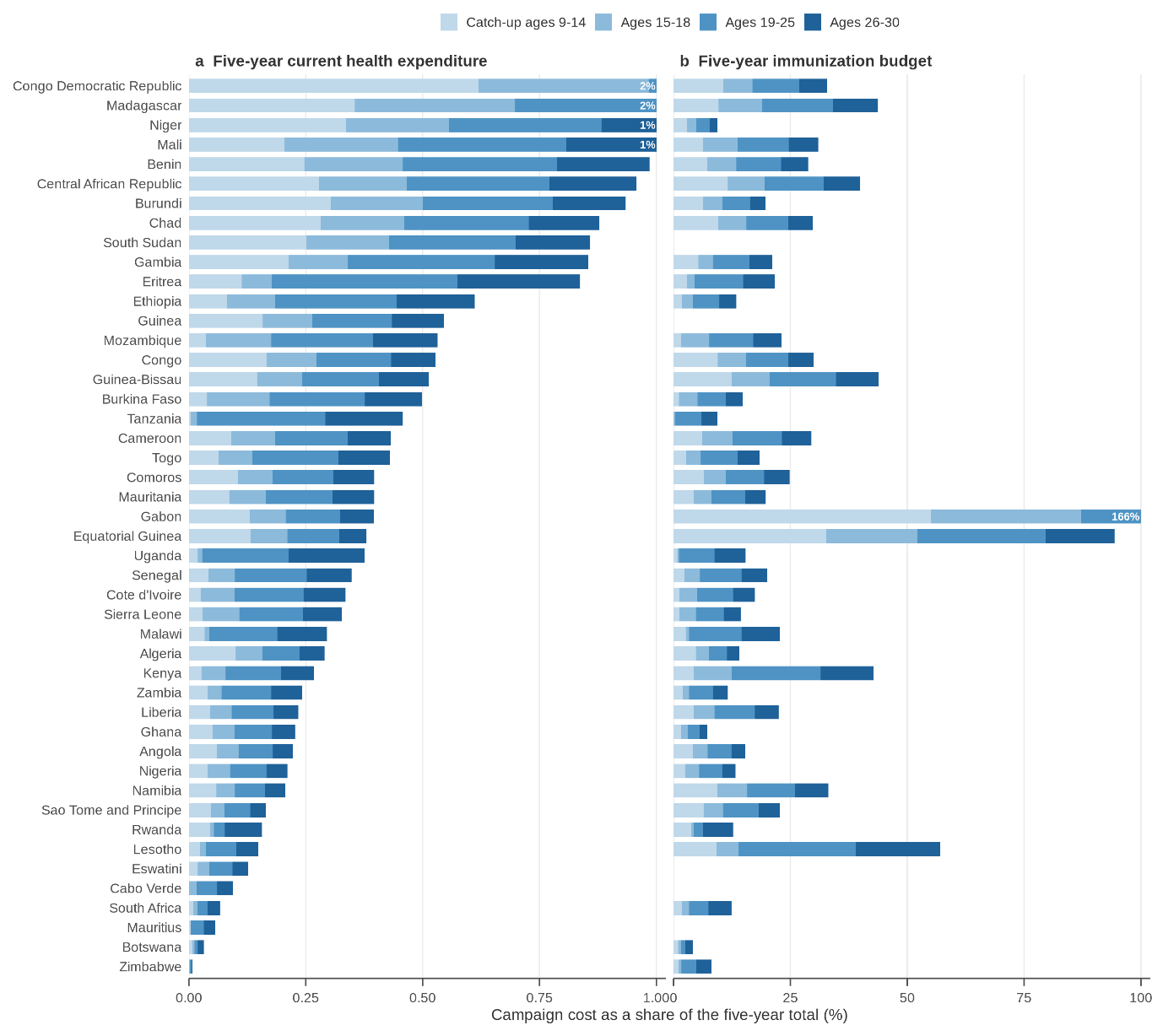

**South-East Asia Region**

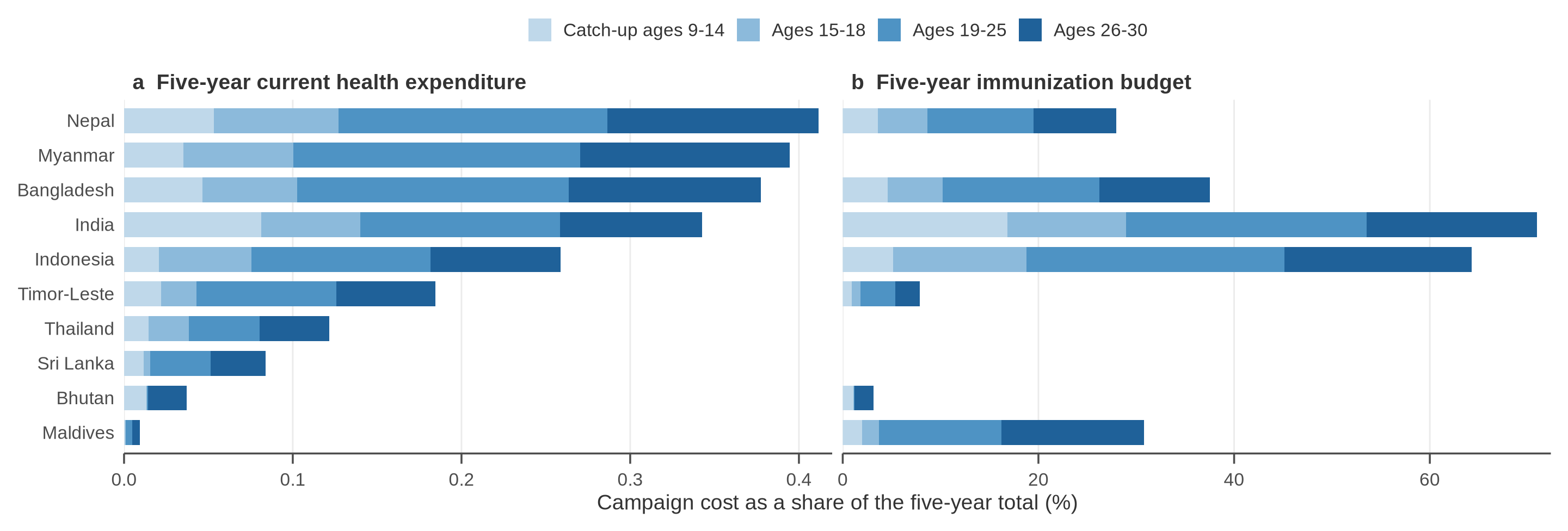

**Western Pacific Region**

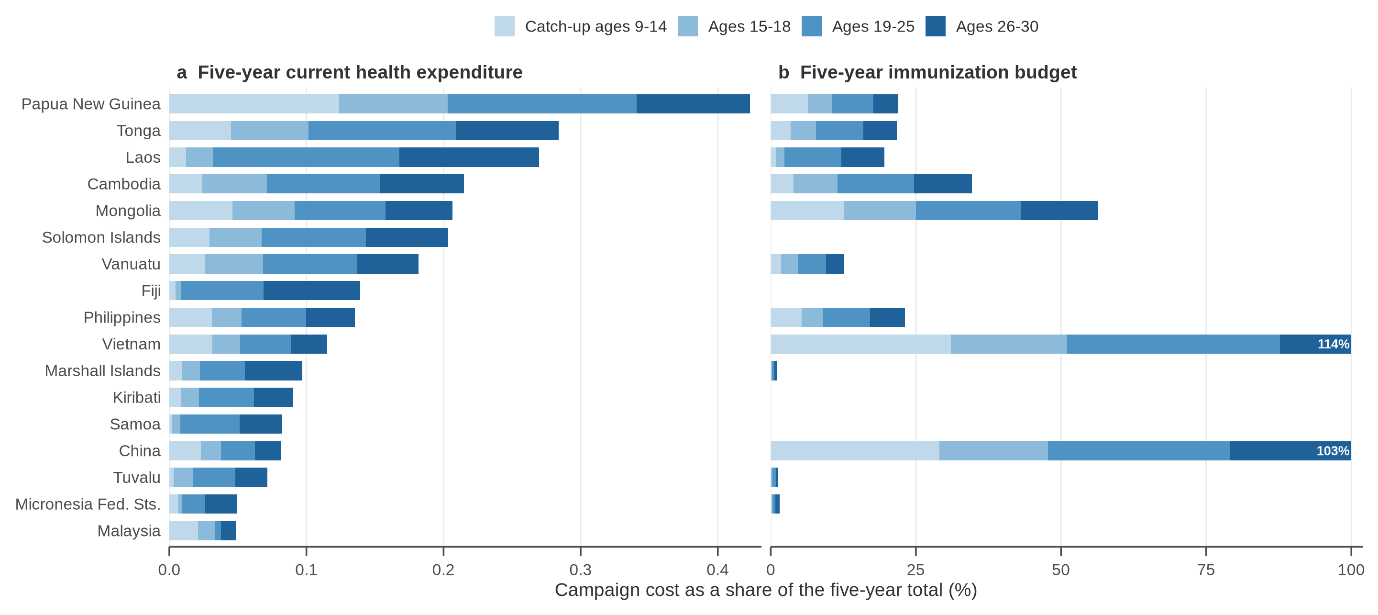

**Americas Region**

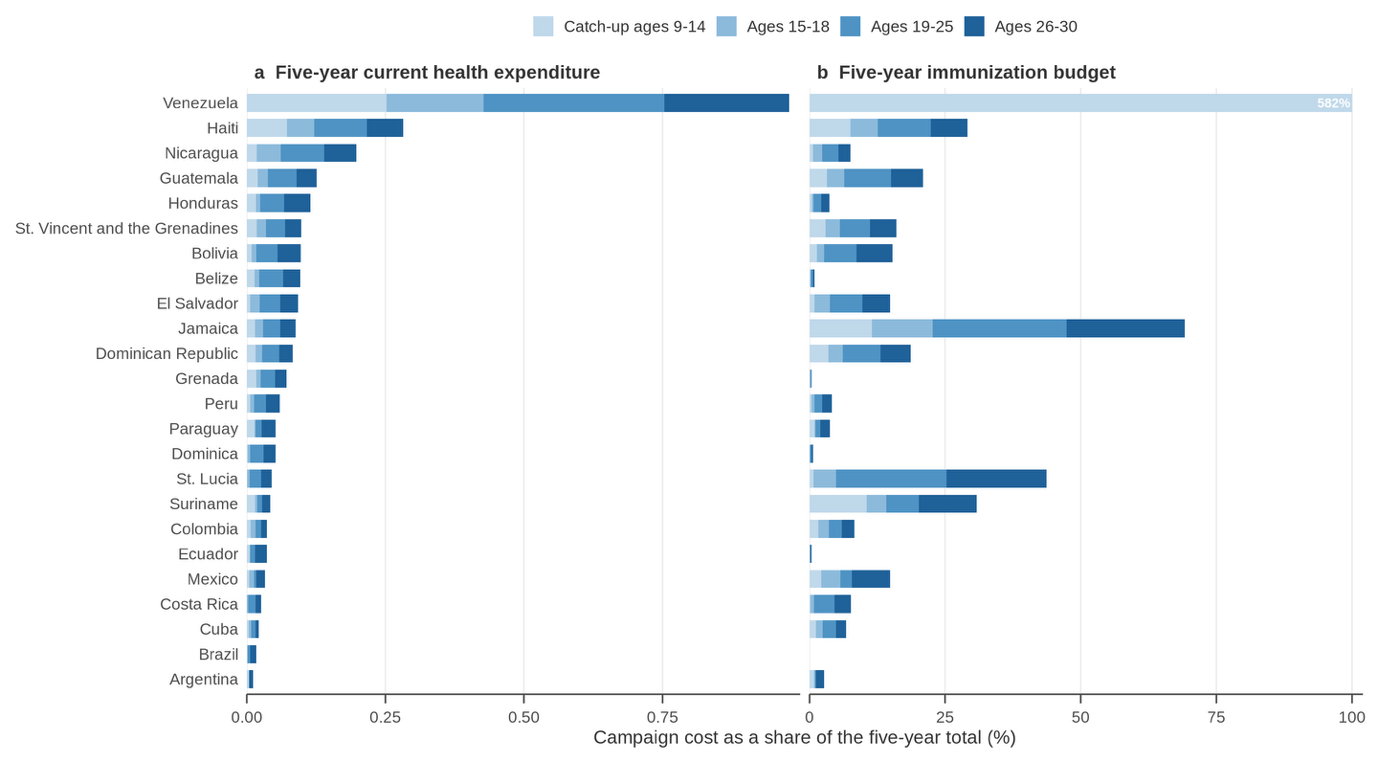

Note: Venezuela's reported immunization budget works out at US$0.88 per female over five years, which is a unit error in the source rather than a budget, so its share in the right-hand panel should not be read as a real figure.

**Eastern Mediterranean Region**

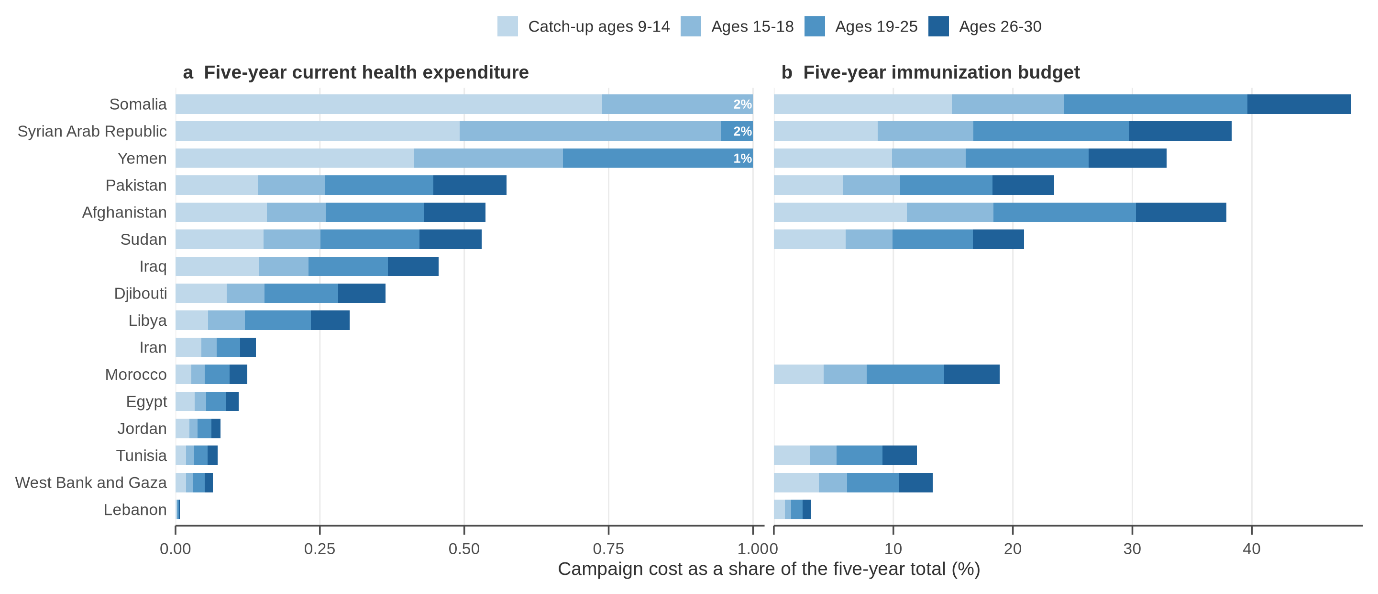

**European Region**

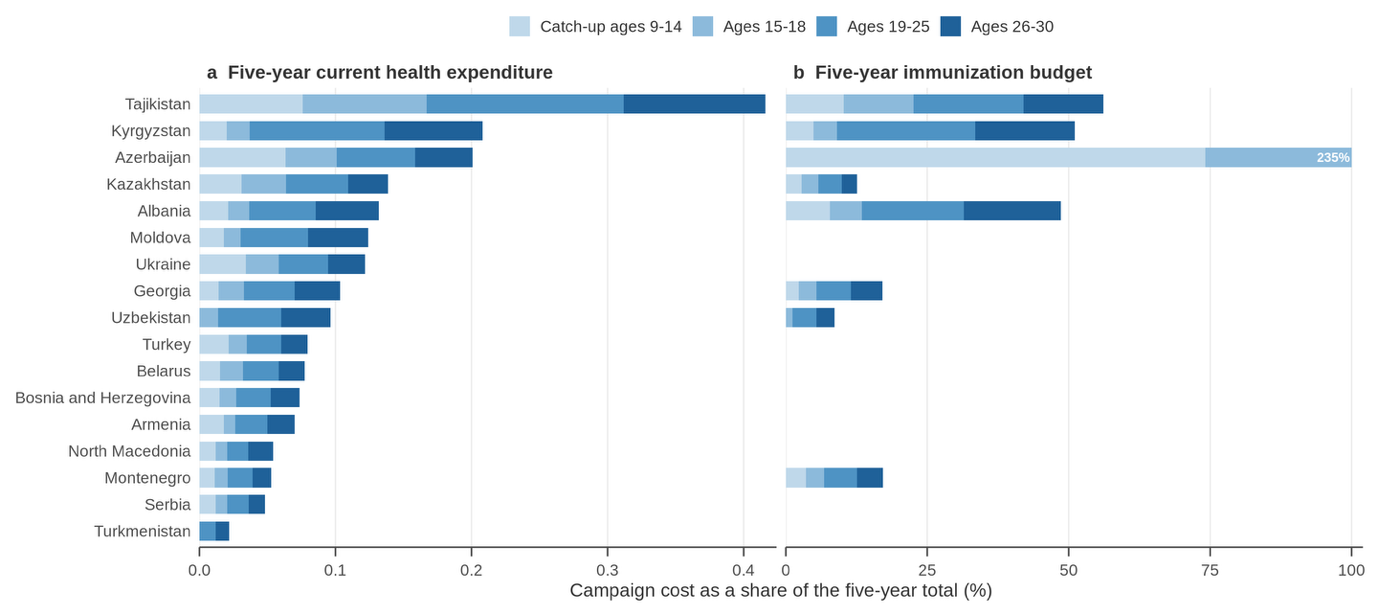

**Figure S2: Estimated cost of female catch-up HPV vaccination (assuming a vaccine targeting HPV 16/18 and 90% vaccine coverage) by age band, country, and WHO region, as percentages of five-year current health expenditure (left) and of the five-year immunization budget (right).** Both vaccine cost and delivery cost are included. Financial cost (and not economic cost) is considered. Segments are incremental: each age band is what it adds to the bands below it, so that a whole bar is a campaign reaching age 30. Countries are ordered by the left-hand panel, health expenditure, which exists for 130 of the 132 LMICs. A country with no data on current health expenditure or immunization budget has no bar in that panel rather than a bar of zero. The horizontal axis is identical on every regional panel so that regions can be compared; it is capped, and a country whose campaign costs more than the cap is drawn to the edge with its true share printed on the bar.

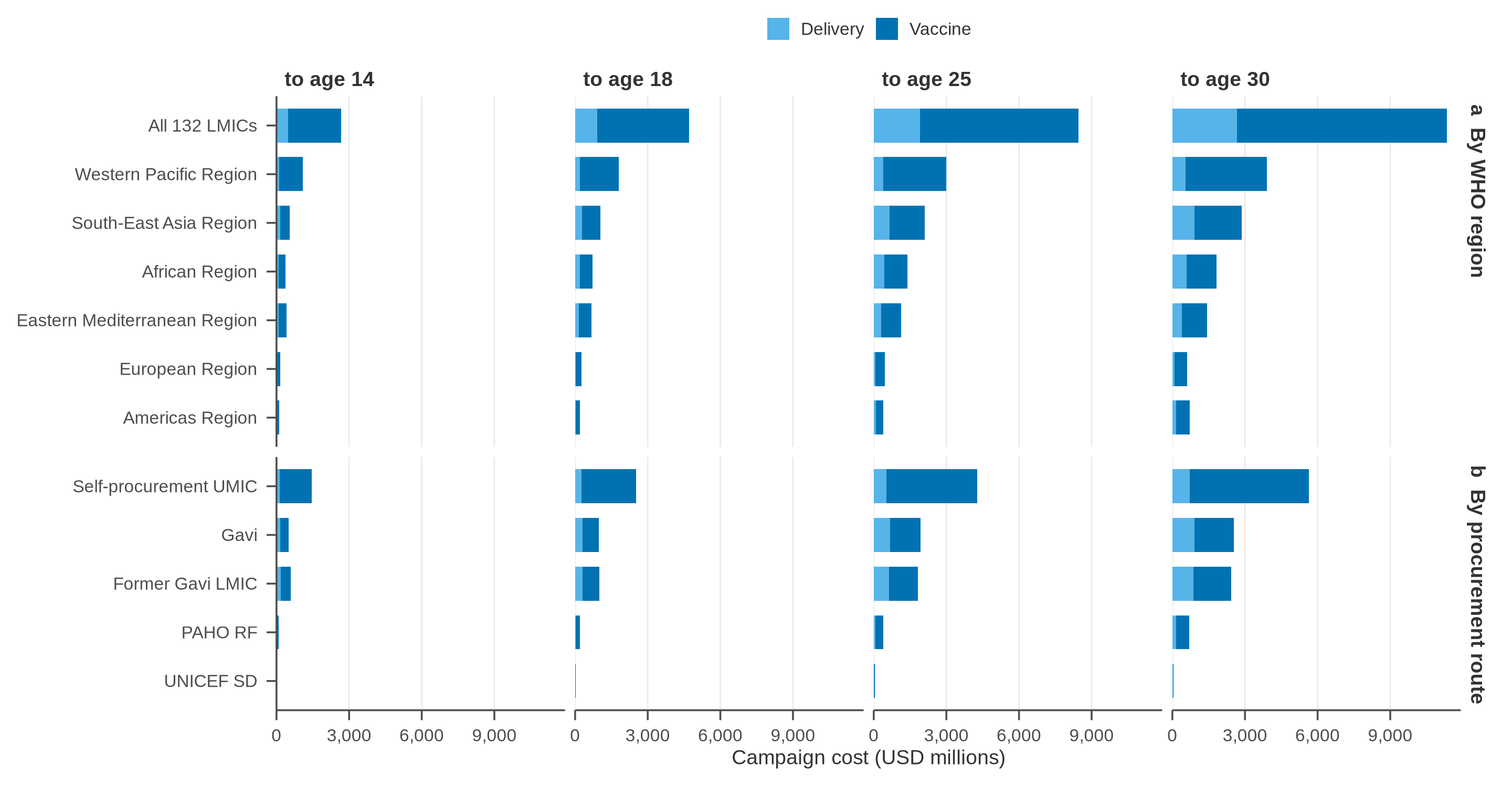

**Figure S3: Estimated vaccine delivery and vaccine purchase costs of female catch-up HPV vaccination from age 9 up to ages 14, 18, 25 and 30 years (assuming a vaccine targeting HPV 16/18 and 90% vaccine coverage) by age band, WHO region, and procurement route, and for all 132 LMICs.** Financial cost (and not economic cost) is considered. In general, delivery costs accounted for a smaller share of the total budget for catch-up vaccination programs than vaccine procurement costs, with a median estimate of 30.6% (IQR: 16.4%–31.8%) for maximum catch-up age of 18 and 35.5% (IQR: 20.1%–36.5%) for maximum age 30, due to higher delivery costs assumed at higher age of vaccination.

**African Region**

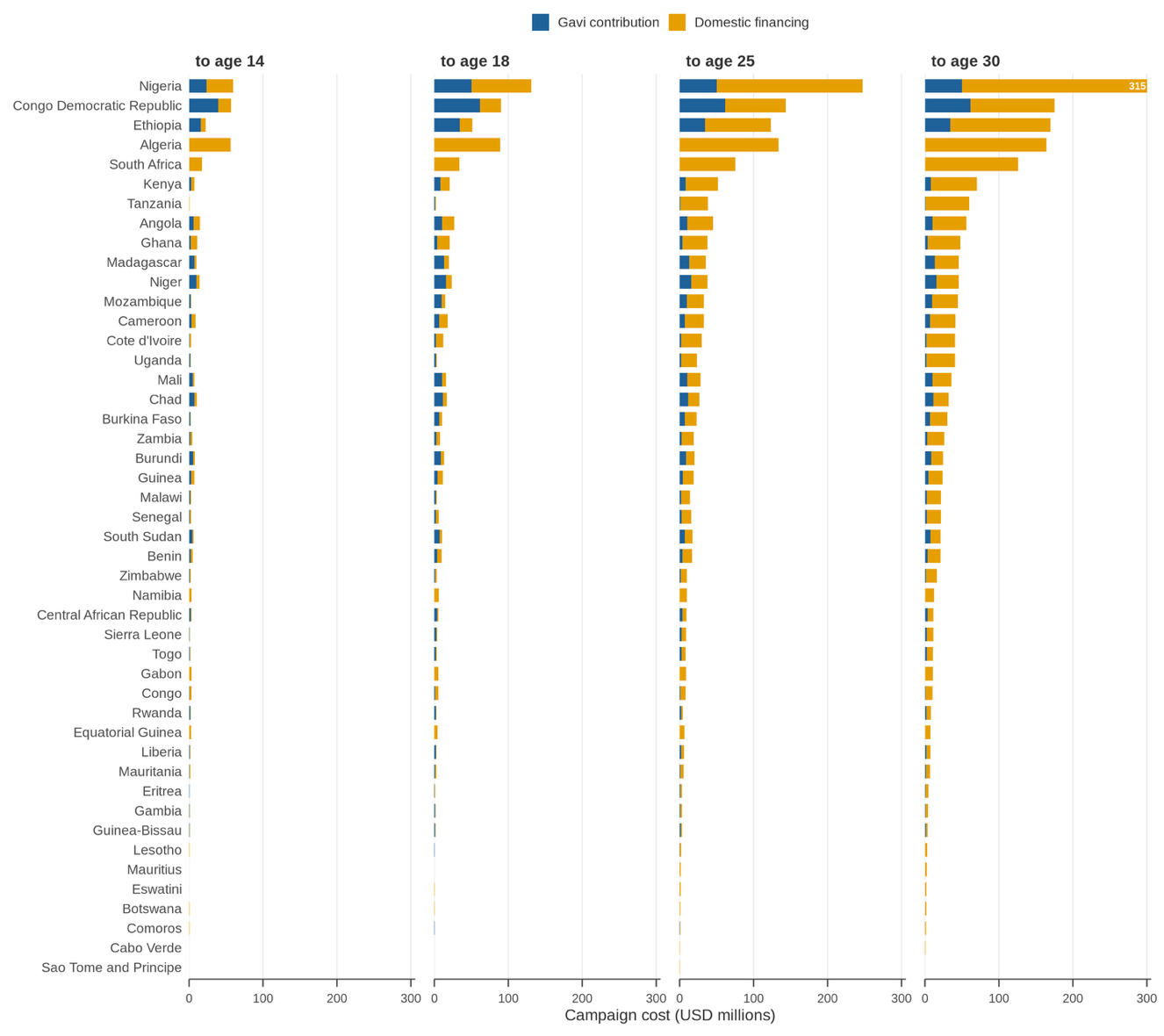

**South-East Asia Region**
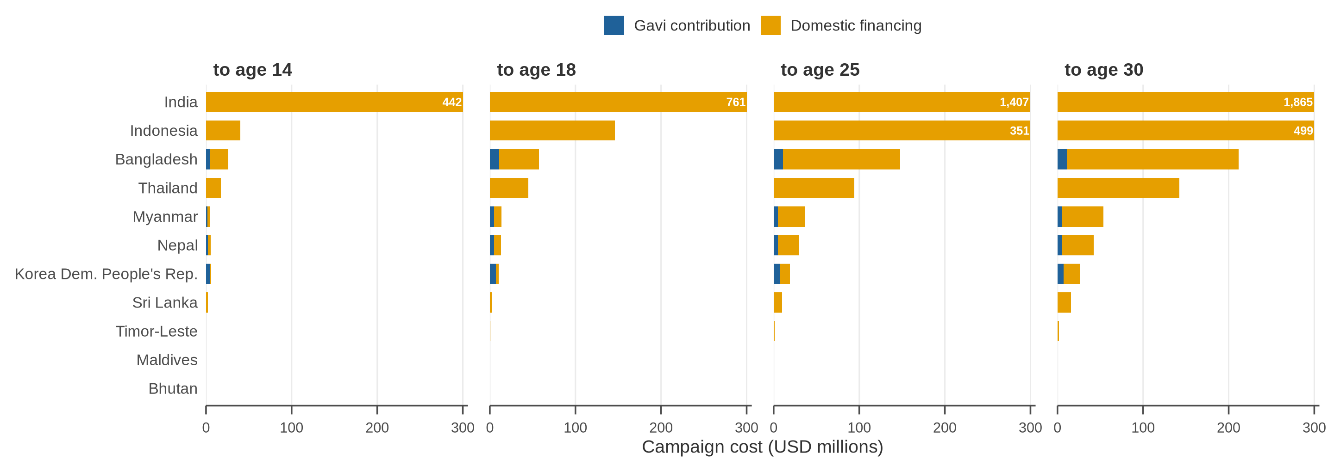

**Western Pacific Region**

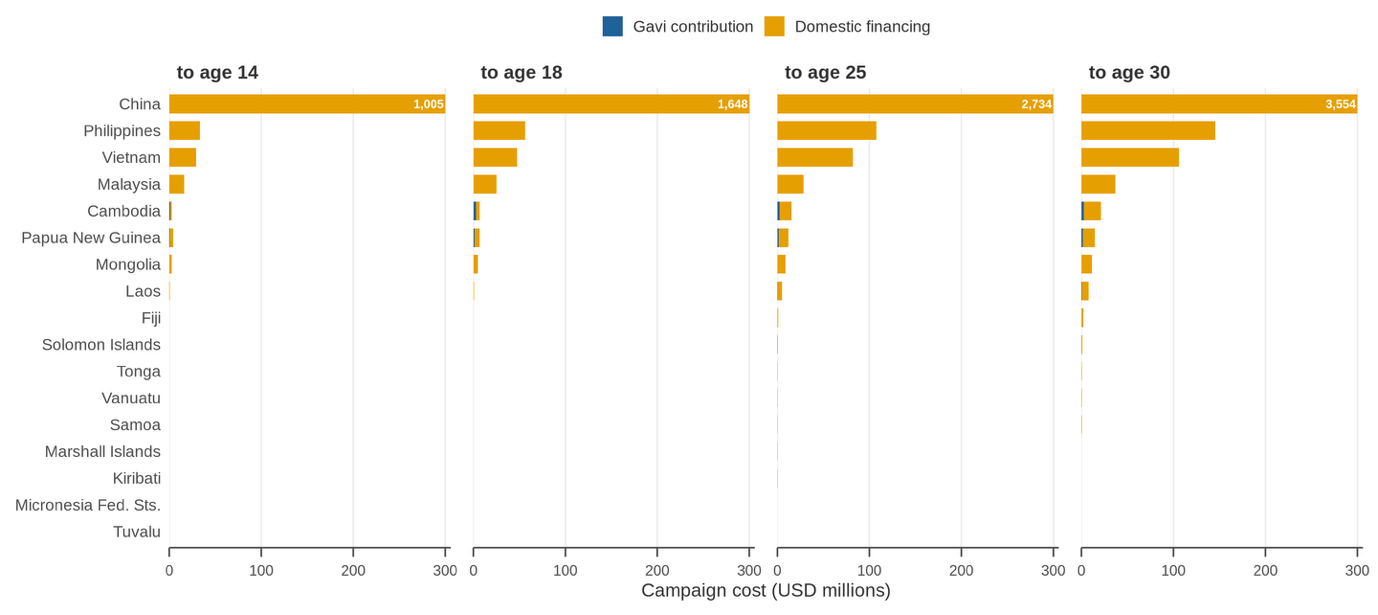

**Americas Region**
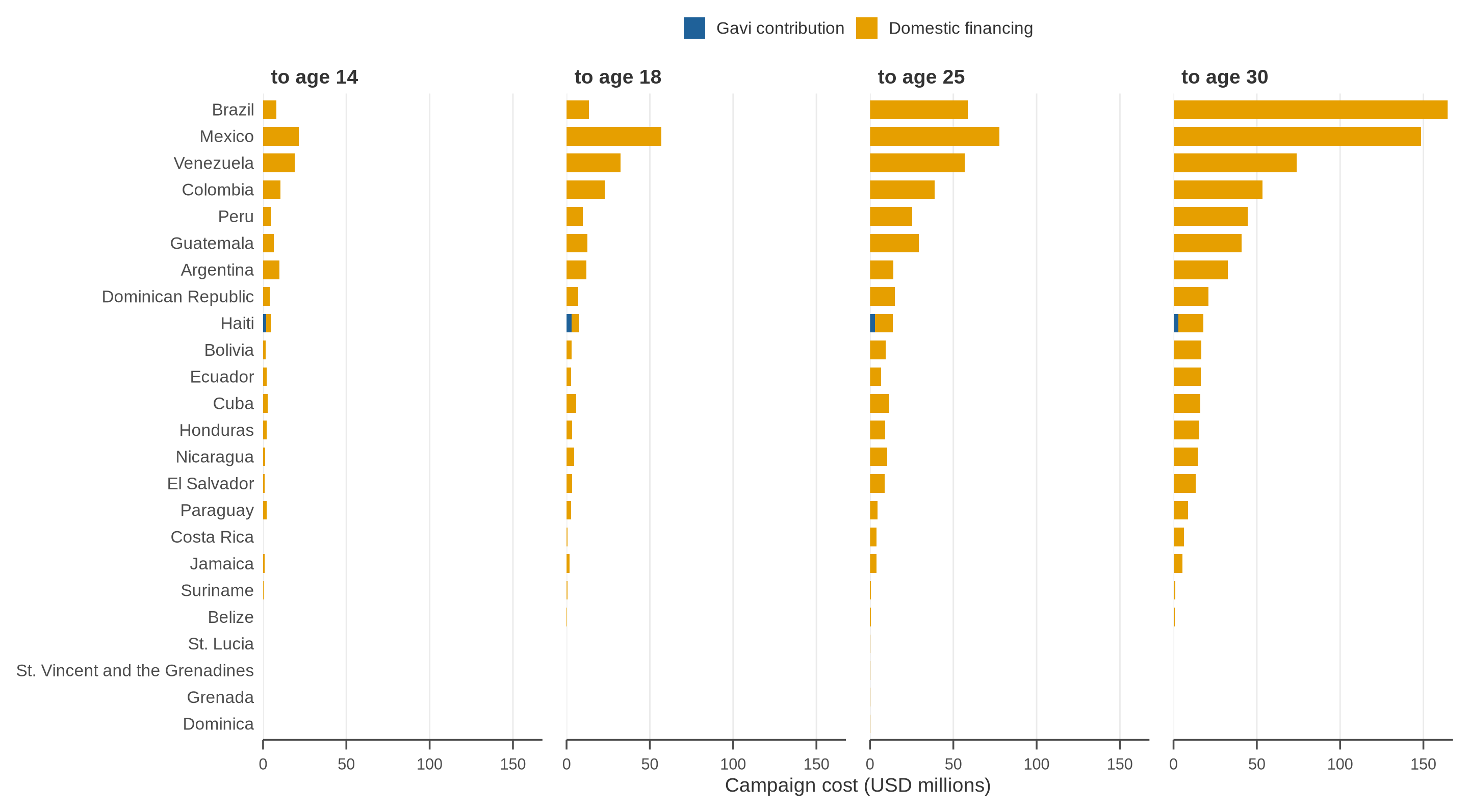

**Eastern Mediterranean Region**

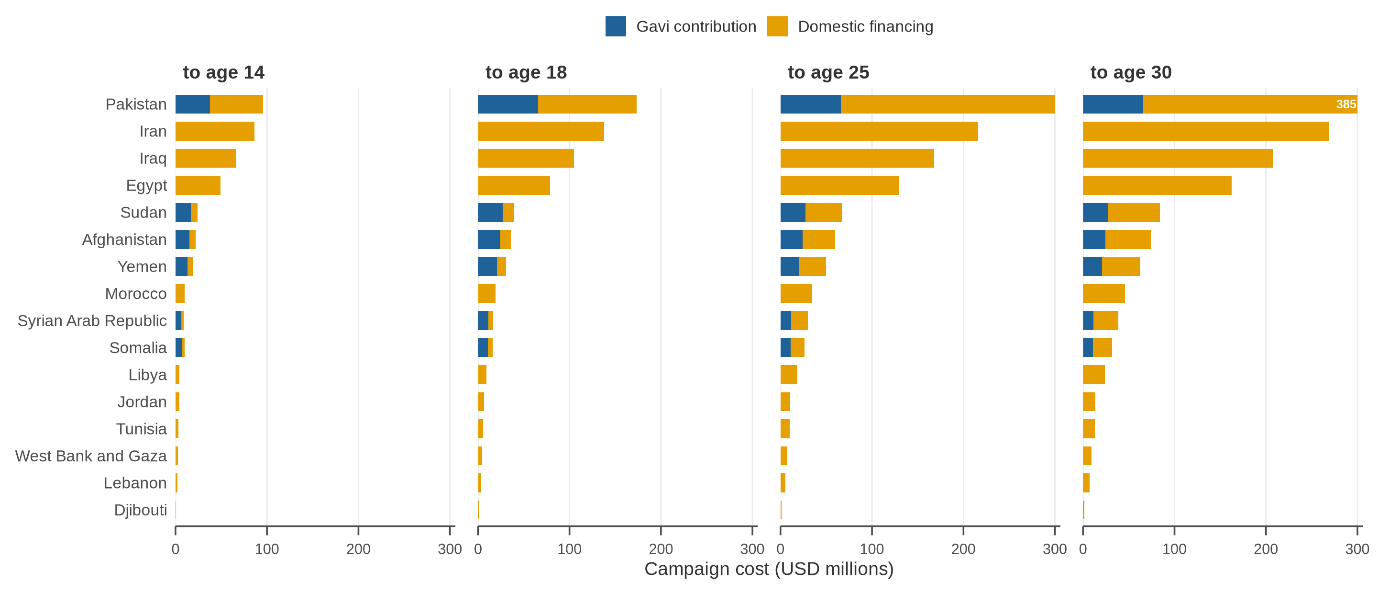

**European Region**

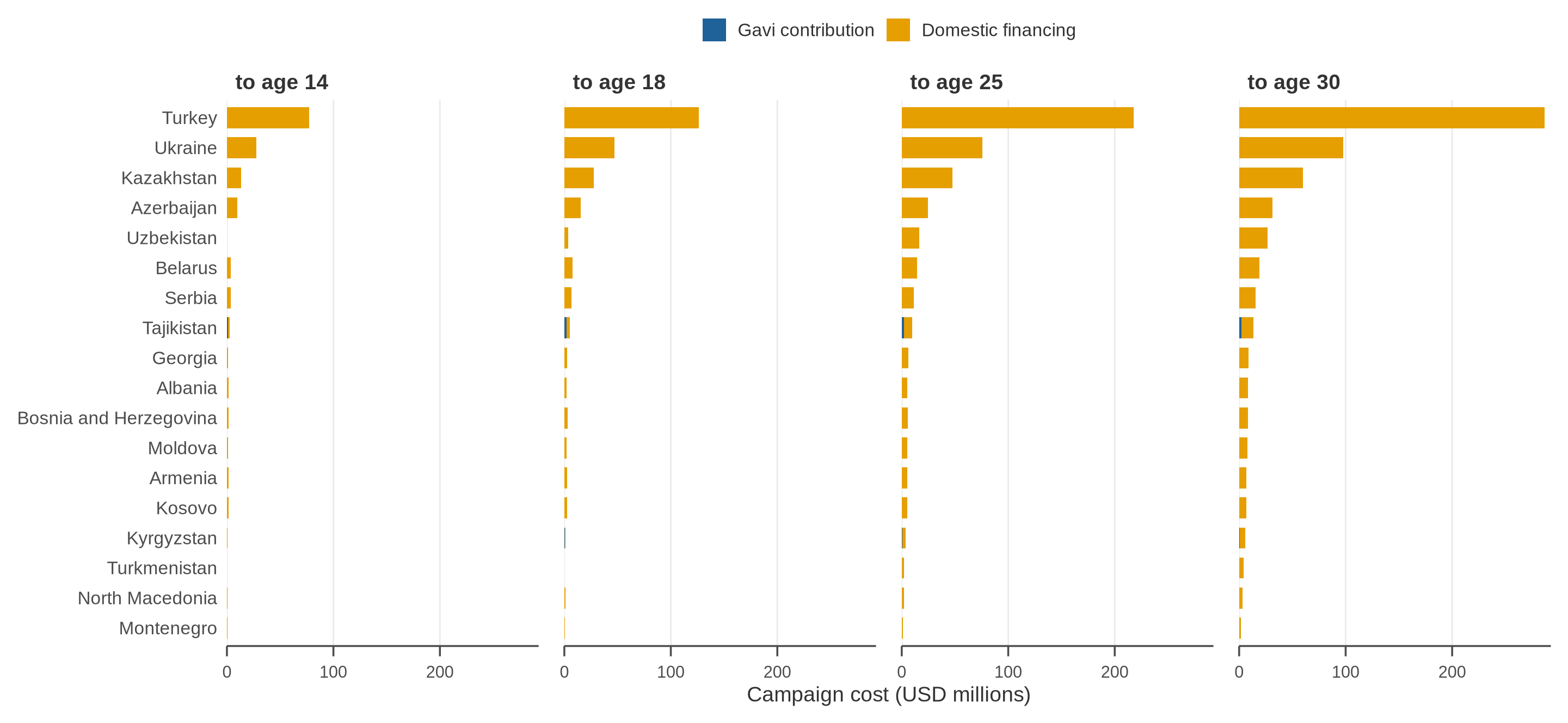

**Figure S4: Estimated cost of female catch-up HPV vaccination (assuming a vaccine targeting HPV 16/18 and 90% vaccine coverage) up to ages 14, 18, 25 and 30 years, by country, divided into the Gavi contribution and domestic financing, with one panel per WHO region.** Costs are in United States dollars (USD). Financial cost (and not economic cost) is considered. The Gavi contribution is the difference between the full campaign cost and what a Gavi-eligible country typically pays under the Gavi 6.0 co-financing guidelines: a share of the vaccine price (4% in the initial self-financing phase, 50% in the preparatory transition phase and 80% in the accelerated transition phase) together with a flat contribution to delivery cost, both applying only to doses given up to age 18. Countries in the Catalytic Phase, fully self-financing countries and countries outside the Gavi co-financing table receive the Gavi-negotiated vaccine price but no co-financing. Across all countries Gavi supports US$ 458 million, which is 10.6% of the cost of a campaign to age 18, for catch-up vaccination up to age 18 years, up to which age Gavi supports MAC campaign according to their latest vaccine funding guidelines (Gavi 6.0). For total vaccination cost up to age 30, the proportion decreased to 4.2%, as there is unlikely Gavi support for vaccinating ages 19-30. The share differs markedly by region, from 41% of the African Region's campaign to age 18 to 0.3% of the Western Pacific Region. Countries are ordered by the cost of the widest campaign, and the horizontal axis is identical on every regional sheet so that countries can be compared across regions; it is capped, and a country whose campaign costs more than the cap is drawn to the edge with its true cost printed on the bar.

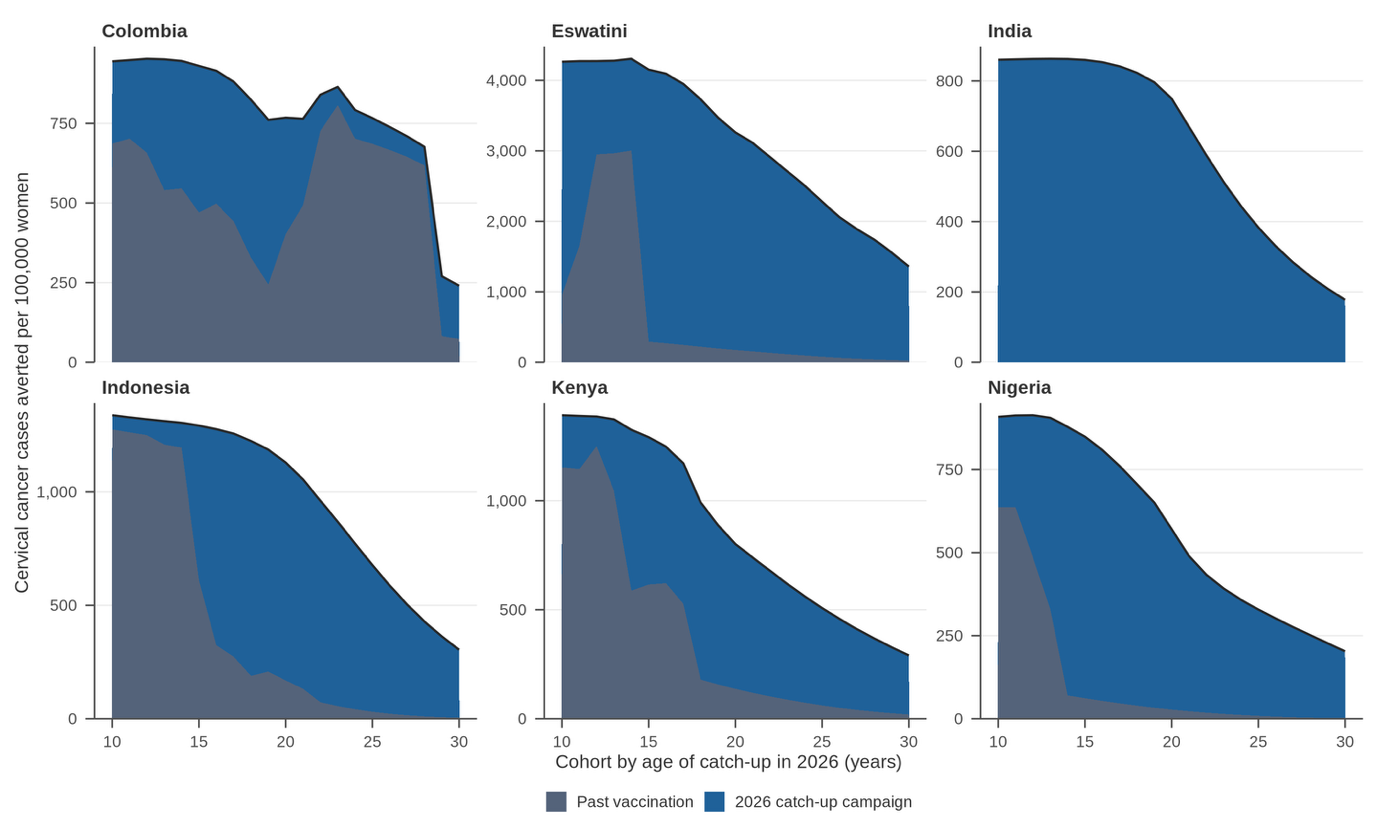

**Figure S5: Cervical cancer cases averted (per 100,000 women alive in 2026) by past vaccination (grey) and a one-time female catch-up HPV vaccination (blue, with a vaccine targeting HPV16/18 and 90% coverage) by birth cohort aged 9–30 years in 2026, in the six selected countries for cost-effectiveness analysis.** The vertical axis scale varies between panels, because the six countries' incidence differs by orders of magnitude and a shared axis would flatten five of them. The companion figure, Figure 3, shows the same quantities as a percentage of each cohort's own expected burden.

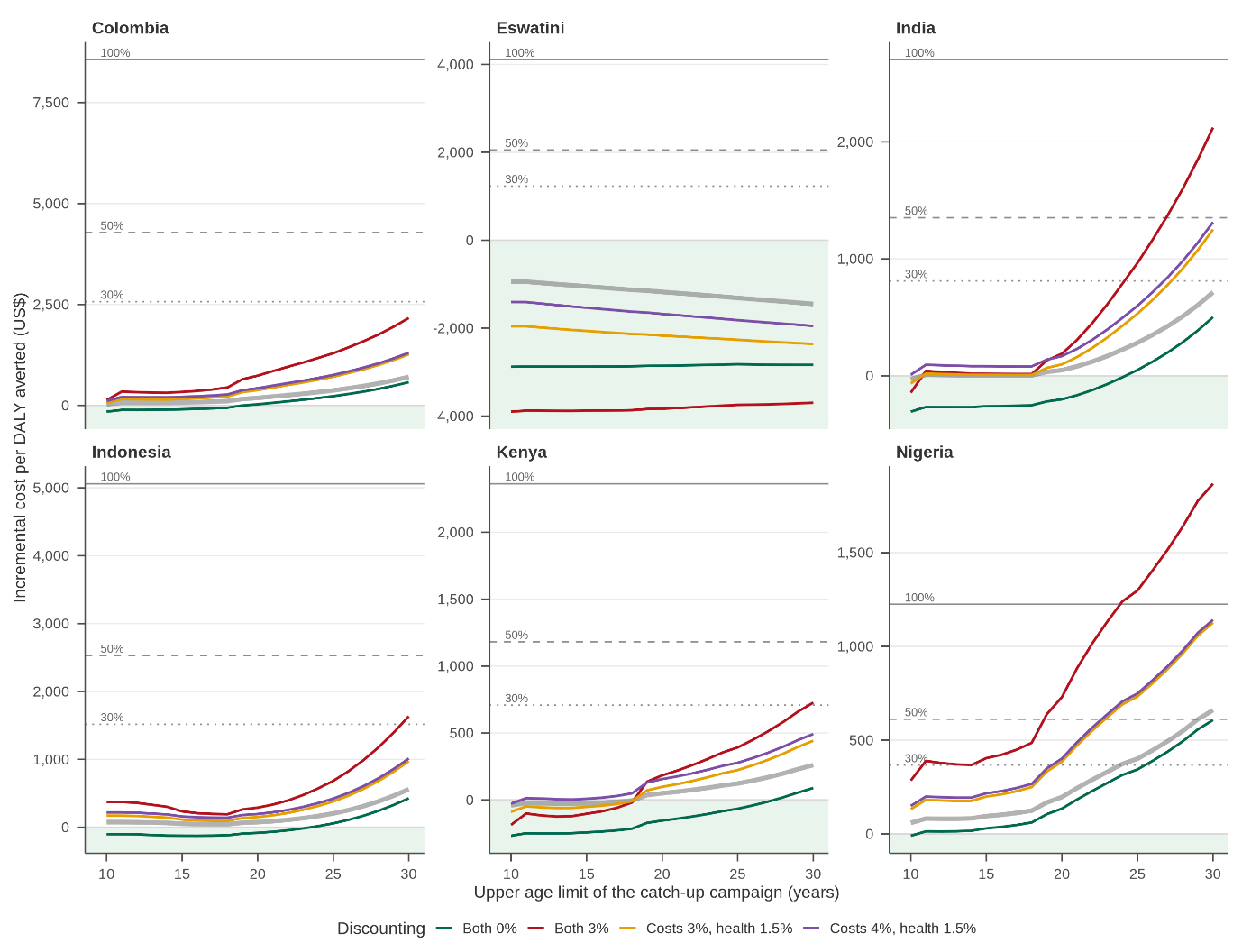

**Figure S6: Sensitivity analyses on discount rate. Incremental cost-effectiveness ratio (ICER) of female HPV catch-up vaccination assuming a vaccine targeting HPV16/18 and 90% coverage.** ICER estimates are reported for each incremental catch-up age. The thick grey curve corresponds to the base case.

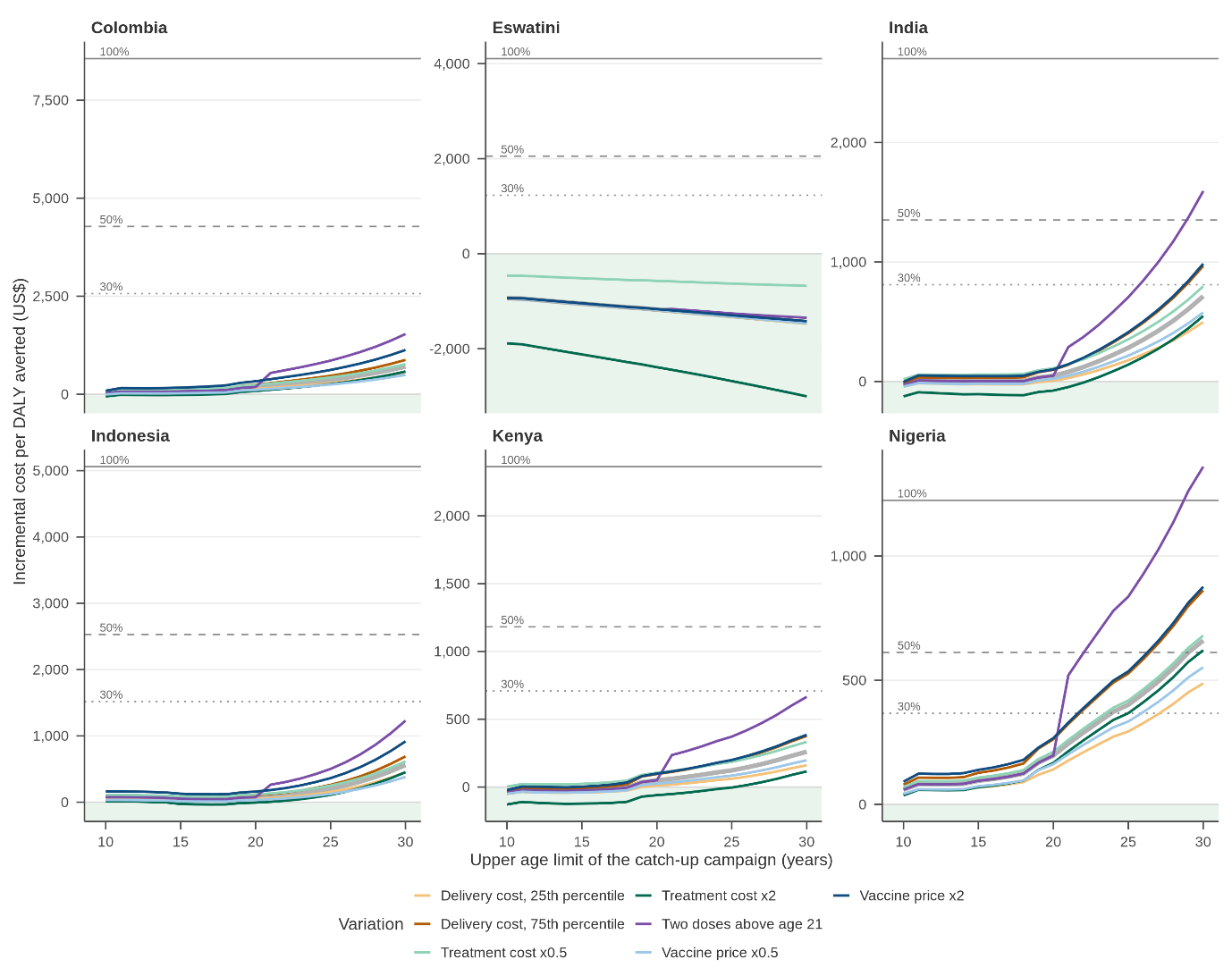

**Figure S7: Sensitivity analyses on cost variables and dose schedule above 21. Incremental cost-effectiveness ratio (ICER) of female HPV catch-up vaccination assuming a vaccine targeting HPV16/18 and 90% coverage.** ICER estimates are reported for each incremental catch-up age. DALYs are calculated using a 0% discount rate, while costs are discounted at 3%. Solid and hollow dots represent being on and not on the efficiency-frontier, respectively. The thick grey curve corresponds to the base case.

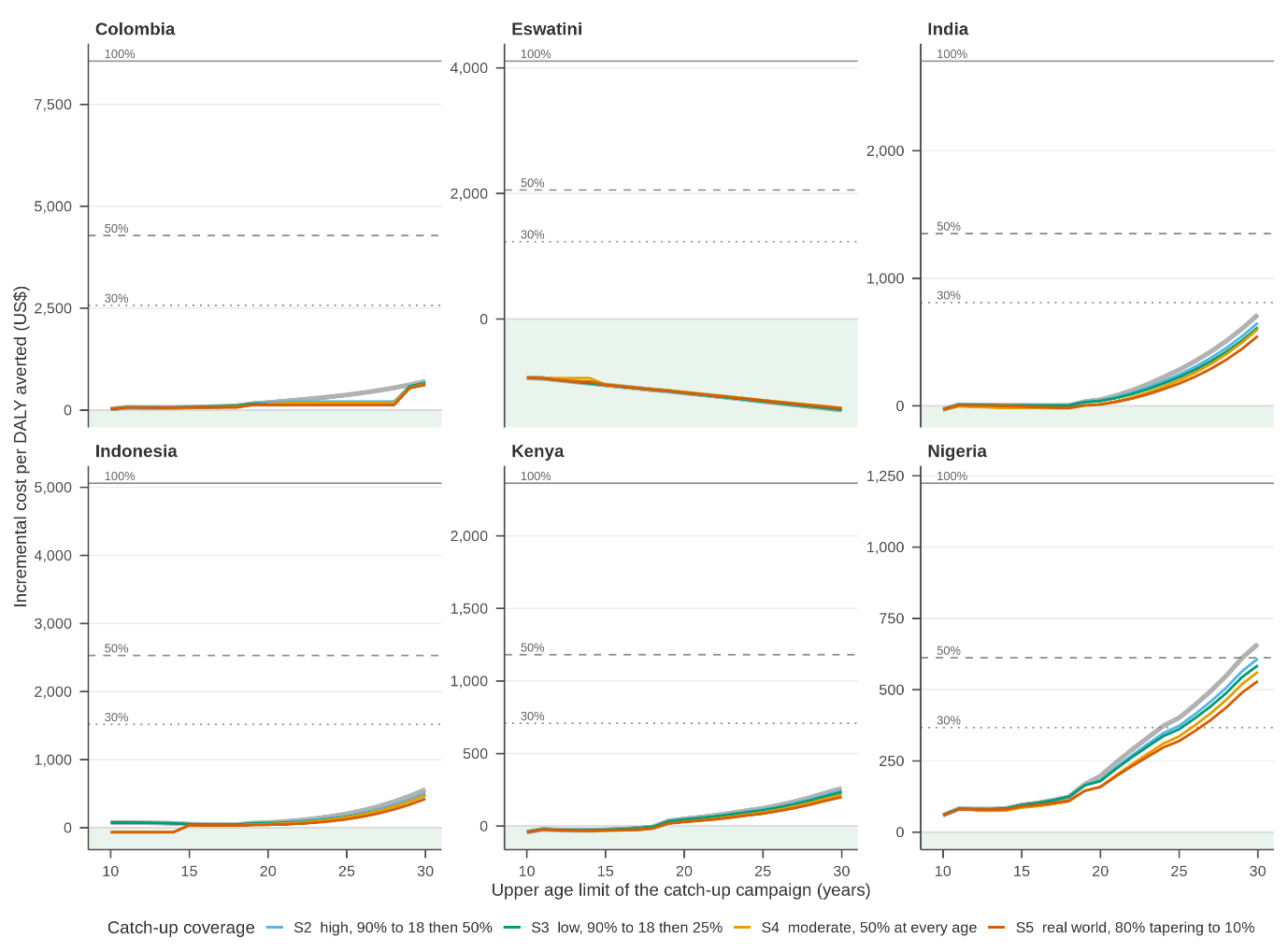

**Figure S8: Sensitivity analyses on catch-up coverage. Incremental cost-effectiveness ratio (ICER) of female HPV catch-up vaccination assuming a vaccine targeting HPV16/18 and 90% coverage.** ICER estimates are reported for each incremental catch-up age. DALYs are calculated using a 0% discount rate, while costs are discounted at 3%. The thick grey curve corresponds to the base case.

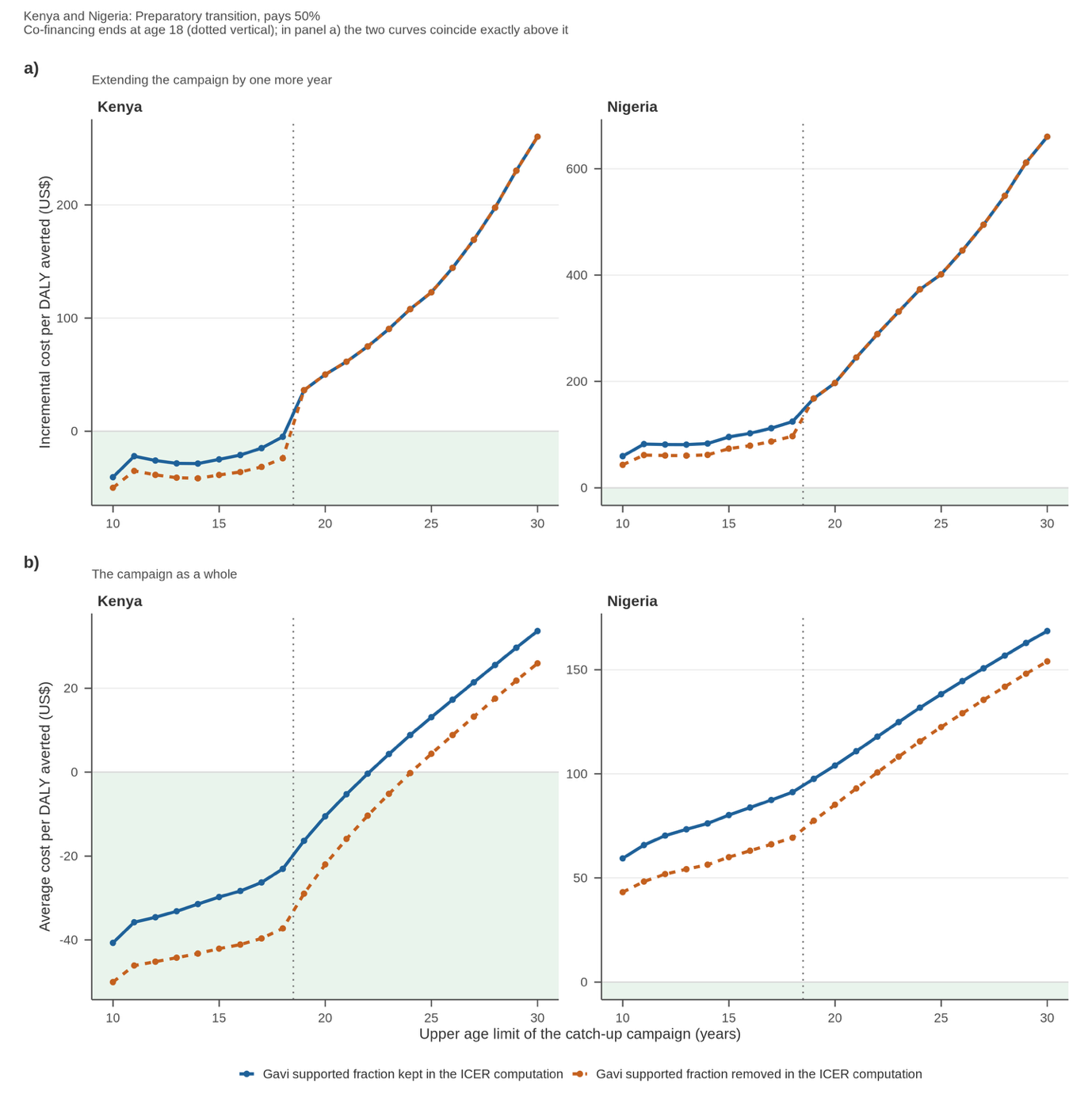

**Figure S9: Incremental cost-effectiveness ratio (ICER) of female HPV catch-up vaccination (assuming a vaccine targeting HPV16/18 and 90% coverage) of (a) extending the campaign by one year of age incrementally or (b) considering the campaign as a whole versus routine vaccination levels with and without the Gavi supported co-financing part included in the ICER computation**. Of the six representative countries for which cost-effectiveness was calculated, only the two countries that receive Gavi co-financing are shown; the other four are beyond the “active transition” phase and are unaffected by co-financing changes. Under the Gavi 6.0 guidelines a country in an active transition phase pays a share of the vaccine price rather than the full price (4% in initial self-financing, increasing fraction of 15% more each year in preparatory transition, 80% in accelerated transition), and only for doses delivered up to age 18. Kenya and Nigeria are both in the preparatory transition phase. As it is unclear to which fraction their country’s contribution would be increased at the time of catch-up, we assumed 50%. Values below zero mean the campaign saves more in averted cervical cancer treatment than it costs, so a curve moving down across zero becomes more cost-saving, not less.

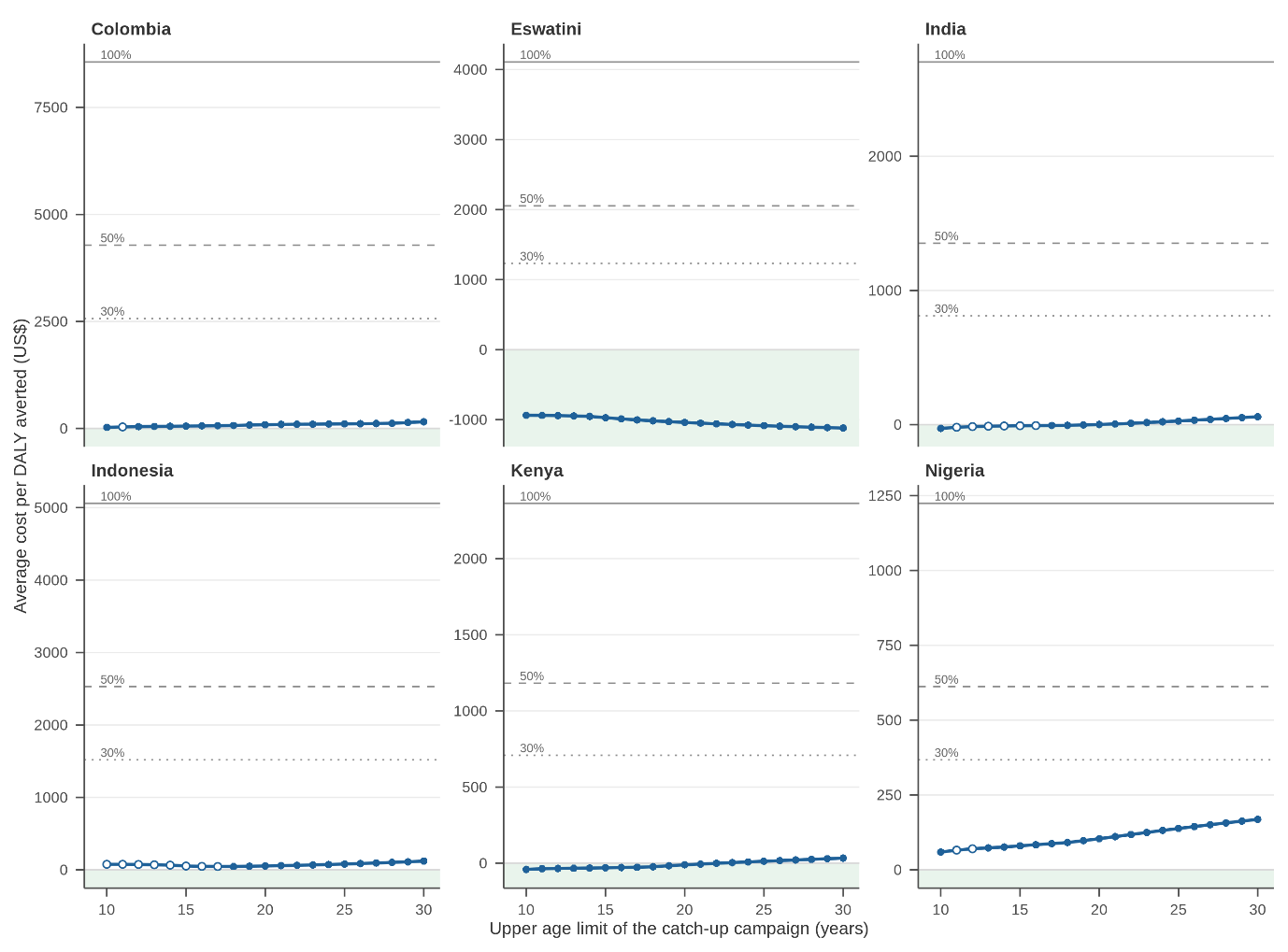
**Figure S10: Overall Incremental cost-effectiveness ratio (ICER) of female HPV catch-up vaccination assuming a vaccine targeting HPV16/18 and 90% coverage, and accounting for past vaccination.** Unlike Figure 4 of the main text, this figure displays the overall ICER estimates, meaning ICER estimates comparing the impact of the entire catch-up program to routine vaccination only (instead of by 1-year incremental age step). DALYs are calculated using a 0% discount rate, while costs are discounted at 3% (vaccine cost is undiscounted). The horizontal lines represent different cost-effectiveness thresholds at 30%, 50% and 100% of GDP per capita of each country, and cost-saving at 0%. Solid and hollow dots represent being on and not on the efficiency-frontier, respectively.

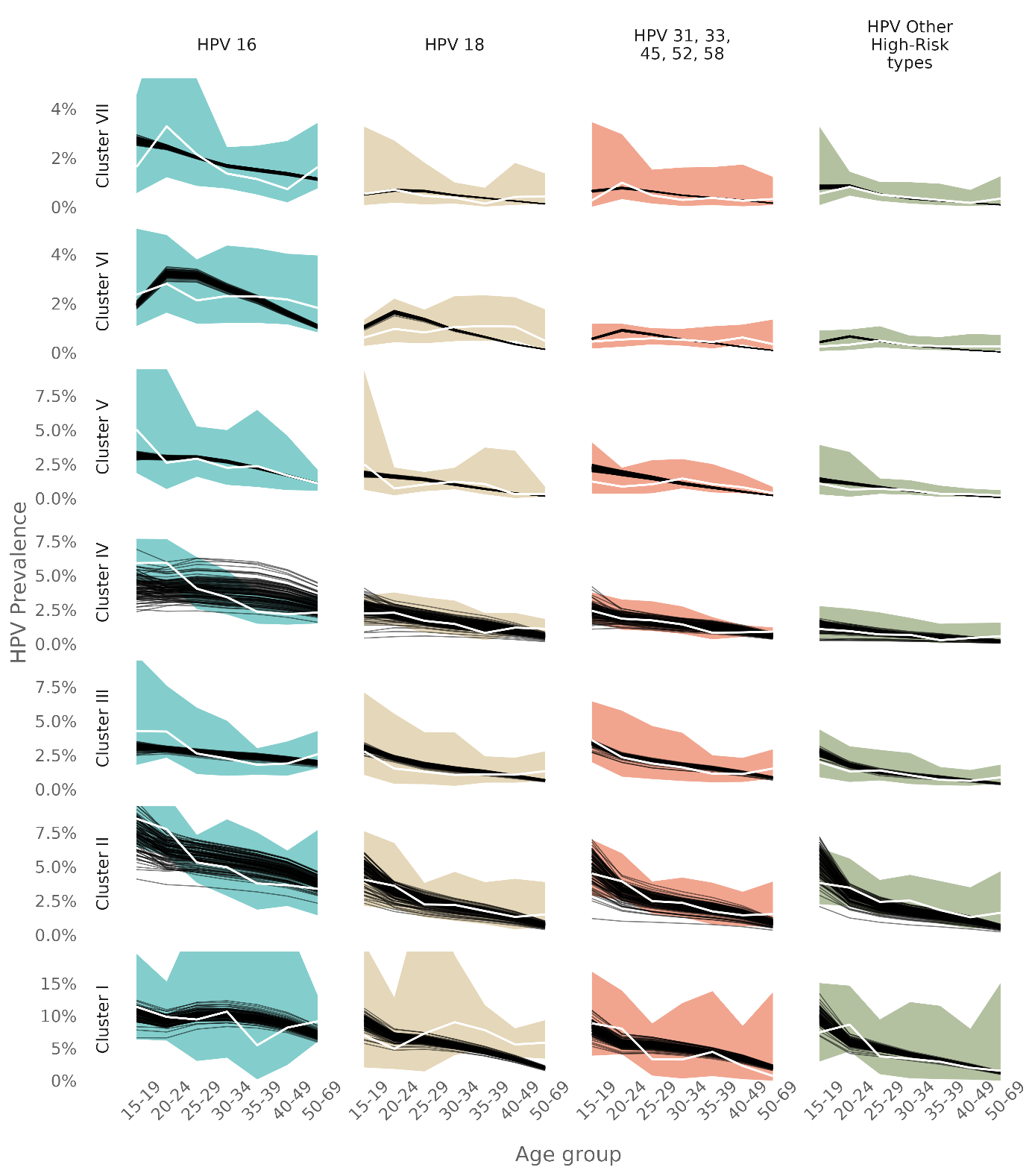

**Figure S11: Model fit to type-specific HPV prevalence by sexual behavior cluster and HPV group**. Model estimates from the 100 best-fitting parameter sets are shown as separate thin black lines. Observed HPV prevalence point estimates are shown by the white line, with colored shading indicating the 95% confidence intervals. Model calibration is described in details in Man et al.^1^

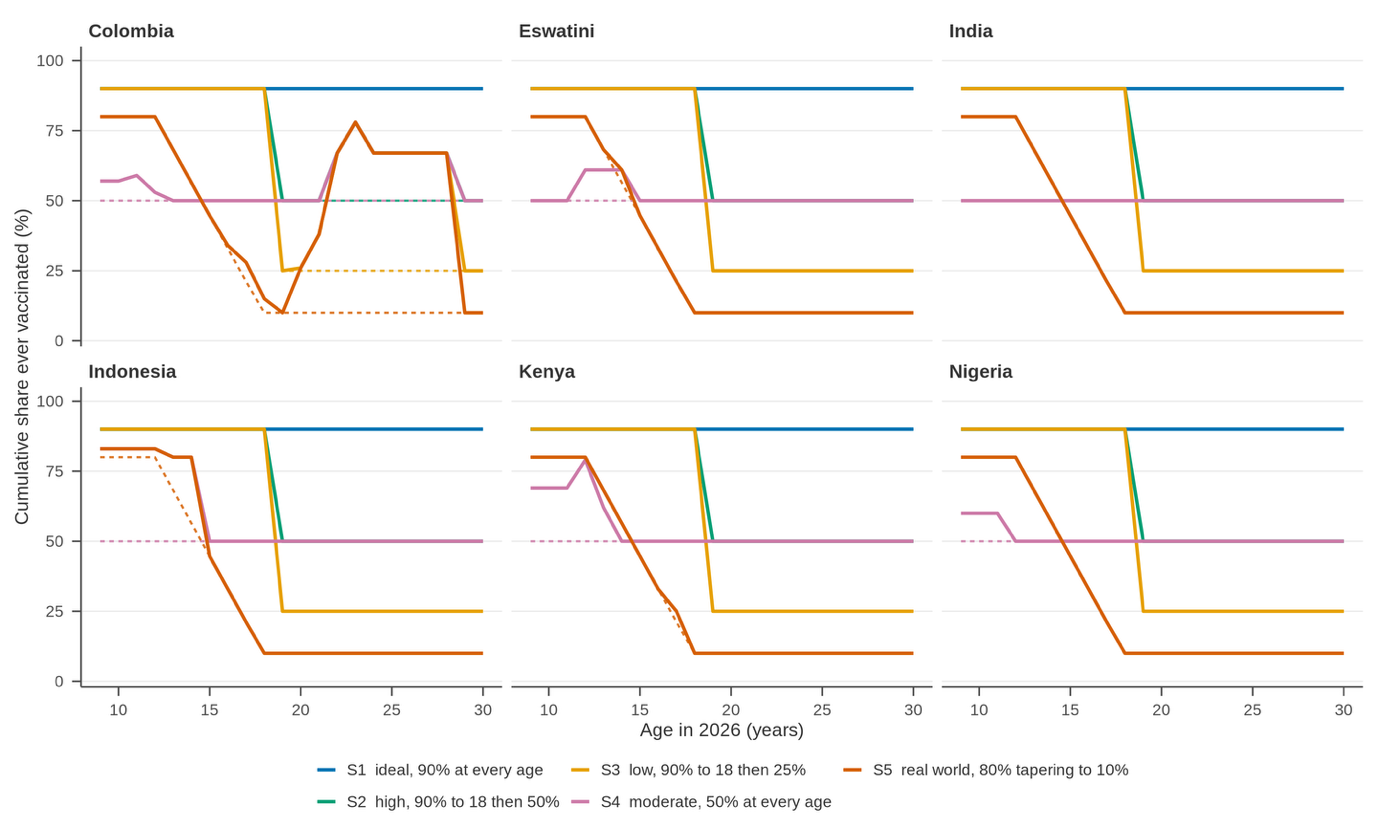

**Figure S12: HPV vaccination coverage assumed under each catch-up scenario, by birth cohort of different age in 2026, in the six countries selected for cost-effectiveness analysis.** Coverage levels for S1-S5 are as listed in the figure’s legend. S5 corresponds to real-world data of coverage observed in Rwanda.^2^ Cohorts already vaccinated by a past vaccination program according to WHO Dashboard^3^ start with coverage above zero and the campaign tops it up towards the scenario target. India has no recorded HPV vaccination before 2026, so its curves are the campaign alone. Solid lines show the coverage assumed for each analysis (which includes when real-world coverage levels exceed the level assumed in each S as indicated by the non-linear segments) and dotted lines show for illustrative purposes what the S coverage assumptions would have been without real-world vaccination history accounted for.

**Table S1: Cervical cancer cases averted (with 95%UI) by past vaccination and a one-time female catch-up HPV vaccination (with a vaccine targeting HPV16/18 and 90% coverage) up to age 14, 18, 25, or 30 in the lifetime of birth cohort aged 9–30 years in 2026.** The 2^nd^ to the 5^th^ columns from left to right correspond to the cumulative impact of adding each catch-up age band in the lifetime of the entire birth cohorts aged 9-30 years in 2026. The impact is the sum of direct and herd effect. The last column is the burden remaining after past vaccination and a catch-up up to age 30. Model outcomes were summarized across the 100 best fitting parameter sets obtained by model calibration, reporting medians and 95% uncertainty intervals (UI).

| **All LMICs / WHO Region / Country** | **Cervical cancer cases averted**  (in the lifetime of birth cohort aged 9–30 years in 2026, with 95%UI) | | | | | **Cervical cancer cases remaining** |
| --- | --- | --- | --- | --- | --- | --- |
|  | **Past vaccination** | **Catch-up  ages 9-14** | **Catch-up  ages 15-18** | **Catch-up  ages 19-25** | **Catch-up  ages 26-30** |  |
| All 132 LMICs | 2,701,294 (2,677,747–2,729,549) | 2,727,393 (2,723,245–2,730,208) | 4,449,272 (4,423,367–4,473,350) | 6,321,046 (6,250,856–6,398,029) | 6,931,875 (6,848,044–7,028,764) | 9,574,905 (9,472,018–9,667,374) |
| African | 1,491,604 (1,474,907–1,516,303) | 848,429 (845,986–850,462) | 1,369,214 (1,344,855–1,392,864) | 1,935,462 (1,871,166–2,006,522) | 2,139,170 (2,063,129–2,228,500) | 4,335,549 (4,234,324–4,422,009) |
| South-East Asia | 385,468 (384,620–386,370) | 839,661 (838,951–840,552) | 1,450,914 (1,449,847–1,452,103) | 2,156,776 (2,152,328–2,161,342) | 2,352,518 (2,346,532–2,358,453) | 2,193,757 (2,187,495–2,200,456) |
| Western Pacific | 106,775 (106,277–107,301) | 704,951 (704,279–705,761) | 1,085,407 (1,084,638–1,086,260) | 1,474,243 (1,471,823–1,476,822) | 1,583,968 (1,580,567–1,587,454) | 1,529,990 (1,526,458–1,533,659) |
| Americas | 635,149 (622,326–657,394) | 99,347 (96,492–99,804) | 173,927 (168,156–177,110) | 258,762 (240,890–270,491) | 326,209 (296,673–344,992) | 907,321 (869,822–937,882) |
| Eastern Mediterranean | 13,081 (13,026–13,140) | 183,097 (182,718–183,483) | 276,131 (275,015–277,404) | 357,537 (355,235–360,074) | 380,143 (377,558–383,000) | 402,918 (400,000–405,480) |
| European | 67,985 (67,927–68,077) | 52,248 (52,208–52,311) | 94,205 (93,998–94,354) | 139,101 (138,103–139,852) | 151,325 (150,351–152,182) | 206,788 (205,889–207,774) |
| Afghanistan | 0 (0–0) | 18,298 (18,279–18,321) | 27,264 (27,243–27,290) | 35,931 (35,885–35,988) | 37,963 (37,900–38,042) | 26,064 (25,984–26,126) |
| Albania | 551 (550–552) | 287 (286–287) | 573 (571–573) | 1,020 (1,012–1,026) | 1,177 (1,169–1,185) | 2,286 (2,277–2,294) |
| Algeria | 0 (0–0) | 17,872 (17,852–17,912) | 25,302 (25,174–25,520) | 30,701 (30,420–31,087) | 32,422 (32,100–32,842) | 39,960 (39,541–40,282) |
| Angola | 24,070 (23,309–24,912) | 45,702 (45,402–45,967) | 70,159 (68,055–72,183) | 89,539 (85,193–94,170) | 95,795 (90,883–101,133) | 143,581 (138,099–148,614) |
| Argentina | 62,280 (61,083–64,377) | 5,502 (4,755–5,761) | 6,583 (5,759–6,869) | 7,164 (6,366–7,465) | 10,489 (9,621–10,965) | 52,199 (50,754–53,307) |
| Armenia | 1,083 (1,081–1,086) | 493 (492–493) | 827 (826–828) | 1,324 (1,314–1,332) | 1,474 (1,465–1,483) | 2,429 (2,419–2,441) |
| Azerbaijan | 0 (0–0) | 3,353 (3,351–3,358) | 5,046 (5,038–5,052) | 6,443 (6,413–6,469) | 6,793 (6,762–6,822) | 6,103 (6,074–6,134) |
| Bangladesh | 68,444 (68,060–68,863) | 57,896 (57,661–58,128) | 104,171 (103,940–104,415) | 174,566 (174,452–174,666) | 193,087 (192,922–193,323) | 213,415 (212,765–213,915) |
| Belarus | 1,874 (1,871–1,878) | 1,251 (1,251–1,252) | 2,881 (2,874–2,887) | 4,214 (4,186–4,237) | 4,559 (4,531–4,584) | 6,334 (6,307–6,362) |
| Belize | 462 (451–479) | 107 (103–108) | 158 (153–161) | 317 (289–337) | 381 (344–408) | 919 (878–957) |
| Benin | 4,685 (4,538–4,836) | 7,987 (7,928–8,042) | 12,480 (12,087–12,857) | 16,197 (15,375–17,076) | 17,457 (16,520–18,474) | 27,411 (26,369–28,361) |
| Bhutan | 1,964 (1,963–1,966) | 0 (0–0) | 1 (1–1) | 2 (2–2) | 25 (25–25) | 679 (678–680) |
| Bolivia | 25,341 (24,846–26,184) | 2,395 (2,206–2,458) | 4,095 (3,869–4,200) | 8,013 (7,306–8,430) | 10,663 (9,587–11,366) | 36,168 (34,790–37,350) |
| Bosnia and Herzegovina | 32 (31–32) | 771 (770–772) | 1,300 (1,298–1,302) | 1,847 (1,836–1,856) | 2,011 (2,000–2,021) | 2,553 (2,542–2,564) |
| Botswana | 10,014 (9,929–10,196) | 1,515 (1,462–1,538) | 2,344 (2,276–2,379) | 3,037 (2,955–3,106) | 3,829 (3,728–3,944) | 11,798 (11,566–11,878) |
| Brazil | 200,345 (197,425–205,211) | 4,155 (3,672–4,401) | 6,669 (6,064–6,985) | 17,502 (15,663–18,593) | 34,626 (30,268–37,739) | 211,551 (205,377–216,774) |
| Burkina Faso | 20,158 (20,116–20,217) | 3,810 (3,791–3,843) | 13,955 (13,775–14,275) | 22,062 (21,669–22,632) | 24,459 (24,004–25,062) | 55,449 (54,868–55,905) |
| Burundi | 0 (0–0) | 26,891 (26,549–27,280) | 36,745 (35,837–37,618) | 44,067 (42,340–45,837) | 46,386 (44,438–48,486) | 54,735 (52,634–56,682) |
| Cabo Verde | 427 (426–428) | 0 (0–0) | 128 (125–132) | 299 (291–308) | 364 (355–376) | 1,168 (1,157–1,177) |
| Cambodia | 8,296 (8,276–8,319) | 3,674 (3,670–3,677) | 10,059 (10,019–10,096) | 17,305 (17,128–17,444) | 19,539 (19,314–19,707) | 24,574 (24,411–24,799) |
| Cameroon | 21,863 (21,333–22,820) | 27,488 (27,175–27,671) | 44,954 (43,648–45,988) | 59,533 (56,629–62,607) | 64,549 (61,103–68,064) | 107,167 (103,130–110,827) |
| Central African Republic | 0 (0–0) | 6,941 (6,847–7,042) | 9,550 (9,305–9,784) | 11,754 (11,262–12,254) | 12,517 (11,960–13,127) | 16,086 (15,476–16,643) |
| Chad | 0 (0–0) | 32,936 (32,898–33,010) | 47,042 (46,801–47,457) | 58,052 (57,494–58,800) | 61,181 (60,550–61,999) | 76,165 (75,347–76,796) |
| China | 0 (0–0) | 582,927 (582,287–583,687) | 888,356 (887,674–889,205) | 1,185,238 (1,183,714–1,187,187) | 1,266,331 (1,264,143–1,269,180) | 1,188,984 (1,186,134–1,191,172) |
| Colombia | 46,513 (45,234–48,977) | 7,816 (7,287–7,890) | 14,671 (13,832–15,060) | 20,095 (18,843–20,786) | 22,439 (20,941–23,219) | 53,292 (51,033–55,242) |
| Comoros | 248 (239–263) | 1,397 (1,384–1,407) | 2,012 (1,958–2,057) | 2,538 (2,425–2,658) | 2,734 (2,604–2,875) | 3,923 (3,776–4,056) |
| Congo | 0 (0–0) | 11,113 (10,969–11,274) | 15,214 (14,835–15,577) | 18,326 (17,599–19,069) | 19,355 (18,533–20,248) | 23,347 (22,454–24,169) |
| Congo Democratic Republic | 0 (0–0) | 223,364 (220,545–226,487) | 300,775 (293,467–307,857) | 362,838 (348,636–377,437) | 383,826 (367,707–401,483) | 465,631 (447,975–481,750) |
| Costa Rica | 2,139 (2,099–2,189) | 40 (39–40) | 284 (258–303) | 1,057 (916–1,165) | 1,383 (1,195–1,525) | 4,161 (3,975–4,338) |
| Cote d'Ivoire | 48,080 (47,373–48,771) | 6,850 (6,730–6,959) | 21,526 (20,182–22,802) | 37,480 (34,385–40,923) | 43,095 (39,573–47,087) | 115,554 (111,367–119,270) |
| Cuba | 1,796 (1,706–1,896) | 2,338 (2,310–2,369) | 4,349 (4,175–4,516) | 6,544 (6,068–6,963) | 7,466 (6,875–7,980) | 11,766 (11,193–12,345) |
| Djibouti | 35 (35–36) | 698 (697–699) | 1,057 (1,050–1,067) | 1,392 (1,376–1,412) | 1,499 (1,480–1,522) | 2,221 (2,198–2,239) |
| Dominica | 43 (42–45) | 4 (4–4) | 9 (9–9) | 28 (25–31) | 37 (32–40) | 104 (99–108) |
| Dominican Republic | 7,559 (7,341–7,933) | 4,142 (4,071–4,159) | 6,686 (6,522–6,785) | 10,344 (9,638–10,860) | 11,980 (11,039–12,683) | 22,308 (21,290–23,255) |
| Ecuador | 30,665 (30,180–31,495) | 1,825 (1,613–1,906) | 2,185 (1,948–2,284) | 3,461 (3,186–3,600) | 5,743 (5,176–6,095) | 30,031 (29,202–30,739) |
| Egypt | 0 (0–0) | 14,882 (14,865–14,915) | 21,260 (21,151–21,448) | 26,446 (26,186–26,787) | 28,078 (27,775–28,456) | 36,517 (36,139–36,820) |
| El Salvador | 4,292 (4,172–4,429) | 945 (937–951) | 3,164 (2,974–3,318) | 5,874 (5,302–6,342) | 7,139 (6,406–7,738) | 15,083 (14,370–15,790) |
| Equatorial Guinea | 0 (0–0) | 1,413 (1,395–1,433) | 1,916 (1,870–1,962) | 2,299 (2,210–2,391) | 2,415 (2,314–2,523) | 2,815 (2,707–2,916) |
| Eritrea | 3,619 (3,610–3,627) | 982 (979–987) | 1,366 (1,359–1,384) | 2,693 (2,647–2,752) | 3,120 (3,062–3,192) | 8,863 (8,789–8,928) |
| Eswatini | 1,973 (1,951–2,016) | 2,198 (2,180–2,214) | 4,198 (4,175–4,262) | 6,371 (6,270–6,600) | 7,185 (7,056–7,484) | 11,293 (10,957–11,411) |
| Ethiopia | 116,912 (116,529–117,222) | 48,148 (48,051–48,316) | 91,996 (91,187–93,470) | 148,766 (146,395–151,985) | 166,658 (163,834–170,327) | 381,636 (378,023–384,668) |
| Fiji | 3,130 (3,124–3,136) | 37 (37–38) | 86 (85–87) | 590 (579–599) | 886 (868–900) | 2,888 (2,872–2,909) |
| Gabon | 0 (0–0) | 2,093 (2,066–2,123) | 2,834 (2,765–2,901) | 3,426 (3,291–3,565) | 3,630 (3,476–3,799) | 4,450 (4,281–4,603) |
| Gambia | 3,789 (3,773–3,802) | 4,497 (4,490–4,510) | 6,424 (6,392–6,484) | 8,872 (8,772–9,009) | 9,633 (9,513–9,786) | 16,922 (16,767–17,052) |
| Georgia | 2,113 (2,110–2,117) | 953 (952–954) | 1,952 (1,947–1,955) | 2,951 (2,929–2,968) | 3,286 (3,265–3,306) | 5,255 (5,233–5,277) |
| Ghana | 23,176 (22,471–23,796) | 31,228 (30,888–31,541) | 51,294 (49,481–53,036) | 68,595 (64,790–72,700) | 74,601 (70,263–79,339) | 125,686 (120,927–130,019) |
| Grenada | 20 (20–21) | 53 (52–54) | 72 (70–73) | 105 (98–110) | 119 (111–126) | 177 (168–185) |
| Guatemala | 19,164 (18,642–19,973) | 9,015 (8,889–9,053) | 15,914 (15,384–16,257) | 27,031 (24,879–28,661) | 31,237 (28,503–33,334) | 58,565 (55,799–61,246) |
| Guinea | 0 (0–0) | 20,576 (20,552–20,623) | 29,810 (29,652–30,081) | 37,408 (37,030–37,903) | 39,875 (39,431–40,427) | 53,298 (52,747–53,742) |
| Guinea-Bissau | 0 (0–0) | 4,160 (4,155–4,170) | 6,064 (6,032–6,120) | 7,658 (7,579–7,761) | 8,163 (8,071–8,278) | 10,998 (10,884–11,091) |
| Haiti | 0 (0–0) | 10,621 (10,384–10,904) | 15,681 (15,230–16,155) | 20,889 (19,737–21,993) | 22,827 (21,454–24,197) | 27,537 (26,166–28,909) |
| Honduras | 14,310 (14,046–14,752) | 1,088 (1,001–1,120) | 1,802 (1,699–1,852) | 3,900 (3,524–4,133) | 5,334 (4,756–5,733) | 19,737 (18,989–20,372) |
| India | 0 (0–0) | 738,432 (737,556–739,480) | 1,159,605 (1,158,644–1,160,826) | 1,608,283 (1,606,017–1,611,246) | 1,726,392 (1,723,139–1,730,700) | 1,328,444 (1,324,135–1,331,696) |
| Indonesia | 224,181 (223,694–224,758) | 13,747 (13,699–13,807) | 117,973 (117,448–118,555) | 249,043 (246,051–251,495) | 288,693 (284,809–291,673) | 444,202 (441,251–448,200) |
| Iran | 0 (0–0) | 9,185 (9,177–9,197) | 13,814 (13,793–13,832) | 17,551 (17,469–17,621) | 18,448 (18,365–18,526) | 16,332 (16,254–16,416) |
| Iraq | 0 (0–0) | 8,747 (8,739–8,758) | 13,091 (13,071–13,108) | 16,763 (16,684–16,832) | 17,618 (17,536–17,694) | 15,740 (15,664–15,822) |
| Jamaica | 960 (928–1,018) | 1,421 (1,400–1,444) | 2,374 (2,307–2,432) | 3,510 (3,291–3,710) | 4,061 (3,766–4,324) | 6,642 (6,329–6,945) |
| Jordan | 0 (0–0) | 3,059 (3,057–3,063) | 4,608 (4,601–4,614) | 5,835 (5,808–5,858) | 6,136 (6,109–6,162) | 5,423 (5,397–5,451) |
| Kazakhstan | 4,331 (4,323–4,340) | 5,146 (5,143–5,149) | 10,078 (10,054–10,093) | 13,801 (13,717–13,867) | 14,632 (14,548–14,704) | 15,933 (15,855–16,017) |
| Kenya | 61,386 (60,317–62,789) | 16,278 (15,924–16,478) | 35,967 (34,444–37,350) | 58,598 (54,577–62,888) | 66,293 (61,644–71,374) | 161,305 (155,500–166,425) |
| Kiribati | 106 (106–106) | 15 (15–15) | 40 (40–40) | 92 (91–93) | 107 (105–108) | 154 (153–155) |
| Korea Dem. People's Rep. | 0 (0–0) | 8,972 (8,962–8,984) | 13,862 (13,851–13,875) | 19,161 (19,135–19,194) | 20,794 (20,756–20,844) | 21,737 (21,687–21,775) |
| Kosovo | 0 (0–0) | 402 (402–403) | 670 (669–671) | 927 (922–931) | 1,005 (1,000–1,010) | 1,229 (1,224–1,234) |
| Kyrgyzstan | 7,471 (7,465–7,479) | 171 (171–171) | 902 (898–905) | 3,110 (3,069–3,143) | 3,678 (3,636–3,714) | 9,777 (9,736–9,820) |
| Laos | 5,854 (5,842–5,867) | 332 (330–333) | 877 (874–880) | 3,593 (3,541–3,637) | 4,446 (4,375–4,501) | 9,104 (9,046–9,178) |
| Lebanon | 0 (0–0) | 1,321 (1,320–1,322) | 1,933 (1,930–1,935) | 2,487 (2,476–2,498) | 2,633 (2,621–2,645) | 2,492 (2,480–2,504) |
| Lesotho | 4,845 (4,801–4,942) | 1,689 (1,656–1,699) | 2,365 (2,346–2,379) | 4,524 (4,431–4,684) | 5,378 (5,260–5,615) | 11,755 (11,438–11,861) |
| Liberia | 5,697 (5,565–5,930) | 4,253 (4,148–4,301) | 6,962 (6,771–7,112) | 9,726 (9,209–10,225) | 10,645 (10,052–11,234) | 19,967 (19,231–20,623) |
| Libya | 1,820 (1,815–1,825) | 1,080 (1,078–1,083) | 2,259 (2,239–2,295) | 3,365 (3,315–3,435) | 3,681 (3,623–3,756) | 7,295 (7,218–7,357) |
| Madagascar | 23,414 (22,702–24,055) | 32,072 (31,742–32,378) | 52,862 (51,002–54,647) | 70,545 (66,647–74,746) | 76,952 (72,486–81,829) | 130,316 (125,406–134,784) |
| Malawi | 150,737 (147,776–154,883) | 16,310 (15,575–16,720) | 19,483 (18,941–19,955) | 48,022 (44,469–51,801) | 60,226 (55,379–65,226) | 254,521 (245,901–261,966) |
| Malaysia | 40,143 (40,025–40,237) | 381 (370–393) | 1,838 (1,799–1,879) | 2,276 (2,232–2,320) | 2,690 (2,650–2,726) | 20,213 (20,150–20,300) |
| Maldives | 311 (310–311) | 11 (11–12) | 22 (22–22) | 81 (80–82) | 113 (111–114) | 270 (268–273) |
| Mali | 20,261 (20,183–20,350) | 25,015 (24,973–25,103) | 44,637 (44,276–45,254) | 59,985 (59,220–61,082) | 64,285 (63,411–65,432) | 105,383 (104,249–106,258) |
| Marshall Islands | 72 (72–72) | 5 (4–5) | 13 (12–13) | 26 (26–27) | 35 (34–35) | 79 (78–79) |
| Mauritania | 4,093 (4,076–4,106) | 3,789 (3,784–3,802) | 6,259 (6,213–6,340) | 8,624 (8,514–8,778) | 9,336 (9,208–9,502) | 16,649 (16,485–16,784) |
| Mauritius | 611 (602–621) | 40 (38–42) | 49 (47–51) | 185 (167–204) | 254 (229–281) | 1,172 (1,141–1,199) |
| Mexico | 144,112 (141,005–149,503) | 8,739 (7,996–8,957) | 27,694 (25,887–28,708) | 34,118 (31,703–35,351) | 46,656 (42,777–48,759) | 162,424 (156,706–166,957) |
| Micronesia Fed. Sts. | 143 (142–143) | 16 (16–17) | 22 (22–23) | 48 (48–49) | 66 (66–67) | 164 (163–165) |
| Moldova | 2,015 (2,011–2,020) | 835 (834–836) | 1,283 (1,281–1,284) | 2,175 (2,158–2,188) | 2,482 (2,465–2,497) | 4,946 (4,928–4,963) |
| Mongolia | 1,093 (1,091–1,095) | 1,157 (1,157–1,158) | 2,265 (2,260–2,269) | 3,117 (3,099–3,132) | 3,341 (3,323–3,357) | 4,765 (4,747–4,783) |
| Montenegro | 61 (61–61) | 212 (212–212) | 373 (372–373) | 525 (522–527) | 568 (565–571) | 714 (711–717) |
| Morocco | 515 (512–517) | 15,796 (15,777–15,834) | 23,815 (23,678–24,049) | 30,643 (30,310–31,078) | 33,027 (32,627–33,517) | 47,807 (47,318–48,209) |
| Mozambique | 76,767 (75,637–78,094) | 11,763 (11,453–11,946) | 42,862 (40,540–44,985) | 67,949 (62,861–73,524) | 77,060 (71,255–83,473) | 189,381 (182,456–195,600) |
| Myanmar | 30,139 (30,071–30,219) | 7,404 (7,398–7,412) | 20,878 (20,788–20,971) | 43,056 (42,543–43,462) | 49,836 (49,179–50,335) | 72,865 (72,379–73,528) |
| Namibia | 992 (974–1,022) | 4,283 (4,264–4,311) | 6,465 (6,432–6,569) | 8,607 (8,499–8,869) | 9,415 (9,278–9,748) | 11,842 (11,490–11,963) |
| Nepal | 9,453 (9,438–9,470) | 6,632 (6,629–6,635) | 15,500 (15,494–15,507) | 25,364 (25,330–25,409) | 28,222 (28,167–28,295) | 31,544 (31,452–31,615) |
| Nicaragua | 4,508 (4,376–4,675) | 1,448 (1,424–1,456) | 4,235 (4,018–4,399) | 7,105 (6,485–7,596) | 8,269 (7,497–8,879) | 15,497 (14,747–16,243) |
| Niger | 0 (0–0) | 27,250 (27,219–27,311) | 38,881 (38,682–39,223) | 48,042 (47,579–48,659) | 50,679 (50,154–51,358) | 63,524 (62,846–64,049) |
| Nigeria | 94,450 (94,114–94,825) | 89,588 (89,463–89,918) | 172,861 (171,373–175,411) | 240,167 (236,876–244,784) | 259,821 (256,060–264,755) | 458,769 (453,917–462,611) |
| North Macedonia | 1,076 (1,073–1,078) | 154 (153–154) | 298 (297–299) | 436 (433–438) | 494 (491–496) | 1,196 (1,193–1,200) |
| Pakistan | 10,645 (10,593–10,701) | 38,696 (38,688–38,703) | 64,601 (64,584–64,616) | 88,771 (88,692–88,879) | 94,713 (94,586–94,877) | 74,328 (74,103–74,503) |
| Papua New Guinea | 0 (0–0) | 16,386 (16,356–16,419) | 24,815 (24,756–24,860) | 33,854 (33,627–34,016) | 36,145 (35,871–36,339) | 25,291 (25,097–25,565) |
| Paraguay | 16,667 (16,283–17,378) | 3,221 (2,932–3,304) | 3,664 (3,338–3,750) | 4,731 (4,431–4,850) | 6,313 (5,845–6,554) | 20,453 (19,832–20,971) |
| Peru | 52,892 (51,784–54,794) | 5,442 (5,149–5,554) | 10,682 (10,176–10,986) | 19,308 (17,595–20,326) | 25,504 (22,976–27,176) | 80,344 (77,175–83,087) |
| Philippines | 46,103 (45,738–46,467) | 49,661 (49,518–49,861) | 80,061 (79,882–80,262) | 121,232 (120,541–122,028) | 134,653 (133,611–135,597) | 153,206 (152,204–154,504) |
| Rwanda | 56,876 (56,229–57,903) | 305 (271–330) | 823 (768–870) | 1,581 (1,460–1,701) | 3,322 (2,985–3,610) | 43,944 (42,839–44,630) |
| Samoa | 120 (120–120) | 3 (3–3) | 14 (14–14) | 64 (63–65) | 79 (78–80) | 149 (148–150) |
| Sao Tome and Principe | 0 (0–0) | 288 (284–292) | 396 (386–406) | 493 (472–514) | 524 (501–550) | 682 (655–705) |
| Senegal | 36,421 (36,323–36,511) | 9,485 (9,466–9,527) | 19,109 (18,917–19,452) | 32,717 (32,152–33,482) | 36,885 (36,216–37,760) | 93,106 (92,255–93,827) |
| Serbia | 71 (71–72) | 2,532 (2,529–2,535) | 4,057 (4,051–4,063) | 5,510 (5,481–5,535) | 5,913 (5,883–5,941) | 6,746 (6,717–6,775) |
| Sierra Leone | 9,367 (9,200–9,548) | 2,581 (2,553–2,602) | 7,084 (6,729–7,410) | 10,974 (10,191–11,829) | 12,317 (11,429–13,292) | 27,910 (26,872–28,846) |
| Solomon Islands | 1,445 (1,439–1,451) | 319 (318–321) | 737 (734–739) | 1,272 (1,262–1,283) | 1,462 (1,447–1,476) | 2,009 (1,995–2,027) |
| Somalia | 0 (0–0) | 24,476 (24,155–24,822) | 33,286 (32,456–34,089) | 40,693 (39,034–42,383) | 43,061 (41,195–45,105) | 53,299 (51,255–55,165) |
| South Africa | 96,164 (95,449–97,591) | 10,380 (10,104–10,493) | 18,763 (18,375–19,019) | 31,947 (31,086–32,959) | 41,720 (40,546–43,587) | 135,048 (132,135–136,001) |
| South Sudan | 0 (0–0) | 13,564 (13,548–13,597) | 19,950 (19,840–20,137) | 24,974 (24,720–25,311) | 26,447 (26,159–26,819) | 34,431 (34,059–34,719) |
| Sri Lanka | 20,231 (20,186–20,274) | 1,056 (1,047–1,065) | 1,781 (1,769–1,793) | 6,655 (6,557–6,739) | 8,762 (8,617–8,878) | 18,630 (18,496–18,797) |
| St. Lucia | 118 (115–121) | 4 (4–4) | 22 (20–23) | 74 (64–81) | 99 (86–110) | 289 (276–302) |
| St. Vincent and the Grenadines | 14 (14–15) | 48 (47–49) | 76 (74–78) | 108 (101–114) | 123 (114–130) | 181 (172–190) |
| Sudan | 0 (0–0) | 31,968 (31,930–32,042) | 46,477 (46,230–46,901) | 58,614 (58,014–59,395) | 62,375 (61,674–63,244) | 83,315 (82,446–84,016) |
| Suriname | 750 (728–800) | 421 (403–423) | 525 (504–530) | 628 (603–640) | 728 (692–744) | 1,313 (1,265–1,355) |
| Syrian Arab Republic | 0 (0–0) | 4,212 (4,207–4,219) | 7,250 (7,237–7,260) | 9,693 (9,641–9,738) | 10,277 (10,223–10,327) | 10,158 (10,108–10,212) |
| Tajikistan | 3,187 (3,182–3,194) | 2,812 (2,810–2,814) | 5,776 (5,761–5,787) | 8,141 (8,087–8,184) | 8,748 (8,693–8,795) | 10,661 (10,611–10,714) |
| Tanzania | 146,296 (144,946–148,086) | 1,014 (989–1,041) | 3,328 (3,142–3,507) | 31,418 (27,838–35,486) | 41,024 (36,377–46,187) | 206,381 (200,057–211,730) |
| Thailand | 29,424 (29,299–29,549) | 5,235 (5,205–5,268) | 16,575 (16,509–16,651) | 29,360 (29,098–29,633) | 35,123 (34,716–35,467) | 59,620 (59,247–60,095) |
| Timor-Leste | 1,312 (1,308–1,317) | 283 (280–285) | 592 (590–594) | 1,349 (1,336–1,362) | 1,572 (1,553–1,587) | 2,487 (2,471–2,509) |
| Togo | 11,000 (10,813–11,249) | 2,934 (2,889–2,969) | 5,434 (5,221–5,635) | 8,893 (8,307–9,537) | 10,075 (9,376–10,845) | 25,437 (24,531–26,236) |
| Tonga | 25 (25–25) | 36 (36–36) | 70 (69–70) | 111 (110–112) | 123 (122–124) | 117 (116–118) |
| Tunisia | 67 (66–67) | 2,034 (2,031–2,038) | 2,957 (2,940–2,984) | 3,695 (3,658–3,745) | 3,965 (3,920–4,020) | 5,475 (5,419–5,520) |
| Turkey | 0 (0–0) | 15,925 (15,911–15,948) | 24,451 (24,412–24,485) | 32,497 (32,329–32,643) | 34,712 (34,537–34,878) | 35,763 (35,597–35,938) |
| Turkmenistan | 6,207 (6,204–6,212) | 0 (0–0) | 0 (0–0) | 995 (976–1,011) | 1,325 (1,305–1,344) | 5,614 (5,593–5,634) |
| Tuvalu | 5 (5–5) | 0 (0–0) | 3 (2–3) | 5 (5–6) | 6 (6–6) | 9 (9–9) |
| Uganda | 195,700 (193,364–198,794) | 7,822 (6,865–8,439) | 9,723 (8,762–10,386) | 28,816 (26,252–31,232) | 39,346 (35,502–42,982) | 223,998 (218,735–228,074) |
| Ukraine | 0 (0–0) | 16,950 (16,933–16,975) | 26,940 (26,897–26,977) | 34,946 (34,780–35,089) | 37,036 (36,865–37,195) | 37,419 (37,260–37,590) |
| Uzbekistan | 37,916 (37,889–37,954) | 0 (0–0) | 6,802 (6,761–6,833) | 18,228 (17,996–18,412) | 21,420 (21,185–21,625) | 51,840 (51,616–52,072) |
| Vanuatu | 246 (245–246) | 105 (105–105) | 277 (276–278) | 452 (448–456) | 499 (494–503) | 513 (509–519) |
| Venezuela | 0 (0–0) | 28,486 (27,835–29,257) | 42,370 (41,129–43,668) | 57,028 (53,820–60,091) | 62,382 (58,565–66,183) | 76,100 (72,299–79,916) |
| Vietnam | 0 (0–0) | 49,890 (49,807–49,991) | 75,889 (75,706–76,030) | 104,978 (104,265–105,507) | 113,569 (112,671–114,222) | 97,787 (97,134–98,685) |
| West Bank and Gaza | 0 (0–0) | 1,126 (1,125–1,128) | 1,715 (1,712–1,717) | 2,227 (2,216–2,237) | 2,351 (2,340–2,362) | 2,197 (2,186–2,208) |
| Yemen | 0 (0–0) | 7,509 (7,500–7,526) | 10,731 (10,677–10,824) | 13,398 (13,266–13,575) | 14,228 (14,072–14,423) | 14,360 (14,165–14,515) |
| Zambia | 89,201 (87,411–92,112) | 27,694 (26,365–28,362) | 40,644 (39,606–41,560) | 67,466 (63,515–71,614) | 76,655 (71,714–81,702) | 196,751 (190,073–202,505) |
| Zimbabwe | 128,432 (126,556–131,215) | 11,050 (10,539–11,430) | 15,836 (15,244–16,397) | 28,180 (26,231–30,009) | 36,075 (33,203–38,797) | 165,426 (161,207–168,683) |

**Table S2: Cervical cancer deaths averted (with 95%UI) by past vaccination and a one-time female catch-up HPV vaccination (with a vaccine targeting HPV16/18 and 90% coverage) up to age 14, 18, 25, or 30 in the lifetime of birth cohort aged 9–30 years in 2026.** The 2^nd^ to the 5^th^ columns from left to right correspond to the cumulative impact of adding each catch-up age band in the lifetime of the entire birth cohorts aged 9-30 years in 2026. The impact is the sum of direct and herd effect. The last column is the burden remaining after past vaccination and a catch-up up to age 30. Model outcomes were summarized across the 100 best fitting parameter sets obtained by model calibration, reporting medians and 95% uncertainty intervals (UI).

| **All LMICs / WHO Region / Country** | **Cervical cancer deaths averted**  (in the lifetime of birth cohort aged 9–30 years in 2026, with 95%UI) | | | | | **Cervical cancer deaths remaining** |
| --- | --- | --- | --- | --- | --- | --- |
|  | **Past vaccination** | **Catch-up  ages 9-14** | **Catch-up  ages 15-18** | **Catch-up  ages 19-25** | **Catch-up  ages 26-30** |  |
| All 132 LMICs | 1,566,682 (1,552,996–1,583,045) | 1,379,586 (1,377,306–1,381,144) | 2,263,480 (2,247,328–2,278,377) | 3,234,344 (3,190,536–3,282,338) | 3,555,756 (3,504,256–3,615,918) | 5,198,306 (5,134,551–5,255,105) |
| African | 927,352 (916,966–942,797) | 530,112 (528,564–531,381) | 855,978 (840,692–870,820) | 1,209,978 (1,169,710–1,254,492) | 1,336,877 (1,289,195–1,392,730) | 2,706,458 (2,643,142–2,760,521) |
| South-East Asia | 211,412 (210,944–211,901) | 404,158 (403,850–404,537) | 705,480 (704,970–706,044) | 1,058,086 (1,055,801–1,060,413) | 1,156,233 (1,153,140–1,159,228) | 1,099,756 (1,096,610–1,103,221) |
| Western Pacific | 53,724 (53,487–53,975) | 259,787 (259,552–260,065) | 401,943 (401,664–402,248) | 549,553 (548,444–550,640) | 591,574 (590,056–592,981) | 576,142 (574,655–577,816) |
| Americas | 328,091 (321,443–339,616) | 54,739 (53,240–54,962) | 94,637 (91,674–96,281) | 141,365 (131,617–147,967) | 177,169 (161,082–187,312) | 480,482 (460,388–496,959) |
| Eastern Mediterranean | 7,768 (7,735–7,805) | 108,579 (108,344–108,820) | 163,755 (163,073–164,509) | 212,112 (210,719–213,623) | 225,464 (223,913–227,165) | 236,811 (235,079–238,363) |
| European | 37,597 (37,567–37,647) | 22,398 (22,382–22,424) | 41,931 (41,829–41,999) | 63,599 (63,115–63,959) | 69,485 (69,017–69,896) | 99,571 (99,138–100,046) |
| Afghanistan | 0 (0–0) | 11,108 (11,096–11,122) | 16,550 (16,538–16,566) | 21,812 (21,784–21,846) | 23,045 (23,007–23,093) | 15,822 (15,773–15,859) |
| Albania | 282 (282–283) | 147 (147–147) | 293 (293–294) | 523 (519–526) | 603 (599–607) | 1,172 (1,167–1,176) |
| Algeria | 0 (0–0) | 8,596 (8,587–8,615) | 12,170 (12,108–12,275) | 14,767 (14,632–14,952) | 15,594 (15,440–15,796) | 19,220 (19,019–19,375) |
| Angola | 15,336 (14,850–15,872) | 29,118 (28,927–29,287) | 44,700 (43,359–45,989) | 57,047 (54,278–59,998) | 61,033 (57,904–64,434) | 91,478 (87,986–94,685) |
| Argentina | 34,794 (34,125–35,965) | 3,074 (2,656–3,218) | 3,678 (3,217–3,838) | 4,002 (3,557–4,170) | 5,860 (5,375–6,126) | 29,162 (28,355–29,781) |
| Armenia | 390 (389–391) | 177 (177–178) | 298 (297–298) | 477 (473–479) | 531 (527–534) | 874 (871–878) |
| Azerbaijan | 0 (0–0) | 1,273 (1,272–1,275) | 1,915 (1,912–1,918) | 2,446 (2,434–2,456) | 2,579 (2,567–2,590) | 2,317 (2,306–2,328) |
| Bangladesh | 41,963 (41,727–42,219) | 35,496 (35,351–35,638) | 63,867 (63,725–64,016) | 107,025 (106,956–107,087) | 118,381 (118,279–118,525) | 130,844 (130,445–131,150) |
| Belarus | 732 (731–734) | 489 (489–489) | 1,126 (1,123–1,128) | 1,646 (1,635–1,655) | 1,781 (1,770–1,791) | 2,475 (2,464–2,486) |
| Belize | 323 (316–335) | 75 (72–76) | 111 (107–113) | 222 (202–236) | 267 (241–285) | 644 (615–670) |
| Benin | 3,014 (2,919–3,111) | 5,138 (5,100–5,173) | 8,028 (7,776–8,271) | 10,419 (9,891–10,985) | 11,230 (10,627–11,884) | 17,633 (16,963–18,244) |
| Bhutan | 1,197 (1,197–1,198) | 0 (0–0) | 1 (1–1) | 1 (1–1) | 15 (15–15) | 414 (413–415) |
| Bolivia | 15,422 (15,121–15,935) | 1,458 (1,343–1,496) | 2,492 (2,355–2,556) | 4,877 (4,446–5,130) | 6,489 (5,834–6,917) | 22,011 (21,172–22,730) |
| Bosnia and Herzegovina | 15 (15–15) | 361 (360–361) | 608 (607–609) | 864 (859–868) | 941 (936–945) | 1,194 (1,189–1,199) |
| Botswana | 6,895 (6,836–7,020) | 1,043 (1,006–1,059) | 1,614 (1,567–1,638) | 2,091 (2,035–2,138) | 2,636 (2,567–2,715) | 8,123 (7,963–8,178) |
| Brazil | 100,088 (98,629–102,519) | 2,076 (1,834–2,199) | 3,332 (3,030–3,490) | 8,744 (7,825–9,289) | 17,298 (15,121–18,853) | 105,686 (102,602–108,295) |
| Burkina Faso | 12,792 (12,764–12,829) | 2,418 (2,406–2,439) | 8,855 (8,741–9,059) | 14,000 (13,750–14,361) | 15,521 (15,232–15,903) | 35,186 (34,817–35,475) |
| Burundi | 0 (0–0) | 16,698 (16,485–16,940) | 22,816 (22,253–23,359) | 27,363 (26,290–28,462) | 28,803 (27,594–30,107) | 33,987 (32,683–35,196) |
| Cabo Verde | 280 (280–281) | 0 (0–0) | 84 (82–87) | 196 (191–202) | 239 (233–247) | 767 (760–773) |
| Cambodia | 5,185 (5,173–5,200) | 2,296 (2,294–2,298) | 6,287 (6,262–6,310) | 10,816 (10,705–10,903) | 12,213 (12,072–12,317) | 15,360 (15,258–15,500) |
| Cameroon | 13,904 (13,567–14,513) | 17,481 (17,282–17,598) | 28,589 (27,759–29,246) | 37,861 (36,014–39,815) | 41,051 (38,859–43,286) | 68,154 (65,587–70,482) |
| Central African Republic | 0 (0–0) | 4,533 (4,472–4,599) | 6,237 (6,078–6,390) | 7,677 (7,356–8,003) | 8,175 (7,811–8,574) | 10,506 (10,108–10,870) |
| Chad | 0 (0–0) | 19,587 (19,565–19,631) | 27,976 (27,833–28,223) | 34,524 (34,192–34,969) | 36,385 (36,010–36,871) | 45,296 (44,809–45,671) |
| China | 0 (0–0) | 198,013 (197,796–198,271) | 301,764 (301,532–302,052) | 402,611 (402,093–403,273) | 430,157 (429,414–431,125) | 403,883 (402,915–404,627) |
| Colombia | 24,217 (23,551–25,500) | 4,069 (3,794–4,108) | 7,639 (7,201–7,841) | 10,462 (9,811–10,822) | 11,683 (10,903–12,089) | 27,747 (26,570–28,762) |
| Comoros | 151 (145–160) | 847 (840–854) | 1,221 (1,188–1,248) | 1,540 (1,472–1,613) | 1,659 (1,580–1,745) | 2,381 (2,292–2,461) |
| Congo | 0 (0–0) | 7,358 (7,263–7,464) | 10,073 (9,822–10,314) | 12,134 (11,652–12,626) | 12,815 (12,271–13,406) | 15,458 (14,867–16,003) |
| Congo Democratic Republic | 0 (0–0) | 139,881 (138,115–141,836) | 188,359 (183,782–192,794) | 227,226 (218,332–236,368) | 240,370 (230,275–251,427) | 291,600 (280,542–301,694) |
| Costa Rica | 1,141 (1,119–1,167) | 21 (21–22) | 151 (138–161) | 563 (488–621) | 737 (637–813) | 2,219 (2,119–2,313) |
| Cote d'Ivoire | 30,672 (30,221–31,113) | 4,370 (4,293–4,439) | 13,732 (12,875–14,546) | 23,910 (21,935–26,106) | 27,492 (25,245–30,039) | 73,716 (71,045–76,086) |
| Cuba | 900 (855–950) | 1,171 (1,157–1,187) | 2,179 (2,091–2,262) | 3,278 (3,040–3,488) | 3,740 (3,444–3,998) | 5,894 (5,607–6,185) |
| Djibouti | 22 (22–22) | 434 (434–435) | 658 (654–664) | 866 (856–879) | 933 (921–947) | 1,382 (1,368–1,394) |
| Dominica | 24 (23–24) | 2 (2–2) | 5 (5–5) | 15 (13–17) | 20 (18–22) | 57 (54–59) |
| Dominican Republic | 4,369 (4,244–4,586) | 2,394 (2,353–2,404) | 3,865 (3,770–3,922) | 5,979 (5,571–6,277) | 6,925 (6,381–7,331) | 12,895 (12,307–13,442) |
| Ecuador | 16,071 (15,816–16,506) | 956 (845–999) | 1,145 (1,021–1,197) | 1,814 (1,670–1,887) | 3,010 (2,712–3,194) | 15,738 (15,304–16,110) |
| Egypt | 0 (0–0) | 7,078 (7,069–7,093) | 10,111 (10,059–10,200) | 12,577 (12,453–12,739) | 13,353 (13,209–13,533) | 17,367 (17,187–17,511) |
| El Salvador | 2,562 (2,491–2,645) | 564 (560–568) | 1,889 (1,776–1,981) | 3,507 (3,166–3,786) | 4,262 (3,825–4,620) | 9,005 (8,580–9,427) |
| Equatorial Guinea | 0 (0–0) | 906 (895–919) | 1,229 (1,199–1,258) | 1,475 (1,417–1,534) | 1,549 (1,484–1,618) | 1,806 (1,736–1,870) |
| Eritrea | 2,315 (2,309–2,320) | 628 (626–631) | 874 (869–885) | 1,722 (1,693–1,760) | 1,995 (1,958–2,042) | 5,669 (5,621–5,710) |
| Eswatini | 1,186 (1,173–1,212) | 1,322 (1,311–1,331) | 2,525 (2,510–2,563) | 3,831 (3,770–3,968) | 4,321 (4,243–4,500) | 6,790 (6,589–6,861) |
| Ethiopia | 74,288 (74,044–74,485) | 30,594 (30,532–30,701) | 58,455 (57,941–59,392) | 94,529 (93,022–96,574) | 105,897 (104,103–108,229) | 242,498 (240,202–244,424) |
| Fiji | 1,981 (1,977–1,985) | 24 (23–24) | 54 (54–55) | 373 (366–379) | 561 (550–570) | 1,828 (1,818–1,841) |
| Gabon | 0 (0–0) | 1,323 (1,306–1,342) | 1,791 (1,747–1,834) | 2,165 (2,080–2,253) | 2,294 (2,197–2,401) | 2,812 (2,706–2,909) |
| Gambia | 2,348 (2,337–2,356) | 2,786 (2,782–2,794) | 3,980 (3,961–4,017) | 5,497 (5,435–5,582) | 5,968 (5,894–6,063) | 10,484 (10,388–10,565) |
| Georgia | 742 (741–744) | 335 (334–335) | 685 (684–687) | 1,036 (1,029–1,042) | 1,154 (1,147–1,161) | 1,845 (1,838–1,853) |
| Ghana | 14,935 (14,481–15,335) | 20,124 (19,905–20,326) | 33,056 (31,887–34,178) | 44,205 (41,753–46,851) | 48,075 (45,280–51,129) | 80,997 (77,930–83,789) |
| Grenada | 11 (11–12) | 29 (29–30) | 39 (38–40) | 57 (54–60) | 65 (61–69) | 97 (92–101) |
| Guatemala | 11,785 (11,464–12,283) | 5,544 (5,467–5,567) | 9,787 (9,460–9,998) | 16,623 (15,300–17,626) | 19,210 (17,528–20,500) | 36,016 (34,315–37,664) |
| Guinea | 0 (0–0) | 13,020 (13,004–13,050) | 18,863 (18,763–19,034) | 23,671 (23,431–23,984) | 25,232 (24,951–25,581) | 33,725 (33,376–34,006) |
| Guinea-Bissau | 0 (0–0) | 2,592 (2,589–2,598) | 3,778 (3,757–3,812) | 4,770 (4,721–4,834) | 5,085 (5,028–5,156) | 6,851 (6,780–6,909) |
| Haiti | 0 (0–0) | 6,607 (6,460–6,783) | 9,755 (9,475–10,050) | 12,995 (12,279–13,682) | 14,201 (13,347–15,053) | 17,131 (16,278–17,985) |
| Honduras | 8,935 (8,771–9,212) | 679 (625–699) | 1,125 (1,061–1,157) | 2,435 (2,200–2,580) | 3,331 (2,970–3,580) | 12,324 (11,857–12,721) |
| India | 0 (0–0) | 344,098 (343,690–344,586) | 540,358 (539,910–540,927) | 749,435 (748,379–750,816) | 804,472 (802,956–806,480) | 619,034 (617,026–620,550) |
| Indonesia | 115,558 (115,307–115,855) | 7,086 (7,061–7,117) | 60,811 (60,541–61,111) | 128,373 (126,831–129,637) | 148,812 (146,810–150,348) | 228,971 (227,450–231,032) |
| Iran | 0 (0–0) | 5,417 (5,412–5,424) | 8,147 (8,134–8,157) | 10,351 (10,302–10,392) | 10,880 (10,831–10,926) | 9,632 (9,586–9,681) |
| Iraq | 0 (0–0) | 5,538 (5,533–5,545) | 8,288 (8,276–8,299) | 10,614 (10,563–10,657) | 11,155 (11,103–11,203) | 9,966 (9,917–10,017) |
| Jamaica | 549 (531–582) | 813 (801–826) | 1,359 (1,320–1,392) | 2,008 (1,883–2,123) | 2,324 (2,155–2,474) | 3,801 (3,622–3,974) |
| Jordan | 0 (0–0) | 1,237 (1,236–1,239) | 1,864 (1,861–1,867) | 2,360 (2,349–2,370) | 2,482 (2,471–2,493) | 2,194 (2,183–2,205) |
| Kazakhstan | 2,399 (2,395–2,404) | 2,851 (2,849–2,852) | 5,583 (5,569–5,591) | 7,645 (7,598–7,682) | 8,106 (8,059–8,145) | 8,826 (8,783–8,873) |
| Kenya | 38,553 (37,882–39,434) | 10,223 (10,001–10,349) | 22,589 (21,633–23,458) | 36,802 (34,277–39,496) | 41,635 (38,715–44,826) | 101,307 (97,661–104,522) |
| Kiribati | 58 (58–58) | 8 (8–8) | 22 (22–22) | 50 (49–50) | 58 (57–59) | 84 (83–85) |
| Korea Dem. People's Rep. | 0 (0–0) | 5,479 (5,473–5,486) | 8,465 (8,458–8,473) | 11,701 (11,685–11,721) | 12,698 (12,675–12,729) | 13,274 (13,243–13,297) |
| Kosovo | 0 (0–0) | 159 (159–159) | 264 (264–265) | 366 (364–367) | 397 (395–399) | 485 (483–487) |
| Kyrgyzstan | 4,266 (4,263–4,271) | 98 (98–98) | 515 (513–517) | 1,776 (1,753–1,795) | 2,100 (2,077–2,121) | 5,583 (5,559–5,608) |
| Laos | 3,438 (3,431–3,446) | 195 (194–196) | 515 (513–517) | 2,110 (2,080–2,136) | 2,611 (2,569–2,644) | 5,347 (5,313–5,390) |
| Lebanon | 0 (0–0) | 521 (520–521) | 762 (761–763) | 981 (976–985) | 1,038 (1,033–1,043) | 982 (978–987) |
| Lesotho | 3,203 (3,175–3,268) | 1,117 (1,095–1,123) | 1,564 (1,551–1,573) | 2,991 (2,930–3,097) | 3,556 (3,478–3,713) | 7,773 (7,563–7,843) |
| Liberia | 3,613 (3,530–3,761) | 2,698 (2,631–2,728) | 4,416 (4,295–4,511) | 6,169 (5,841–6,485) | 6,752 (6,376–7,126) | 12,665 (12,198–13,081) |
| Libya | 875 (872–877) | 519 (518–520) | 1,086 (1,076–1,103) | 1,617 (1,593–1,650) | 1,769 (1,741–1,805) | 3,505 (3,469–3,535) |
| Madagascar | 14,212 (13,780–14,601) | 19,467 (19,267–19,653) | 32,087 (30,958–33,170) | 42,820 (40,454–45,370) | 46,709 (43,998–49,670) | 79,100 (76,120–81,813) |
| Malawi | 92,708 (90,886–95,257) | 10,031 (9,579–10,283) | 11,983 (11,649–12,273) | 29,535 (27,350–31,859) | 37,041 (34,060–40,116) | 156,538 (151,237–161,117) |
| Malaysia | 20,434 (20,374–20,482) | 194 (189–200) | 936 (916–957) | 1,158 (1,136–1,181) | 1,369 (1,349–1,388) | 10,289 (10,257–10,333) |
| Maldives | 202 (202–202) | 7 (7–7) | 14 (14–14) | 53 (52–54) | 73 (72–74) | 176 (174–177) |
| Mali | 12,826 (12,777–12,882) | 15,836 (15,809–15,891) | 28,257 (28,029–28,647) | 37,973 (37,488–38,667) | 40,695 (40,141–41,421) | 66,711 (65,993–67,265) |
| Marshall Islands | 39 (39–39) | 2 (2–3) | 7 (7–7) | 14 (14–14) | 19 (19–19) | 43 (43–43) |
| Mauritania | 2,605 (2,595–2,614) | 2,412 (2,409–2,420) | 3,984 (3,955–4,036) | 5,490 (5,420–5,589) | 5,943 (5,862–6,049) | 10,599 (10,495–10,685) |
| Mauritius | 313 (308–318) | 21 (19–21) | 25 (24–26) | 95 (86–105) | 130 (117–144) | 600 (584–614) |
| Mexico | 66,510 (65,076–68,998) | 4,033 (3,690–4,134) | 12,781 (11,947–13,249) | 15,746 (14,632–16,315) | 21,532 (19,742–22,503) | 74,961 (72,322–77,053) |
| Micronesia Fed. Sts. | 78 (78–79) | 9 (9–9) | 12 (12–12) | 27 (26–27) | 37 (36–37) | 90 (89–91) |
| Moldova | 747 (746–749) | 310 (309–310) | 476 (475–476) | 806 (800–811) | 920 (914–926) | 1,833 (1,827–1,840) |
| Mongolia | 683 (682–685) | 723 (723–724) | 1,416 (1,413–1,418) | 1,949 (1,937–1,958) | 2,089 (2,077–2,099) | 2,978 (2,967–2,990) |
| Montenegro | 28 (28–29) | 99 (99–99) | 174 (174–174) | 245 (243–246) | 265 (263–266) | 333 (332–334) |
| Morocco | 241 (240–242) | 7,409 (7,400–7,427) | 11,171 (11,107–11,281) | 14,374 (14,217–14,578) | 15,492 (15,305–15,722) | 22,425 (22,196–22,613) |
| Mozambique | 46,311 (45,630–47,112) | 7,096 (6,909–7,207) | 25,858 (24,457–27,138) | 40,992 (37,922–44,355) | 46,488 (42,986–50,357) | 114,248 (110,071–118,000) |
| Myanmar | 18,956 (18,914–19,006) | 4,657 (4,653–4,662) | 13,132 (13,075–13,190) | 27,081 (26,758–27,336) | 31,345 (30,932–31,659) | 45,829 (45,524–46,247) |
| Namibia | 670 (658–690) | 2,894 (2,881–2,913) | 4,368 (4,346–4,438) | 5,815 (5,742–5,993) | 6,361 (6,269–6,586) | 8,002 (7,763–8,083) |
| Nepal | 5,591 (5,582–5,601) | 3,923 (3,921–3,924) | 9,168 (9,164–9,172) | 15,001 (14,981–15,028) | 16,692 (16,659–16,735) | 18,656 (18,602–18,698) |
| Nicaragua | 2,757 (2,676–2,859) | 885 (871–890) | 2,590 (2,457–2,690) | 4,345 (3,966–4,645) | 5,056 (4,585–5,429) | 9,477 (9,018–9,933) |
| Niger | 0 (0–0) | 17,159 (17,140–17,198) | 24,483 (24,358–24,699) | 30,252 (29,960–30,641) | 31,913 (31,582–32,340) | 40,001 (39,574–40,332) |
| Nigeria | 60,057 (59,844–60,295) | 56,966 (56,886–57,175) | 109,915 (108,969–111,537) | 152,713 (150,620–155,648) | 165,210 (162,818–168,347) | 291,713 (288,627–294,156) |
| North Macedonia | 547 (546–548) | 78 (78–79) | 151 (151–152) | 222 (220–223) | 251 (250–252) | 608 (606–610) |
| Pakistan | 6,599 (6,567–6,634) | 23,989 (23,984–23,993) | 40,048 (40,038–40,057) | 55,032 (54,983–55,099) | 58,716 (58,637–58,817) | 46,078 (45,939–46,186) |
| Papua New Guinea | 0 (0–0) | 10,290 (10,271–10,310) | 15,583 (15,546–15,611) | 21,259 (21,117–21,361) | 22,697 (22,526–22,820) | 15,882 (15,760–16,054) |
| Paraguay | 9,420 (9,203–9,822) | 1,821 (1,657–1,868) | 2,071 (1,886–2,119) | 2,674 (2,504–2,741) | 3,568 (3,304–3,704) | 11,560 (11,209–11,853) |
| Peru | 27,552 (26,975–28,543) | 2,835 (2,682–2,893) | 5,565 (5,301–5,723) | 10,058 (9,166–10,588) | 13,285 (11,968–14,156) | 41,852 (40,202–43,281) |
| Philippines | 20,637 (20,473–20,800) | 22,229 (22,165–22,319) | 35,837 (35,757–35,927) | 54,266 (53,956–54,622) | 60,273 (59,807–60,696) | 68,578 (68,129–69,159) |
| Rwanda | 35,834 (35,426–36,481) | 192 (171–208) | 518 (484–548) | 996 (920–1,072) | 2,093 (1,881–2,275) | 27,687 (26,990–28,119) |
| Samoa | 47 (47–47) | 1 (1–1) | 5 (5–5) | 25 (24–25) | 31 (30–31) | 58 (57–58) |
| Sao Tome and Principe | 0 (0–0) | 185 (182–187) | 254 (248–261) | 316 (303–330) | 336 (321–353) | 437 (420–452) |
| Senegal | 22,836 (22,775–22,893) | 5,947 (5,935–5,974) | 11,982 (11,861–12,197) | 20,514 (20,160–20,994) | 23,128 (22,708–23,676) | 58,380 (57,846–58,831) |
| Serbia | 32 (32–32) | 1,126 (1,125–1,127) | 1,804 (1,801–1,807) | 2,450 (2,437–2,461) | 2,629 (2,616–2,642) | 3,000 (2,987–3,013) |
| Sierra Leone | 5,902 (5,797–6,016) | 1,626 (1,609–1,640) | 4,464 (4,240–4,669) | 6,915 (6,422–7,453) | 7,761 (7,201–8,375) | 17,586 (16,932–18,176) |
| Solomon Islands | 915 (911–919) | 202 (201–203) | 466 (465–468) | 805 (799–813) | 926 (916–934) | 1,272 (1,263–1,283) |
| Somalia | 0 (0–0) | 15,450 (15,247–15,668) | 21,011 (20,487–21,518) | 25,686 (24,639–26,753) | 27,181 (26,004–28,472) | 33,644 (32,354–34,822) |
| South Africa | 52,619 (52,229–53,401) | 5,680 (5,529–5,741) | 10,267 (10,054–10,407) | 17,481 (17,010–18,035) | 22,828 (22,186–23,850) | 73,896 (72,302–74,418) |
| South Sudan | 0 (0–0) | 8,665 (8,655–8,686) | 12,744 (12,675–12,864) | 15,954 (15,792–16,170) | 16,895 (16,711–17,133) | 21,996 (21,758–22,180) |
| Sri Lanka | 12,573 (12,546–12,600) | 656 (651–662) | 1,107 (1,100–1,114) | 4,136 (4,075–4,188) | 5,446 (5,355–5,517) | 11,578 (11,495–11,682) |
| St. Lucia | 64 (63–66) | 2 (2–2) | 12 (11–13) | 40 (35–44) | 54 (47–60) | 158 (151–165) |
| St. Vincent and the Grenadines | 8 (8–8) | 26 (25–26) | 41 (40–42) | 58 (55–62) | 67 (62–71) | 98 (94–103) |
| Sudan | 0 (0–0) | 21,132 (21,107–21,181) | 30,723 (30,559–31,003) | 38,746 (38,349–39,262) | 41,232 (40,769–41,807) | 55,074 (54,500–55,538) |
| Suriname | 485 (471–518) | 272 (261–274) | 340 (326–343) | 406 (390–414) | 471 (448–481) | 850 (818–876) |
| Syrian Arab Republic | 0 (0–0) | 2,646 (2,643–2,650) | 4,554 (4,546–4,561) | 6,089 (6,057–6,117) | 6,456 (6,423–6,488) | 6,382 (6,350–6,415) |
| Tajikistan | 1,847 (1,844–1,851) | 1,630 (1,628–1,631) | 3,347 (3,339–3,354) | 4,718 (4,686–4,743) | 5,070 (5,038–5,097) | 6,178 (6,149–6,209) |
| Tanzania | 91,964 (91,116–93,090) | 637 (622–654) | 2,092 (1,975–2,205) | 19,750 (17,499–22,307) | 25,789 (22,867–29,034) | 129,735 (125,760–133,097) |
| Thailand | 14,686 (14,624–14,749) | 2,613 (2,598–2,630) | 8,273 (8,240–8,311) | 14,654 (14,524–14,791) | 17,531 (17,328–17,703) | 29,758 (29,572–29,996) |
| Timor-Leste | 681 (679–684) | 147 (145–148) | 307 (306–309) | 700 (694–707) | 816 (806–824) | 1,291 (1,283–1,302) |
| Togo | 6,898 (6,781–7,054) | 1,840 (1,812–1,861) | 3,407 (3,274–3,534) | 5,577 (5,209–5,980) | 6,318 (5,879–6,800) | 15,951 (15,382–16,452) |
| Tonga | 11 (11–11) | 16 (16–16) | 31 (31–31) | 49 (49–49) | 54 (54–55) | 52 (52–53) |
| Tunisia | 32 (31–32) | 961 (960–963) | 1,398 (1,390–1,411) | 1,747 (1,729–1,770) | 1,874 (1,853–1,900) | 2,588 (2,562–2,609) |
| Turkey | 0 (0–0) | 6,403 (6,398–6,413) | 9,831 (9,816–9,845) | 13,067 (12,999–13,125) | 13,957 (13,887–14,024) | 14,380 (14,313–14,450) |
| Turkmenistan | 3,551 (3,549–3,553) | 0 (0–0) | 0 (0–0) | 569 (558–579) | 758 (747–769) | 3,211 (3,200–3,223) |
| Tuvalu | 2 (2–2) | 0 (0–0) | 1 (1–1) | 2 (2–2) | 3 (3–3) | 4 (4–4) |
| Uganda | 121,188 (119,742–123,104) | 4,844 (4,251–5,226) | 6,021 (5,426–6,431) | 17,844 (16,257–19,340) | 24,365 (21,985–26,617) | 138,712 (135,453–141,236) |
| Ukraine | 0 (0–0) | 6,864 (6,857–6,874) | 10,909 (10,892–10,925) | 14,152 (14,084–14,210) | 14,998 (14,929–15,062) | 15,153 (15,089–15,222) |
| Uzbekistan | 22,021 (22,005–22,043) | 0 (0–0) | 3,950 (3,927–3,968) | 10,587 (10,452–10,694) | 12,440 (12,304–12,560) | 30,108 (29,978–30,242) |
| Vanuatu | 218 (218–219) | 94 (94–94) | 246 (246–247) | 403 (399–406) | 444 (440–448) | 456 (453–461) |
| Venezuela | 0 (0–0) | 15,289 (14,940–15,703) | 22,741 (22,075–23,438) | 30,608 (28,887–32,252) | 33,482 (31,433–35,522) | 40,845 (38,805–42,893) |
| Vietnam | 0 (0–0) | 25,487 (25,444–25,538) | 38,768 (38,675–38,841) | 53,629 (53,265–53,899) | 58,018 (57,559–58,351) | 49,955 (49,622–50,414) |
| West Bank and Gaza | 0 (0–0) | 419 (419–420) | 638 (637–639) | 829 (825–832) | 875 (871–879) | 817 (814–822) |
| Yemen | 0 (0–0) | 4,715 (4,710–4,726) | 6,738 (6,704–6,797) | 8,413 (8,330–8,524) | 8,934 (8,836–9,056) | 9,017 (8,894–9,115) |
| Zambia | 55,839 (54,719–57,661) | 17,336 (16,504–17,754) | 25,443 (24,793–26,016) | 42,233 (39,760–44,830) | 47,985 (44,892–51,145) | 123,165 (118,984–126,766) |
| Zimbabwe | 81,173 (79,987–82,932) | 6,984 (6,661–7,224) | 10,009 (9,635–10,363) | 17,811 (16,579–18,967) | 22,801 (20,985–24,521) | 104,554 (101,888–106,613) |

**Table S3: The total budget (in USD$) required for the catch-up campaign extending to different maximum catch-up ages.** Financial cost, before any Gavi contribution, in constant 2024 United States dollars. Each column is the total budget for a campaign reaching that age, not the increment over the column before it. Both vaccine purchase and delivery costs are included..

| **All LMICs / WHO Region / Country** | **Vaccination Budget (US$) Up to different maximum ages of catch-up** | | | |
| --- | --- | --- | --- | --- |
|  | **Up to age 14** | **Up to age 18** | **Up to age 25** | **Up to age 30** |
| All 132 LMICs | 2,669,335,908 | 4,705,315,199 | 8,469,016,376 | 11,348,779,572 |
| African | 373,827,824 | 712,872,017 | 1,385,994,173 | 1,834,508,112 |
| South-East Asia | 543,832,965 | 1,050,101,304 | 2,096,032,797 | 2,858,534,110 |
| Western Pacific | 1,092,972,264 | 1,798,462,737 | 2,995,091,388 | 3,901,765,309 |
| Americas | 103,202,369 | 202,915,442 | 398,464,114 | 712,507,838 |
| Eastern Mediterranean | 407,223,672 | 682,974,661 | 1,133,642,823 | 1,429,333,678 |
| European | 148,276,812 | 257,989,038 | 459,791,081 | 612,130,526 |
| Afghanistan | 21,881,296 | 36,076,410 | 59,506,028 | 74,341,785 |
| Albania | 1,334,304 | 2,296,108 | 5,371,648 | 8,301,433 |
| Algeria | 56,564,207 | 88,791,088 | 133,900,477 | 164,259,718 |
| Angola | 14,943,571 | 26,781,459 | 45,321,571 | 56,045,300 |
| Argentina | 9,885,501 | 11,890,840 | 13,990,103 | 32,576,497 |
| Armenia | 1,783,523 | 2,627,102 | 4,989,575 | 6,984,326 |
| Azerbaijan | 9,796,148 | 15,617,263 | 24,553,624 | 31,088,644 |
| Bangladesh | 26,051,078 | 57,611,393 | 147,997,642 | 211,864,652 |
| Belarus | 3,744,914 | 7,878,210 | 14,334,125 | 18,977,119 |
| Belize | 93,815 | 148,232 | 438,011 | 643,766 |
| Benin | 5,226,144 | 9,679,629 | 16,646,454 | 20,825,533 |
| Bhutan | 68,830 | 70,656 | 74,016 | 194,458 |
| Bolivia | 1,452,807 | 2,967,225 | 9,468,175 | 16,598,291 |
| Bosnia and Herzegovina | 1,667,232 | 3,035,722 | 5,877,821 | 8,203,231 |
| Botswana | 454,894 | 734,523 | 1,153,870 | 1,892,967 |
| Brazil | 8,074,863 | 13,302,560 | 58,552,593 | 164,442,604 |
| Burkina Faso | 2,386,786 | 10,545,752 | 22,979,599 | 30,407,190 |
| Burundi | 7,800,714 | 12,861,921 | 20,018,273 | 24,043,688 |
| Cabo Verde | 0 | 104,970 | 389,528 | 612,372 |
| Cambodia | 2,407,915 | 7,072,887 | 15,250,674 | 21,380,328 |
| Cameroon | 8,736,378 | 17,789,508 | 32,628,257 | 41,494,593 |
| Central African Republic | 3,276,643 | 5,493,995 | 9,075,076 | 11,270,315 |
| Chad | 10,251,098 | 16,758,273 | 26,404,556 | 31,899,131 |
| China | 1,004,658,280 | 1,648,245,362 | 2,734,177,323 | 3,553,848,960 |
| Colombia | 10,572,956 | 23,009,339 | 38,637,946 | 53,373,446 |
| Comoros | 366,559 | 628,628 | 1,086,748 | 1,393,308 |
| Congo | 3,151,289 | 5,157,737 | 8,189,387 | 9,978,984 |
| Congo Democratic Republic | 56,747,316 | 90,131,096 | 143,380,089 | 175,295,155 |
| Costa Rica | 80,993 | 678,477 | 3,841,404 | 6,280,167 |
| Cote d'Ivoire | 3,026,324 | 11,890,482 | 29,918,024 | 40,962,177 |
| Cuba | 2,830,844 | 5,769,897 | 11,523,512 | 15,929,699 |
| Djibouti | 400,344 | 698,978 | 1,276,805 | 1,645,413 |
| Dominica | 4,236 | 11,539 | 58,693 | 98,824 |
| Dominican Republic | 3,931,120 | 6,925,322 | 14,785,037 | 21,014,325 |
| Ecuador | 2,282,012 | 2,804,925 | 6,691,121 | 16,380,369 |
| Egypt | 49,108,471 | 78,845,156 | 129,577,592 | 162,421,427 |
| El Salvador | 867,672 | 3,372,926 | 8,785,177 | 13,410,962 |
| Equatorial Guinea | 2,587,000 | 4,132,469 | 6,301,846 | 7,465,490 |
| Eritrea | 605,604 | 946,912 | 3,077,056 | 4,481,748 |
| Eswatini | 302,632 | 698,671 | 1,466,384 | 1,991,878 |
| Ethiopia | 22,624,319 | 51,374,925 | 123,557,639 | 169,882,336 |
| Fiji | 64,591 | 124,909 | 968,827 | 1,968,960 |
| Gabon | 3,472,193 | 5,501,786 | 8,590,375 | 10,499,502 |
| Gambia | 977,005 | 1,557,808 | 2,996,940 | 3,909,081 |
| Georgia | 1,222,059 | 2,860,586 | 6,094,897 | 9,017,682 |
| Ghana | 10,764,340 | 20,450,194 | 37,303,721 | 47,672,827 |
| Grenada | 52,292 | 78,610 | 157,854 | 221,070 |
| Guatemala | 6,331,154 | 12,422,573 | 29,192,354 | 40,807,778 |
| Guinea | 6,939,278 | 11,600,163 | 19,048,151 | 23,943,688 |
| Guinea-Bissau | 1,045,430 | 1,731,712 | 2,915,793 | 3,681,408 |
| Haiti | 4,594,622 | 7,715,816 | 13,673,864 | 17,849,497 |
| Honduras | 2,174,329 | 3,270,715 | 9,129,452 | 15,531,702 |
| India | 441,739,862 | 761,465,764 | 1,406,987,096 | 1,864,817,845 |
| Indonesia | 40,049,176 | 145,895,181 | 350,911,405 | 499,327,479 |
| Iran | 86,133,704 | 137,686,246 | 216,215,323 | 269,170,125 |
| Iraq | 65,975,172 | 105,065,467 | 167,787,613 | 207,804,210 |
| Jamaica | 907,814 | 1,791,207 | 3,745,478 | 5,464,500 |
| Jordan | 4,122,332 | 6,639,761 | 10,701,914 | 13,510,438 |
| Kazakhstan | 13,438,208 | 27,591,383 | 47,438,610 | 60,107,431 |
| Kenya | 7,026,286 | 20,555,135 | 51,613,275 | 70,170,076 |
| Kiribati | 13,314 | 34,046 | 97,069 | 141,850 |
| Korea Dem. People's Rep. | 6,151,783 | 10,176,121 | 18,857,929 | 26,173,622 |
| Kosovo | 1,507,499 | 2,721,135 | 5,007,561 | 6,967,186 |
| Kyrgyzstan | 569,771 | 1,042,601 | 3,860,127 | 5,892,630 |
| Laos | 360,146 | 931,531 | 4,894,690 | 7,867,092 |
| Lebanon | 2,081,049 | 3,233,428 | 5,433,096 | 7,014,137 |
| Lesotho | 449,483 | 680,585 | 1,907,892 | 2,789,253 |
| Liberia | 1,433,771 | 2,901,199 | 5,771,488 | 7,441,681 |
| Libya | 4,457,708 | 9,467,337 | 18,475,765 | 23,770,956 |
| Madagascar | 10,068,758 | 19,801,513 | 35,607,399 | 45,761,145 |
| Malawi | 2,500,295 | 3,158,775 | 13,906,305 | 21,698,368 |
| Malaysia | 15,972,111 | 25,193,778 | 28,603,430 | 36,649,313 |
| Maldives | 15,482 | 28,893 | 126,529 | 240,176 |
| Mali | 7,234,679 | 15,771,762 | 28,479,346 | 35,668,588 |
| Marshall Islands | 17,222 | 42,135 | 103,903 | 182,555 |
| Mauritania | 1,517,340 | 2,874,022 | 5,390,420 | 6,952,297 |
| Mauritius | 145,138 | 183,353 | 1,347,450 | 2,361,093 |
| Mexico | 21,380,936 | 56,876,014 | 77,640,124 | 148,571,626 |
| Micronesia Fed. Sts. | 16,057 | 22,334 | 63,645 | 120,970 |
| Moldova | 1,120,662 | 1,900,506 | 4,998,048 | 7,783,789 |
| Mongolia | 2,528,826 | 5,035,088 | 8,678,661 | 11,363,754 |
| Montenegro | 356,251 | 666,857 | 1,249,491 | 1,696,558 |
| Morocco | 10,165,656 | 18,899,949 | 34,670,062 | 46,032,444 |
| Mozambique | 3,025,506 | 14,677,589 | 32,832,988 | 44,377,486 |
| Myanmar | 4,769,229 | 13,590,417 | 36,681,884 | 53,518,442 |
| Namibia | 3,513,305 | 5,904,841 | 9,731,344 | 12,394,009 |
| Nepal | 5,470,415 | 13,055,829 | 29,463,826 | 42,298,434 |
| Nicaragua | 1,357,814 | 4,517,835 | 10,299,837 | 14,598,217 |
| Niger | 14,223,943 | 23,538,619 | 37,370,979 | 45,462,548 |
| Nigeria | 59,643,683 | 130,926,192 | 247,187,417 | 314,594,531 |
| North Macedonia | 697,682 | 1,190,240 | 2,091,227 | 3,160,749 |
| Pakistan | 95,540,943 | 173,313,729 | 299,993,066 | 384,650,652 |
| Papua New Guinea | 4,232,647 | 6,948,892 | 11,653,718 | 14,480,499 |
| Paraguay | 2,231,732 | 2,620,641 | 4,488,788 | 8,633,651 |
| Peru | 4,643,363 | 9,890,034 | 25,452,851 | 44,604,599 |
| Philippines | 33,460,492 | 56,540,835 | 107,326,645 | 145,361,557 |
| Rwanda | 2,368,555 | 2,772,796 | 3,937,078 | 8,046,152 |
| Samoa | 6,464 | 24,604 | 155,574 | 249,858 |
| Sao Tome and Principe | 120,174 | 197,233 | 339,734 | 423,468 |
| Senegal | 2,605,883 | 6,133,954 | 15,688,786 | 21,660,980 |
| Serbia | 3,778,846 | 6,473,638 | 11,518,068 | 15,390,923 |
| Sierra Leone | 996,338 | 3,668,259 | 8,304,330 | 11,094,127 |
| Solomon Islands | 122,507 | 282,066 | 600,039 | 851,872 |
| Somalia | 9,856,969 | 16,071,897 | 26,247,523 | 32,003,071 |
| South Africa | 17,788,880 | 33,700,902 | 75,406,114 | 125,878,284 |
| South Sudan | 6,208,306 | 10,578,058 | 17,245,652 | 21,194,052 |
| Sri Lanka | 2,227,077 | 2,929,694 | 9,708,848 | 15,908,558 |
| St. Lucia | 4,029 | 27,653 | 142,365 | 246,574 |
| St. Vincent and the Grenadines | 43,486 | 83,140 | 164,524 | 235,542 |
| Sudan | 24,098,518 | 39,781,031 | 66,888,559 | 83,982,717 |
| Suriname | 335,499 | 451,301 | 642,626 | 983,713 |
| Syrian Arab Republic | 8,697,237 | 16,699,622 | 29,728,218 | 38,321,264 |
| Tajikistan | 2,422,765 | 5,337,869 | 9,960,639 | 13,286,848 |
| Tanzania | 585,248 | 2,253,450 | 38,058,919 | 59,641,990 |
| Thailand | 17,088,980 | 44,883,868 | 94,067,533 | 142,496,410 |
| Timor-Leste | 201,054 | 393,487 | 1,156,090 | 1,694,033 |
| Togo | 1,581,572 | 3,381,308 | 7,981,002 | 10,744,958 |
| Tonga | 85,955 | 194,176 | 399,628 | 542,973 |
| Tunisia | 3,283,005 | 5,728,578 | 9,885,485 | 13,045,549 |
| Turkey | 77,387,149 | 126,088,662 | 217,641,788 | 286,950,895 |
| Turkmenistan | 0 | 0 | 2,281,368 | 4,119,175 |
| Tuvalu | 1,580 | 8,989 | 24,724 | 36,834 |
| Uganda | 2,110,226 | 3,098,077 | 23,180,452 | 40,743,481 |
| Ukraine | 27,449,799 | 46,859,860 | 75,897,542 | 97,559,999 |
| Uzbekistan | 0 | 3,801,296 | 16,624,923 | 26,641,909 |
| Vanuatu | 58,286 | 151,971 | 305,246 | 404,187 |
| Venezuela | 19,068,480 | 32,288,621 | 56,962,227 | 74,010,418 |
| Vietnam | 28,965,871 | 47,609,134 | 81,787,590 | 106,313,749 |
| West Bank and Gaza | 2,616,023 | 4,248,423 | 7,241,310 | 9,221,856 |
| Yemen | 18,805,245 | 30,518,649 | 50,014,463 | 62,397,632 |
| Zambia | 4,236,240 | 7,407,792 | 18,741,950 | 25,675,598 |
| Zimbabwe | 2,194,492 | 3,362,901 | 9,614,039 | 15,825,556 |

**Table S4: Country-specific vaccine prices, delivery costs by age group, and five-year current health expenditure and immunization budgets**

| **Country** | **Vaccine Procurement Mechanism/category ^a^** | **GAVI eligibility category ᶜ** | **Vaccine Price per dose  (US$) ᵃ ᶜ** | **Vaccine delivery cost  (ages 9–14, US$) ᵇ ᶜ** | **Vaccine delivery cost  (ages 15–18, US$) ᵇ ᶜ** | **Vaccine delivery cost  (ages 19–30, US$) ᵇ ᶜ** | **5-year Current Health Expenditure (2024 US$) ᵈ** | **5-year Immunization Budget (2024 US$) ᵈ** |
| --- | --- | --- | --- | --- | --- | --- | --- | --- |
| Afghanistan | Gavi | Initial self-financing | 4,50 | 2,13 | 2,90 | 3,19 | 13 854 203 789 | 196 343 558 |
| Albania | Self-procurement UMIC | NA | 19,58 | 2,13 | 2,26 | 3,19 | 6 295 754 501 | 17 085 096 |
| Algeria | Self-procurement UMIC | NA | 19,58 | 2,13 | 2,32 | 3,19 | 56 600 804 477 | 1 169 500 000 |
| Angola | Former Gavi LMIC | Preparatory transition | 4,50 | 2,13 | 2,51 | 3,19 | 25 201 326 602 | 365 858 453 |
| Argentina | PAHO RF | NA | 10,48 | 2,13 | 2,19 | 3,19 | 295 546 232 045 | 1 199 371 026 |
| Armenia | Self-procurement UMIC | Fully self-financing | 19,58 | 2,13 | 2,18 | 3,19 | 10 002 080 719 | ND |
| Azerbaijan | Self-procurement UMIC | Fully self-financing | 19,58 | 2,13 | 2,23 | 3,19 | 15 473 445 162 | 13 204 877 |
| Bangladesh | Gavi | Accelerated transition | 4,50 | 2,13 | 2,41 | 3,19 | 56 137 416 636 | 564 432 779 |
| Belarus | Self-procurement UMIC | NA | 19,58 | 2,13 | 2,14 | 3,19 | 24 511 252 989 | ND |
| Belize | PAHO RF | Catalytic countries | 10,48 | 2,13 | 2,50 | 3,19 | 666 057 058 | 67 186 840 |
| Benin | Gavi | Preparatory transition | 4,50 | 2,13 | 2,83 | 3,19 | 2 113 286 433 | 72 242 376 |
| Bhutan | Former Gavi LMIC | Catalytic countries | 4,50 | 2,13 | 2,56 | 3,19 | 523 619 220 | 6 199 607 |
| Bolivia | PAHO RF | Catalytic countries | 10,48 | 2,13 | 2,25 | 3,19 | 17 026 993 012 | 108 748 222 |
| Bosnia and Herzegovina | Self-procurement UMIC | NA | 19,58 | 2,13 | 2,19 | 3,19 | 11 185 675 117 | ND |
| Botswana | UNICEF SD | NA | 4,50 | 2,13 | 2,43 | 3,19 | 5 953 309 726 | 45 443 676 |
| Brazil | PAHO RF | NA | 10,48 | 2,13 | 2,21 | 3,19 | 939 656 948 689 | ND |
| Burkina Faso | Gavi | Initial self-financing | 4,50 | 2,13 | 2,97 | 3,19 | 6 105 226 582 | 205 368 516 |
| Burundi | Gavi | Initial self-financing | 4,50 | 2,13 | 2,71 | 3,19 | 2 572 230 189 | 122 176 368 |
| Cabo Verde | UNICEF SD | Catalytic countries | 4,50 | 2,13 | 2,65 | 3,19 | 647 724 846 | ND |
| Cambodia | Gavi | Preparatory transition | 4,50 | 2,13 | 2,79 | 3,19 | 9 944 420 583 | 61 713 323 |
| Cameroon | Gavi | Preparatory transition | 4,50 | 2,13 | 2,65 | 3,19 | 9 605 256 100 | 140 694 568 |
| Central African Republic | Gavi | Initial self-financing | 4,50 | 2,13 | 2,81 | 3,19 | 1 177 034 107 | 28 186 368 |
| Chad | Gavi | Initial self-financing | 4,50 | 2,13 | 2,92 | 3,19 | 3 631 126 751 | 107 252 440 |
| China | Self-procurement UMIC | NA | 19,58 | 2,13 | 2,27 | 3,19 | 4 380 353 492 674 | 3 454 602 923 |
| Colombia | PAHO RF | NA | 10,48 | 2,13 | 2,30 | 3,19 | 146 939 174 961 | 644 329 574 |
| Comoros | Gavi | Preparatory transition | 4,50 | 2,13 | 2,44 | 3,19 | 351 164 851 | 5 601 736 |
| Congo | Gavi | Accelerated transition | 4,50 | 2,13 | 2,46 | 3,19 | 1 892 464 576 | 33 336 864 |
| Congo Democratic Republic | Gavi | Initial self-financing | 4,50 | 2,13 | 2,49 | 3,19 | 9 166 128 797 | 533 286 722 |
| Costa Rica | PAHO RF | NA | 10,48 | 2,13 | 2,21 | 3,19 | 24 273 621 630 | 82 190 388 |
| Cote d'Ivoire | Gavi | Accelerated transition | 4,50 | 2,13 | 2,83 | 3,19 | 12 215 045 186 | 234 930 877 |
| Cuba | PAHO RF | Fully self-financing | 10,48 | 2,13 | 2,32 | 3,19 | 73 475 644 607 | 236 545 378 |
| Djibouti | Gavi | Accelerated transition | 4,50 | 2,13 | 2,65 | 3,19 | 452 928 055 | ND |
| Dominica | PAHO RF | Catalytic countries | 10,48 | 2,13 | 2,32 | 3,19 | 191 515 705 | 14 222 157 |
| Dominican Republic | PAHO RF | NA | 10,48 | 2,13 | 2,30 | 3,19 | 25 285 522 390 | 112 756 231 |
| Ecuador | PAHO RF | NA | 10,48 | 2,13 | 2,29 | 3,19 | 45 560 892 766 | 3 849 971 407 |
| Egypt | Former Gavi LMIC | Catalytic countries | 4,50 | 2,13 | 2,42 | 3,19 | 148 202 127 150 | ND |
| El Salvador | PAHO RF | NA | 10,48 | 2,13 | 2,50 | 3,19 | 14 466 578 091 | 90 064 621 |
| Equatorial Guinea | Self-procurement UMIC | NA | 19,58 | 2,13 | 2,65 | 3,19 | 1 962 752 000 | 7 914 052 |
| Eritrea | Gavi | Initial self-financing | 4,50 | 2,13 | 2,65 | 3,19 | 535 766 541 | 20 681 524 |
| Eswatini | Former Gavi LMIC | Catalytic countries | 4,50 | 2,13 | 2,38 | 3,19 | 1 570 361 652 | ND |
| Ethiopia | Gavi | Initial self-financing | 4,50 | 2,13 | 2,76 | 3,19 | 27 772 782 406 | 1 261 709 410 |
| Fiji | Self-procurement UMIC | Catalytic countries | 19,58 | 2,13 | 2,38 | 3,19 | 1 413 067 764 | ND |
| Gabon | Self-procurement UMIC | NA | 19,58 | 2,13 | 2,33 | 3,19 | 2 655 908 075 | 6 310 572 |
| Gambia | Gavi | Initial self-financing | 4,50 | 2,13 | 2,72 | 3,19 | 457 767 982 | 18 503 147 |
| Georgia | Self-procurement UMIC | Fully self-financing | 19,58 | 2,13 | 2,22 | 3,19 | 8 723 105 071 | 52 897 506 |
| Ghana | Gavi | Accelerated transition | 4,50 | 2,13 | 2,44 | 3,19 | 20 960 052 647 | 663 998 632 |
| Grenada | PAHO RF | Catalytic countries | 10,48 | 2,13 | 2,32 | 3,19 | 308 613 834 | 56 945 704 |
| Guatemala | PAHO RF | NA | 10,48 | 2,13 | 2,70 | 3,19 | 32 423 929 742 | 194 517 395 |
| Guinea | Gavi | Preparatory transition | 4,50 | 2,13 | 2,91 | 3,19 | 4 385 475 779 | ND |
| Guinea-Bissau | Gavi | Initial self-financing | 4,50 | 2,13 | 2,51 | 3,19 | 717 060 575 | 8 390 921 |
| Haiti | Gavi | Preparatory transition | 4,50 | 2,13 | 2,29 | 3,19 | 6 317 883 214 | 61 216 377 |
| Honduras | PAHO RF | Catalytic countries | 10,48 | 2,13 | 2,61 | 3,19 | 13 533 974 950 | 425 566 681 |
| India | Former Gavi LMIC | Fully self-financing | 4,50 | 2,13 | 2,40 | 3,19 | 544 231 093 600 | 2 628 306 818 |
| Indonesia | Self-procurement UMIC | Fully self-financing | 11,53 | 2,13 | 2,33 | 3,19 | 193 103 434 917 | 776 884 137 |
| Iran | Self-procurement UMIC | NA | 19,58 | 2,13 | 2,40 | 3,19 | 193 935 913 249 | ND |
| Iraq | Self-procurement UMIC | NA | 19,58 | 2,13 | 2,62 | 3,19 | 45 577 850 681 | ND |
| Jamaica | PAHO RF | NA | 10,48 | 2,13 | 2,20 | 3,19 | 6 175 719 937 | 7 901 216 |
| Jordan | Former Gavi LMIC | Catalytic countries | 4,50 | 2,13 | 2,31 | 3,19 | 17 263 825 555 | ND |
| Kazakhstan | Self-procurement UMIC | NA | 19,58 | 2,13 | 2,15 | 3,19 | 43 335 533 935 | 477 890 454 |
| Kenya | Gavi | Preparatory transition | 4,50 | 2,13 | 2,25 | 3,19 | 26 182 812 074 | 164 096 925 |
| Kiribati | Former Gavi LMIC | Catalytic countries | 4,50 | 2,13 | 2,45 | 3,19 | 157 090 016 | ND |
| Korea Dem. People's Rep. | Gavi | Initial self-financing | 4,50 | 2,13 | 2,14 | 3,19 | ND | ND |
| Kosovo | Self-procurement UMIC | Catalytic countries | 19,58 | 2,13 | 2,22 | 3,19 | ND | ND |
| Kyrgyzstan | Gavi | Preparatory transition | 4,50 | 2,13 | 2,27 | 3,19 | 2 832 987 775 | 11 537 121 |
| Laos | Gavi | Preparatory transition | 4,50 | 2,13 | 2,58 | 3,19 | 2 919 335 088 | 40 184 435 |
| Lebanon | Former Gavi LMIC | Catalytic countries | 4,50 | 2,13 | 2,40 | 3,19 | 95 410 005 543 | 224 071 447 |
| Lesotho | Gavi | Preparatory transition | 4,50 | 2,13 | 2,42 | 3,19 | 1 879 493 589 | 4 888 492 |
| Liberia | Gavi | Initial self-financing | 4,50 | 2,13 | 2,50 | 3,19 | 3 181 567 929 | 33 122 806 |
| Libya | Self-procurement UMIC | NA | 19,58 | 2,13 | 2,40 | 3,19 | 7 878 401 000 | ND |
| Madagascar | Gavi | Initial self-financing | 4,50 | 2,13 | 2,84 | 3,19 | 2 838 424 231 | 104 608 905 |
| Malawi | Gavi | Initial self-financing | 4,50 | 2,13 | 2,52 | 3,19 | 7 355 346 623 | 95 267 105 |
| Malaysia | Self-procurement UMIC | NA | 19,58 | 2,13 | 2,38 | 3,19 | 75 784 627 641 | ND |
| Maldives | UNICEF SD | Catalytic countries | 4,50 | 2,13 | 2,37 | 3,19 | 2 541 072 082 | 779 757 |
| Mali | Gavi | Initial self-financing | 4,50 | 2,13 | 2,98 | 3,19 | 3 526 267 533 | 115 319 525 |
| Marshall Islands | Self-procurement UMIC | Catalytic countries | 19,58 | 2,13 | 2,38 | 3,19 | 188 214 169 | 16 766 211 |
| Mauritania | Gavi | Preparatory transition | 4,50 | 2,13 | 2,77 | 3,19 | 1 753 842 127 | 35 208 782 |
| Mauritius | Self-procurement UMIC | NA | 19,58 | 2,13 | 2,65 | 3,19 | 4 209 095 358 | ND |
| Mexico | PAHO RF | NA | 10,48 | 2,13 | 2,38 | 3,19 | 447 910 178 831 | 999 033 912 |
| Micronesia Fed. Sts. | Former Gavi LMIC | Catalytic countries | 4,50 | 2,13 | 2,38 | 3,19 | 243 561 344 | 7 968 156 |
| Moldova | Self-procurement UMIC | Fully self-financing | 19,58 | 2,13 | 2,34 | 3,19 | 6 262 469 535 | ND |
| Mongolia | Self-procurement UMIC | Fully self-financing | 19,58 | 2,13 | 2,19 | 3,19 | 5 506 950 530 | 20 158 906 |
| Montenegro | Self-procurement UMIC | NA | 19,58 | 2,13 | 2,20 | 3,19 | 3 208 090 342 | 9 903 893 |
| Morocco | UNICEF SD | Catalytic countries | 4,50 | 2,13 | 2,40 | 3,19 | 37 153 485 710 | 243 533 317 |
| Mozambique | Gavi | Initial self-financing | 4,50 | 2,13 | 2,80 | 3,19 | 8 339 006 212 | 192 083 240 |
| Myanmar | Gavi | Preparatory transition | 4,50 | 2,13 | 2,65 | 3,19 | 13 561 130 945 | ND |
| Namibia | Self-procurement UMIC | Catalytic countries | 19,58 | 2,13 | 2,51 | 3,19 | 6 005 214 231 | 37 387 655 |
| Nepal | Gavi | Preparatory transition | 4,50 | 2,13 | 2,29 | 3,19 | 10 279 483 304 | 151 288 017 |
| Nicaragua | PAHO RF | Catalytic countries | 10,48 | 2,13 | 2,32 | 3,19 | 7 384 678 250 | 194 276 894 |
| Niger | Gavi | Initial self-financing | 4,50 | 2,13 | 3,10 | 3,19 | 4 234 593 691 | 484 317 124 |
| Nigeria | Gavi | Preparatory transition | 4,50 | 2,13 | 2,49 | 3,19 | 149 187 529 591 | 2 379 199 877 |
| North Macedonia | Self-procurement UMIC | NA | 19,58 | 2,13 | 2,25 | 3,19 | 5 821 822 722 | ND |
| Pakistan | Gavi | Preparatory transition | 4,50 | 2,13 | 2,71 | 3,19 | 67 158 147 347 | 1 640 839 898 |
| Papua New Guinea | Gavi | Accelerated transition | 4,50 | 2,13 | 2,38 | 3,19 | 3 420 369 571 | 65 831 660 |
| Paraguay | PAHO RF | NA | 10,48 | 2,13 | 2,41 | 3,19 | 16 532 482 494 | 228 983 733 |
| Peru | PAHO RF | NA | 10,48 | 2,13 | 2,30 | 3,19 | 74 549 570 528 | 1 096 791 089 |
| Philippines | Former Gavi LMIC | Catalytic countries | 4,50 | 2,13 | 2,18 | 3,19 | 107 519 817 057 | 627 794 220 |
| Rwanda | Gavi | Initial self-financing | 4,50 | 2,13 | 2,76 | 3,19 | 5 145 142 684 | 62 989 788 |
| Samoa | Former Gavi LMIC | Catalytic countries | 4,50 | 2,13 | 2,27 | 3,19 | 303 912 073 | ND |
| Sao Tome and Principe | Gavi | Accelerated transition | 4,50 | 2,13 | 2,30 | 3,19 | 258 110 885 | 1 861 598 |
| Senegal | Gavi | Preparatory transition | 4,50 | 2,13 | 2,72 | 3,19 | 6 223 394 221 | 107 689 175 |
| Serbia | Self-procurement UMIC | NA | 19,58 | 2,13 | 2,17 | 3,19 | 31 861 879 974 | ND |
| Sierra Leone | Gavi | Initial self-financing | 4,50 | 2,13 | 2,55 | 3,19 | 3 392 248 135 | 77 011 823 |
| Solomon Islands | Gavi | Preparatory transition | 4,50 | 2,13 | 2,38 | 3,19 | 418 567 808 | ND |
| Somalia | Gavi | Initial self-financing | 4,50 | 2,13 | 2,65 | 3,19 | 1 334 597 961 | 66 225 813 |
| South Africa | Self-procurement UMIC | NA | 19,58 | 2,13 | 2,31 | 3,19 | 188 008 228 085 | 1 012 256 968 |
| South Sudan | Gavi | Initial self-financing | 4,50 | 2,13 | 2,85 | 3,19 | 2 469 161 076 | ND |
| Sri Lanka | Former Gavi LMIC | Catalytic countries | 4,50 | 2,13 | 2,30 | 3,19 | 18 944 315 828 | ND |
| St. Lucia | PAHO RF | Catalytic countries | 10,48 | 2,13 | 2,28 | 3,19 | 546 572 815 | 564 207 |
| St. Vincent and the Grenadines | PAHO RF | Catalytic countries | 10,48 | 2,13 | 2,32 | 3,19 | 239 216 203 | 1 473 016 |
| Sudan | Gavi | Initial self-financing | 4,50 | 2,13 | 2,51 | 3,19 | 15 835 143 177 | 401 369 174 |
| Suriname | PAHO RF | Catalytic countries | 10,48 | 2,13 | 2,32 | 3,19 | 2 336 155 687 | 3 190 276 |
| Syrian Arab Republic | Gavi | Initial self-financing | 4,50 | 2,13 | 2,40 | 3,19 | 1 768 320 357 | 100 000 000 |
| Tajikistan | Gavi | Preparatory transition | 4,50 | 2,13 | 2,53 | 3,19 | 3 194 559 064 | 23 682 122 |
| Tanzania | Gavi | Preparatory transition | 4,50 | 2,13 | 2,99 | 3,19 | 13 019 620 732 | 633 315 095 |
| Thailand | Self-procurement UMIC | NA | 19,58 | 2,13 | 2,25 | 3,19 | 117 060 870 352 |  |
| Timor-Leste | Former Gavi LMIC | Preparatory transition | 4,50 | 2,13 | 2,28 | 3,19 | 918 458 842 | 21 566 882 |
| Togo | Gavi | Initial self-financing | 4,50 | 2,13 | 2,56 | 3,19 | 2 499 378 370 | 58 317 200 |
| Tonga | Self-procurement UMIC | Catalytic countries | 19,58 | 2,13 | 2,44 | 3,19 | 191 251 032 | 2 500 000 |
| Tunisia | Former Gavi LMIC | Catalytic countries | 4,50 | 2,13 | 2,35 | 3,19 | 17 799 504 737 | 108 993 888 |
| Turkey | Self-procurement UMIC | NA | 19,58 | 2,13 | 2,23 | 3,19 | 361 795 485 898 | ND |
| Turkmenistan | UNICEF SD | NA | 4,50 | 2,13 | 2,14 | 3,19 | 18 933 253 411 | ND |
| Tuvalu | Self-procurement UMIC | Catalytic countries | 19,58 | 2,13 | 2,52 | 3,19 | 51 431 320 | 2 831 810 |
| Uganda | Gavi | Initial self-financing | 4,50 | 2,13 | 2,85 | 3,19 | 10 849 311 449 | 263 563 362 |
| Ukraine | Self-procurement UMIC | Fully self-financing | 19,58 | 2,13 | 2,14 | 3,19 | 80 268 744 425 | ND |
| Uzbekistan | Former Gavi LMIC | Catalytic countries | 4,50 | 2,13 | 2,23 | 3,19 | 27 656 537 070 | 309 213 949 |
| Vanuatu | Former Gavi LMIC | Catalytic countries | 4,50 | 2,13 | 2,38 | 3,19 | 222 405 136 | 3 217 965 |
| Venezuela | PAHO RF | Catalytic countries | 10,48 | 2,13 | 2,32 | 3,19 | 7 561 910 341 | 12 720 653 |
| Vietnam | Former Gavi LMIC | Catalytic countries | 4,50 | 2,13 | 2,36 | 3,19 | 92 410 493 830 | 93 274 918 |
| West Bank and Gaza | Former Gavi LMIC | Catalytic countries | 4,50 | 2,13 | 2,26 | 3,19 | 14 320 097 610 | 69 393 394 |
| Yemen | Gavi | Initial self-financing | 4,50 | 2,13 | 2,70 | 3,19 | 4 548 534 450 | 189 925 696 |
| Zambia | Gavi | Preparatory transition | 4,50 | 2,13 | 2,78 | 3,19 | 10 632 078 752 | 221 374 717 |
| Zimbabwe | Gavi | Preparatory transition | 4,50 | 2,13 | 2,71 | 3,19 | 192 652 416 236 | 195 756 920 |

LIC: Low-income country, LMIC: lower-middle-income country, UMIC: upper-middle-income country, PAHO RF, Pan American Health organization Revolving Fund, UNICEF SD: United Nations Children's Fund Supply Division, NA: not applicable, ND: data not available

^a^ Vaccine procurement cost data was sourced from the Market Information for Access (MI4A) vaccine purchase database.^4^ Because MI4A reports prices anonymously by country, certain assumptions were applied to derive country-specific prices per dose. All Gavi-supported countries were assumed to pay $4.50 per dose. Non-Gavi countries procuring through UNICEF were assigned the UNICEF Supply Division price. Countries in the WHO's Americas Region (AMRO) were considered part of PAHO and assumed to purchase via the PAHO Revolving Fund at its average price. For other lower-middle-income countries, a price of $4.50 per dose was assumed. For upper-middle-income countries that self-procure, the median self-procurement price was used.

^b^ For the budget impact analysis, we used a financial cost which is one-third of the economic costs. School-based programs target in-school girls (9–14); outreach targets out-of-school girls and young women (19–30). Girls aged 15–18 may in theory be reached by either method. For this analysis we calculated the delivery cost for this age as a weighted average of school and outreach costs based on each country’s secondary school enrolment rate.
^c^ Co-financing assumptions for Gavi countries: For vaccine cost co-financing, we assumed country co-financing based on Gavi 6.0 2026 guidelines.^5^ Countries in the “initial self-financing phase” were assumed to pay 4% of the vaccine price with Gavi financing the rest, then up to 50% average co-financing for countries in the “preparatory transition phase”, then 90% co-financing for countries in the “accelerated transition phase”, and finally 100% co-financing by countries in the “catalytic phase”. For delivery cost co-financing, we based our per-dose cost assumptions on the Gavi 2022 guidelines ($0.45, $0.55, or $0.65 per girl, as per country co-financing phase), because in Gavi 6.0 there are no delivery costs per-dose specified. Also per the guidelines, this financial support was available for multi-age-cohort campaigns up to age 18, and for ages older than this we assumed no financial support from Gavi even in Gavi countries, though this policy may change in the future.

^d^ Country‑specific Current Health Expenditure (CHE) was obtained from the WHO Global Health Expenditure Database,^6^ and immunization budget data from the WHO/UNICEF Joint Reporting Form (JRF).^7^ Because JRF budget data were not consistently available for every year, we approximated each country’s 5‑year budget by: (i) extracting all available annual values from 2018 onward, (ii) inflating each to 2024 using country‑specific inflation rates, (iii) computing the mean of the adjusted values, and (iv) multiplying by five. All costs were expressed in 2024 UD$ by first applying the Consumer Price Index (CPI) inflation adjustment ^8^ and then converting local currencies using 2024 US$ exchange rates.^9^

**Table S5: Key input parameters and country profile data included in the cost-effectiveness analysis**

| **Data category** | **Colombia** | **Eswatini** | **India** | **Indonesia** | **Kenya** | **Nigeria** |
| --- | --- | --- | --- | --- | --- | --- |
| Total population size aged 9-30 in 2026 (in million) ^a^ | 8.8 | 0.27 | 263 | 49.3 | 13.4 | 54.2 |
| Income level ^b^ | Upper–middle | Lower–middle | Lower–middle | Upper–middle | Lower–middle | Lower–middle |
| GDP per capita in 2025 (USD$) ^b^ | 8,562 | 4,108 | 2,702 | 5,060 | 2,363 | 1,224 |
| Cervical cancer epidemiological profile ^c^ | Cluster IV | Cluster I | Cluster VI | Cluster V | Cluster II | Cluster III |
| Median age at first sexual intercourse (Women 20 – 49 years) ^d^ | 17.6 | 18.1 | 19.3 | – | 18.1 | 17.9 |
| Mean number of lifetime sexual partner (Women 15 – 49 years) ^d^ | 3.0 | 2.4 | 1.7 | – | 2.3 | 1.8 |
| Vaccine cost per dose (USD$) | 10.48^e^ | 4.50^f^ | 4.50^f^ | 11.53^g^ | 4.50^f^ | 4.50^f^ |
| Vaccine delivery cost by age group in years (USD$) ^h^ |  |  |  |  |  |  |
| Ages 9 – 14 ^i^ | 2.13 | 2.13 | 2.13 | 2.13 | 2.13 | 2.13 |
| Ages 15 – 18 ^j^ | 2.30 | 2.38 | 2.40 | 2.33 | 2.25 | 2.49 |
| Ages 19 – 30 ^k^ | 3.19 | 3.19 | 3.19 | 3.19 | 3.19 | 3.19 |
| Cervical cancer treatment cost (USD$) ^l^ | 3,063 ^10^ | 40,717 ^11^ | 3,882 ^12^ | 2,766 ^13-16^ | 4,148 ^17^ | 969 ^18^ |
| Age-standardized cervical cancer incidence  per 100,000 woman-years in 2024 ^m^ | 13.81 | 76.52 | 10.53 | 18.55 | 24 | 19.08 |
| Age-standardized cervical cancer mortality  per 100,000 woman-years in 2024 ^m^ | 6.4 | 44.3 | 4.86 | 9.29 | 14.87 | 12 |

^a^ United Nations population.^19^

^b^ World Bank Group.^20^

^c^ Countries are clustered based on their sexual behaviour data reported in a preprint.^1^ We did not include a country from Cluster VII, as none of the countries have high-quality primary cervical cancer treatment cost data.

^d^ Demographic and Health Surveys STATcompiler. Each country data from the most recent survey.^21^

^e^ PAHO revolving fund price.

^f^ Gavi price.

^g^ Price for a locally manufactured HPV vaccine.

^h^ Shows financial cost. For the cost-effectiveness analysis, we used the economic cost which is 3x the financial costs.

^i^ School based delivery cost.

^j^ School-based programs target in-school girls, while outreach program target on out-of-school girls. The delivery cost is calculated as the weighted average of the school-based and outreach delivery costs, based on each country's secondary school enrolment rate^22^

^k^ Outreach delivery cost.

^l^ Treatment costs were expressed in 2024 US$ by first applying the Consumer Price Index (CPI) inflation adjustment^8^ and then converting local currencies using 2024 US$ exchange rates^9^.

^m^ GLOBOCAN ^23^

GDP: Gross domestic product; USD$: United States dollars

**Table S6: HPV-FRAME checklist*^24^***

| **HPV-FRAME core reporting standards** | | | |
| --- | --- | --- | --- |
| **a) Inputs** | **Reported by age?**  **(Y/N)** | **Report by sex?**  **(F-only, M-only or both)** | **Comments** |
| Target population for intervention | Yes | F-only | Girls/women aged 9-30 years in 2026 (catch-up vaccination) |
| Sexual behaviour | Yes | Both | Derivation of the following parameters from surveys reported: age- and sex-specific partnership acquisition rates; age- and sex-specific age mixing matrices. Calibration of assortativeness parameters and type-specific probability of HPV transmission were reported (Reported in the previous publication^1^) |
| Cohort examined for evaluation / time horizon | No | Female only | Lifetime risk of cervical cancer was presented for the female cohorts aged 9-30 at 2026. |
| Quality of life assumptions | NA | NA | NA |
| Calibration | Yes | Both | Reported in the previous publication^25^ |
| Validation (where possible) | NA | NA | NA |
| Costs | Yes | F-only | All costs were converted to 2024 US dollars (USD) by first adjusting for inflation using country-specific Consumer Price Index data, followed by currency conversion using 2024 exchange rates. Future costs were discounted at 3% annual discount rate. |
| **b) Outputs** | **Reported? (Y/N)** | **Reported by age? (Y/N)** | **Report by sex? (F-only, M-only or both)** |
| Cancer incidence, mortality, life years, DALYs | Y | N | F-only |
| HPV prevalence, pre-intervention | Y | Y | Both |
| CIN2 detected | N | NA | NA |
| Sensitivity analysis on key inputs | Y | NA | F-only |
| Incremental cost-effectiveness ratios and costs saved | Y | NA | F-only |

| **Reporting standard for HPV vaccination in adolescent individuals** | | | | |
| --- | --- | --- | --- | --- |
| **a) Inputs** | **Reported?  (Y/N)** | **Reported by age? (Y/N)** | **Report by sex?  (F-only, M-only or both)** | **Report as calibration or validation target? (Y/N)** |
| Vaccine uptake | Yes | Yes | F-only | Past vaccination was simulated as comparator. In base case scenario 90% coverage was assumed for catch-up vaccination. Other vaccination coverage scenarios were also considered in the sensitivity analysis. |
| Vaccine efficacy | Yes | Yes | F-only | HPV type specific efficacy was considered. Efficacy was independent of age. |
| Vaccine cross-protection | Yes | NA | NA | Level of cross-protection for HPV 31/33/45 was reported independent of age. |
| Duration vaccine protection and waning | Yes | Yes | NA | Lifelong vaccine-acquired immunity was assumed independent of age. |
| Vaccine and delivery costs | Yes | Yes (for delivery cost) | NA | Country specific HPV vaccine prices were estimated based on the Market Information for Access (MI4A) vaccine purchase data. Delivery costs were stratified by age group and corresponding delivery strategy: school-based for ages 9–14, outreach-based for ages 19 and above, and mixed delivery for ages 15–18 depending on school enrolment status |
| Pre-vaccination disease burden (including population attributable fractions for HPV) | Yes | Yes | F-only | Sexual behaviour cluster and age-specific prevalence rates of HPV infection; Sexual behaviour cluster and age-specific incidence and mortality rates of detected cervical cancer and Sexual behaviour cluster -specific attributable fraction by HPV type to cervical cancer burden were reported. |
| Duration of natural immunity | Yes | NA | F-only | Age-independent natural immunity was reported. |
| **b) Outputs** | **Reported?  (Y/N)** | **Reported by age? (Y/N)** | **Report by sex? (F-only, M-only or both)** | **Comments** |
| Absolute reductions in HPV infections, and/or warts, post-vaccination | No | NA | NA | Model-derived estimates of HPV infection reduction were used to project reduction in cervical cancer risk but not reported. |
| Absolute reductions in CIN2+ post-vaccination | No | NA | NA | NA |
| Absolute reductions in invasive cancer (cervical and other HPV cancers, as relevant) | Yes,  for cervical cancer. | No | Female only,  cervical cancer. | Model-estimates of cervical cancer risk reduction were reported. |

| **Reporting standards for evaluations of vaccination at older ages** | | | | |
| --- | --- | --- | --- | --- |
| **a) Inputs** | **Detail** | **Reported by age? (Y/N)** | **Report by sex? (F-only, M-only or both)** | **Comments** |
| Natural history | Nature history structure used in the model (including progression and regression from high grade to low grade disease/productive HPV infection). | NA | Yes | Reported in the previous publication^25^ |
| Natural history | Rate of clearance of HPV infection | NA | Yes | Reported in the previous publication^25^ |
| Natural history | Rate of loss of naturally acquired immunity | NA | Yes | Reported in the previous publication^25^ |
| Natural history | Simulation of latency by HPV type (handling of apparently new infections in older women – is the possibility that some are reactivated latent infections explored in sensitivity analysis?) | NA | NA | No latency assumed in the model. |
| Vaccination | Vaccine coverage at older ages | Yes | NA | In base case scenario coverage for catch-up vaccination in females at older age was assumed to be 90%. Other scenarios with various coverage points were also used in the sensitivity analysis. |
| Vaccination | Whether screen-and-vaccinate is being modelled, or just vaccination at older ages (without linking to screening/HPV status) | NA | NA | We did not model screen-and-vaccinate. We modelled vaccination at older ages but without linking to screening/HPV status. |

| **Reporting standard for models of HPV prevention in LMIC** | | |  |  |
| --- | --- | --- | --- | --- |
| **a) Inputs** | **Reported? (Y/N)** | **Reported by age? (Y/N)** | **Report by sex?  (F-only, M-only or both)** | **Comments** |
| HIV prevalence rates if endemic in country | No | No | No | The effects of HIV are not modelled. |
| Description of any opportunistic or pilot/demonstration screening project ongoing | No | NA | NA | No screening was accounted in this study |

| **Reporting standards for evaluations assessing alternative vaccine types or reduced-dose schedules** | | | | |
| --- | --- | --- | --- | --- |
| **a) Inputs** | **Reported? (Y/N)** | **Reported by age? (Y/N)** | **Report by sex?  (F-only, M-only or both)** | **Comments** |
| Vaccine efficacy/waning | Y | Y | F-only | In the base case (scenario A), one-dose schedule was assumed to provide 95% protection with a lifelong duration against vaccine-targeted HPV types. In the sensitivity analyses, two single-dose long-term vaccine protection waning scenarios were considered. Scenario B: waning to ~80% efficacy 20 years postvaccination; scenario C: lower initial efficacies at approximately 87.5%, and waning to ~75% 20 years postvaccination. |
| Timing between doses (for 2-dose) | NA | NA | NA |  |
| Vaccine cross-protection | Y | N | F-only | Cross-protection for HPV 31/33/45 was modelled. Scenario A: 9% lifelong; scenarios B and C: initial efficacy of 9% and waning to 3% in 20 years post vaccination. |
| Cost | See Comments. | See Comments. | See Comments. | Cost per dose, delivery cost per dose was reported. |
| **b) Outputs** | **Reported? (Y/N)** | **Reported by age? (Y/N)** | **Report by sex? (F-only, M-only or both)** | **Report as calibration or validation target (Y/N)?** |
| Threshold cost per dose | N | N | N |  |

**References**

1.	Man I, Macacu A, Eynard M, et al. A unified modeling platform for informing cervical cancer prevention policy decisions in 132 low- and middle-income countries (DOI: 10.64898/2026.03.18.26348700). *medRxiv* 2026.

2.	Sayinzoga F, Umulisa MC, Sibomana H, Tenet V, Baussano I, Clifford GM. Human papillomavirus vaccine coverage in Rwanda: A population-level analysis by birth cohort. *Vaccine* 2020; **38**(24): 4001-5.

3.	World Health Organization. HPV Dashboard. <https://www.who.int/teams/immunization-vaccines-and-biologicals/diseases/human-papillomavirus-vaccines-(HPV)/hpv-clearing-house/hpv-dashboard> (accessed July 2026).

4.	World Health Organization. Market Information for Access (MI4A) vaccine purchase database. <https://www.who.int/teams/immunization-vaccines-and-biologicals/vaccine-access/mi4a/mi4a-vaccine-purchase-data> (accessed May 2025).

5.	Gavi. Gavi 6.0 funding guidelines. 2026. <https://www.gavi.org/country-support/guidelines>.

6.	World Health Organization. Global Health Expenditure Database. <https://apps.who.int/nha/database/Select/Indicators/en> (accessed May 2025).

7.	World Health Organization. WHO/UNICEF Joint Reporting Form <https://www.who.int/teams/immunization-vaccines-and-biologicals/immunization-analysis-and-insights/global-monitoring/who-unicef-joint-reporting-process> (accessed May 2025).

8.	World Bank Group. Consumer price index (2010 = 100). <https://data.worldbank.org/indicator/FP.CPI.TOTL> (accessed May 2025).

9.	World Bank Group. Official exchange rate (LCU per US$, period average). <https://data.worldbank.org/indicator/PA.NUS.FCRF> (accessed May 2025).

10.	Akhavan-Tabatabaei R, Sanchez DM, Yeung TG. A Markov Decision Process Model for Cervical Cancer Screening Policies in Colombia. *Med Decis Making* 2017; **37**(2): 196-211.

11.	Ngcamphalala C, Ostensson E, Ginindza TG. The economic burden of cervical cancer in Eswatini: Societal perspective. *Plos One* 2021; **16**(4): e0250113.

12.	M De Carvalho T, Man I, Georges D, et al. Health and economic effects of introducing single-dose or two-dose human papillomavirus vaccination in India. *BMJ Global Health* 2023; **8**(11): e012580.

13.	Endarti D, Riewpaiboon A, Thavorncharoensap M, Praditsitthikorn N, Hutubessy R, Kristina SA. Evaluation of Health-Related Quality of Life among Patients with Cervical Cancer in Indonesia. *Asian Pac J Cancer Prev* 2015; **16**(8): 3345-50.

14.	Puspitasari IM, Legianawati D, Sinuraya RK, Suwantika AA. Cost-Effectiveness Analysis of Chemoradiation and Radiotherapy Treatment for Stage IIB and IIIB Cervical Cancer Patients. *Int J Womens Health* 2021; **13**: 221-9.

15.	Satibi S, Andayani TM, Endarti D, Suwantara IPT, Agustini NPD. Comparison of Real Cost Versus the Indonesian Case Base Groups (INA-CBGs) Tariff Rates Among Patients of High- Incidence Cancers Under the National Health Insurance Scheme. *Asian Pac J Cancer Prev* 2019; **20**(1): 117-22.

16.	Setiawan D, Dolk FC, Suwantika AA, Westra TA, JC WI, Postma MJ. Cost-Utility Analysis of Human Papillomavirus Vaccination and Cervical Screening on Cervical Cancer Patient in Indonesia. *Value Health Reg Issues* 2016; **9**: 84-92.

17.	Subramanian S, Gakunga R, Kibachio J, et al. Cost and affordability of non-communicable disease screening, diagnosis and treatment in Kenya: Patient payments in the private and public sectors. *Plos One* 2018; **13**(1): e0190113.

18.	Demarteau N, Morhason-Bello IO, Akinwunmi B, Adewole IF. Modeling optimal cervical cancer prevention strategies in Nigeria. *BMC Cancer* 2014; **14**: 365.

19.	United Nations Department of Economic and Social Affairs Population Division. World Population Prospects 2024. <https://population.un.org/wpp/> (accessed 12/05/2025.

20.	World Bank Group. 2025. <https://datatopics.worldbank.org/world-development-indicators/the-world-by-income-and-region.html> (accessed July 2026).

21.	The DHS Program. Demographic and Health Surveys. <https://dhsprogram.com/> (accessed June 2025).

22.	UNICEF. Out-of-school rate for youth of upper secondary school age. <https://data.unicef.org/resources/data_explorer/unicef_f/?ag=UNICEF&df=GLOBAL_DATAFLOW&ver=1.0&dq=.ED_ROFST_L3..&startPeriod=1999&endPeriod=2023> (accessed 05/05/2025 2025).

23.	Ervik M, Laversanne M, Lam F, et al. Global Cancer Observatory: Cancer Today (2024 estimates, version 1.0). Lyon, France: International Agency for Research on Cancer. . 2026. <https://gco.iarc.who.int/today,https://dx.doi.org/10.3322/caac.21834>.

24.	Canfell K, Kim JJ, Kulasingam S, et al. HPV-FRAME: A consensus statement and quality framework for modelled evaluations of HPV-related cancer control. *Papillomavirus Res* 2019; **8**: 100184.

25.	Man I, Georges D, Basu P, Baussano I. Leveraging single-dose human papillomavirus vaccination dose-efficiency to attain cervical cancer elimination in resource-constrained settings. *J Natl Cancer Inst Monogr* 2024; **2024**(67): 400-9.
